# identifying cancer patients from GC-patterned fragment ends of cell-free DNA

**DOI:** 10.1101/2022.08.02.22278319

**Authors:** Samuel D. Curtis, Mahmoud Summers, Joshua D. Cohen, Yuxuan Wang, Nadine Nehme, Maria Popoli, Janine Ptak, Natalie Sillman, Lisa Dobbyn, Adam Buchanan, Jeanne Tie, Peter Gibbs, Lan T. Ho-Pham, Bich N. H. Tran, Shibin Zhou, Chetan Bettegowda, Anne Marie Lennon, Ralph H. Hruban, Kenneth W. Kinzler, Nickolas Papadopoulos, Bert Vogelstein, Christopher Douville

**Affiliations:** Ludwig Center, Johns Hopkins University School of Medicine, Baltimore, MD 21287; Sidney Kimmel Comprehensive Cancer Center, Johns Hopkins University School of Medicine, Baltimore, MD 21287; Sol Goldman Pancreatic Cancer Research Center, Johns Hopkins University School of Medicine, Baltimore, MD 21287; Howard Hughes Medical Institute, Johns Hopkins Medical Institutions, Baltimore, MD 21287; Department of Pharmacology and Molecular Science, Johns Hopkins School of Medicine, Baltimore, MD 21287; Department of Biomedical Engineering, Johns Hopkins University, Baltimore, MD 21218; Genomic Medicine Institute, Geisinger, Danville, PA 17822; Division of Personalised Oncology, Walter and Eliza Hall Institute of Medical Research, Melbourne, Australia; Department of Medical Oncology, Western Health, Melbourne, Australia; Faculty of Medicine, Dentistry and Health Sciences, University of Melbourne, Melbourne, Australia; Department of Medical Oncology, Peter MacCallum Cancer Centre, Melbourne, Australia; BioMedical Research Center, Pham Ngoc Thach University of Medicine; Saigon Precision Medicine Research Center, Vietnam; University of New South Wales, Australia; Department of Neurosurgery, Johns Hopkins Medical Institutions, Baltimore, MD 21287; Department of Medicine, Johns Hopkins Medical Institutions,, Baltimore, MD 21287; Department of Pathology, Johns Hopkins Medical Institutions,, Baltimore, MD 21287

## Abstract

One of the most intriguing characteristics of cell-free DNA (cfDNA) from plasma is the sequence at the ends of the fragments. Previous studies have shown that these end-sequences are somewhat different in cancer patients than in healthy individuals. While investigating this characteristic, we noticed that the bases at the 5’-ends of a double-stranded fragment were highly correlated with the GC content of that particular fragment. This led us to develop a method, called MendSeqS (Modified End-based sequencing System), that incorporates the correlation between end-motifs and GC content into the analysis of shallow (0.5x) whole genome sequencing (WGS). When applied to plasma samples, MendSeqS was able to classify patients with a sensitivity of 96% at 98% specificity in a cohort comprised of 107 individuals evaluated in our laboratory (43 with cancer and 64 without). In cohorts evaluated in three other laboratories, comprising a total of 401 individuals (193 with cancer and 208 without), MendSeqS achieved a sensitivity of 87% at 98% specificity. MendSeqS could in principle be combined with other methods of cfDNA analysis to enhance cancer detection.

## INTRODUCTION

The earlier detection of cancer has the potential to substantially reduce cancer morbidity and mortality because all cancer treatments are more successful when there’s a lower tumor burden in the patient^1-2^. The evaluation of cell-free DNA (cfDNA) from plasma is one of the most promising approaches for such earlier detection. Numerous ways to use cfDNA have been described in the literature. Genetic alterations in cfDNA – such as mutations or copy number alterations – have been extensively used for this purpose. Epigenetic alterations, in particular changes in DNA methylation, have also been used to identify patients with cancer^3-6^. Other types of epigenetic changes, reflecting chromatin organization rather than covalent modifications of DNA, have more recently gained attention^3, 7-12^. Because DNA is always wrapped in nucleosomes, whether in the cell or in the circulation, changes in chromatin structure result in changes of the fragments produced by nucleases in the cell of origin or in the circulation^11, 13-14^. This gives rise to different fragmentation patterns as well as differences in fragment sizes or the sequences at the ends of fragments. Because epigenetics, rather than genetics, is responsible for cell differentiation epigenetic patterns in plasma cfDNA can often reveal the cell of origin of the fragments.

Though the results to date of these cfDNA-based technologies are promising, further research to increase the sensitivity of cancer detection while maintaining high specificity is a research and clinical priority. We here report a new heuristic, inspired by previous studies of cfDNA fragmentation patterns in cancer patients, particularly studies on fragment end-motifs, for classifying patients based on data from shallow whole genome sequencing^13, 15-17^.

## RESULTS

We began by evaluating 43 cancer patients and 64 healthy individuals of similar age (Cohort 1, Table 1) Whole genome sequencing was performed on the cfDNA of each of these patients (Methods) to an average depth of 17 million high quality reads (∼0.5x genome coverage). Two key observations about the sequences of the bases at the ends of fragments were made during this initial evaluation, which led us to evaluate more patients and develop algorithms for classifying cancer patients based on them.

### Observation 1: End-motif frequency is influenced by local GC content

We first found that the frequencies of trimers at the 5’-ends of fragments correlated with the GC content of the entire fragment (i.e., the 3 base pairs at each of the two ends plus the ∼70 to 350 bp (average ∼170 bp) between the trimers). We made similar observations with the two bases at the ends (dimers) or the four bases at the ends (tetramers), but we focused on trimers in this study. We noticed a few general trends dependent on the GC content of the particular trimer (Fig. 1 and Supplementary Data 1). The frequency of fragments with ends containing two A:T bp and one G:C bp were *negatively* correlated with the GC content of the entire fragment (e.g., Fig. 1A). Conversely, the frequency of fragments with ends containing one A:T bp and two G:C bp were *positively* correlated with the GC content of the entire fragment (e.g., Fig. 1B). Figure 1C illustrates the trend when the trimer composition was extreme, with G:C bp at all three positions. Though a positive correlation with of the frequency of the extreme type of trimer and the GC content of the entire fragment was still evident, the relationship was exponential rather than linear. These trends were observed in plasma cfDNA derived from healthy individuals as well as from cancer patients in Cohort 1 (Fig. 1A to 1D) as well as in publicly available data^18^ (Fig 1 E to 1H). The GC-dependent frequencies of all 64 trimers in the cohorts used in this study are presented in Supplementary Table 2. To test whether these trends were specific to cell-free DNA, we performed in silico shearing of the hg19 reference genome. When the human genome was randomly sheared to the same fragment size distribution as cfDNA, the trends in end-motif trimer frequency as a function of GC content were similar to those of actual cfDNA (Fig 1 I to 1L).

**Figure 1.**
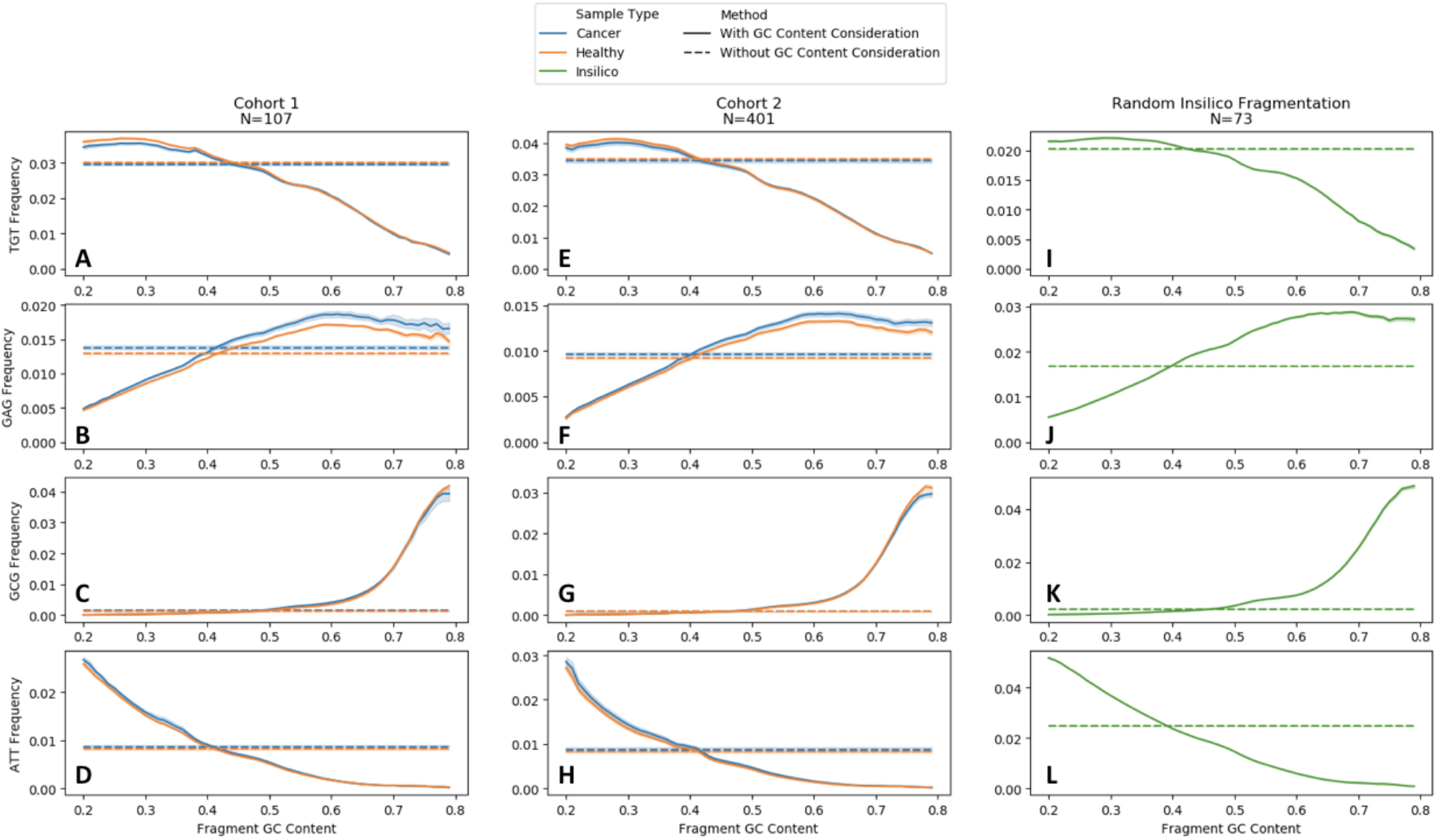
Correlations between end-motif frequencies and GC-contents of the fragment from which the ends were derived. (A-D) End-motif trimer frequencies in healthy individuals and cancer patients in Cohort 1. The dotted lines represent the frequencies of the indicated end-motif trimers regardless of the GC content of the fragment from which the trimer was derived and are therefore horizontal lines in this plot. The solid lines represent the frequencies of the indicated trimers as a function of the GC content (x-axis) of the fragments containing them. (E-H) Similar patterns were observed in cfDNA samples from Cohort 3 and in in silico-generated fragments from the hg19 genome (I-L).

### Observation 2: Binning end-motifs by fragment GC content improves the distinction between healthy individuals and cancer patients

It has previously been demonstrated that the frequencies of trimers at the 5’-ends of cfDNA fragments from healthy individuals is different than those derived from patients with cancer^13, 17, 19-20^. In this study, we observed that cancer-specific differences in end-motifs are substantially more pronounced if the GC content of the entire fragment is taken into account. This is illustrated in Fig. 2 for each of the four trimers shown in Fig. 1. For each trimer, the frequency of fragments containing that trimer was determined from the WGS data of Cohort 1. Using the frequencies in the 64 healthy individuals in this cohort as a reference distribution, Z-scores for each sample in Cohort 1 could be calculated, with a Z-score of zero corresponding to the average frequency of that trimer in the healthy individuals. The average Z-scores for the 43 cancer patients in Cohort 1 are represented by the horizontal blue lines in Fig. 2. Similarly, Z-scores could be calculated for each trimer present in bins of fragments with similar GC contents. Sixty such Z-scores were obtained for each trimer, corresponding to bins of 20% to 21% GC content, 21% to 22%, etc. all the way up to 79% to 80%. The average Z-scores for the 43 cancer patients in Cohort 1, as a function of the GC content of the underlying fragment (x-axis) are represented by the orange lines in Fig. 2.

**Figure 2.**
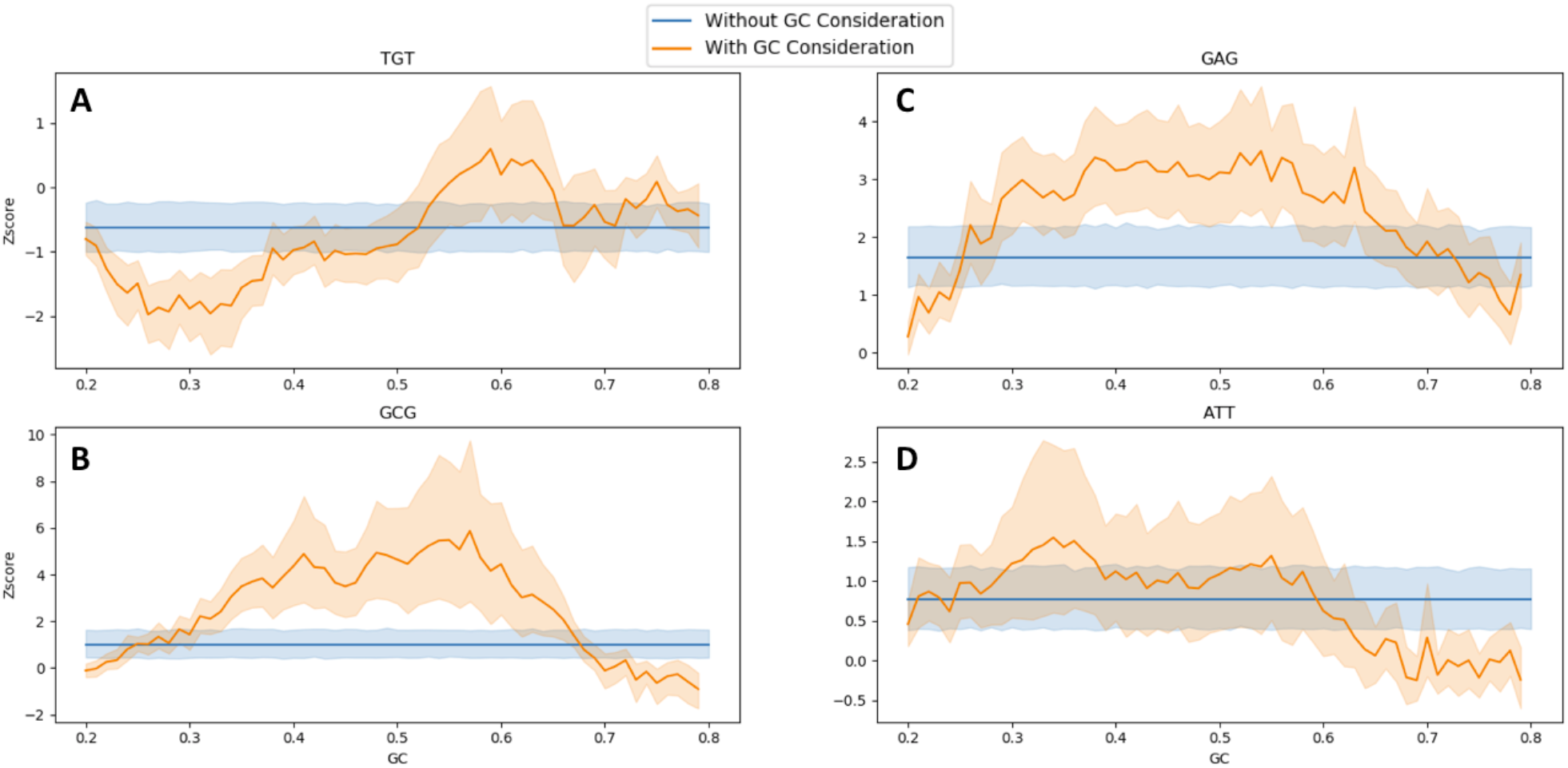
Comparisons of end-motif trimer frequencies in cfDNA fragments from cancer patients and healthy individuals with or without consideration of GC content. Average Z-scores (lines) and 95% confidence intervals (shaded) are shown for the 22 cancer patients in Cohort 1, using the trimer frequencies in the 64 healthy individuals in Cohort 1 as references. (A) Z-scores for TGT; (B) Z-scores for GCG (C); Z-scores for GAG; (D) Z-scores for ATT.

As an example, with trimer TGT the average Z-score was -0.619 without consideration of GC (blue line in Fig. 2A) while the average Z-scores ranged from -2.0 in fragments with a GC content of 29% to +0.6 in fragments with a GC content of 60% (orange line in Fig. 2A). The maximum absolute Z-score for the TGT trimer, when considering GC contents, was three times as high as the Z-score for the same trimer when GC contents were not considered. This translated to an improved distinction between cancer patients and healthy individuals with TGT trimers (Mann-Whitney p-values of 1.1e-11 *vs*. 0.002 with and without GC content consideration, respectively; Supplementary Table 2).

The distinction between cancer patients and normal individuals was also observed with trimers GAG and GCG, though the patterns were different (Fig. 2B, C). At low or high fragment GC contents, these trimers displayed relatively low Z-scores, with no improvement in Z-scores evident after consideration of GC content. However, at GC contents between 35% and 65%, there was a large increase in Z-scores when GC contents were considered, with mean Z-scores as high as 5.9 and 3.5 for GCG and GAG, respectively – as much as three-fold higher than obtained without consideration of GC content. As with the TGT trimer, this translated to an improved distinction between samples from cancer patients and healthy individuals (Mann-Whitney p-values of 8.3e-14 *vs*. 5.7e-5 with and without GC content consideration, respectively, for GCG; 3.3e-14 with GC content *vs*. 3.3e-8 with and without GC content consideration, respectively, for GAG, Supplementary Table 2).

Fig 2D illustrates a trimer (ATT) in which there is relatively little change in the Z-scores as a function of GC content of the fragment. However, there were still cancer-specific differences in the frequencies of ATT end-motifs, with a max z-score of 1.55 with GC content consideration and mean Z-score of 0.77 without GC content consideration (Mann-Whitney p-values of 5.4e-05 and 0.006 with and without GC content consideration, respectively; Supplementary Table 2).

#### A classifier

Based on the two observations described above, it was clear that a subset of trimers exhibited GC-dependent, cancer-associated differences in frequencies. To incorporate the relationship between GC content and trimer frequencies into a classifier, we performed feature selection on the basis of their mutual information and Mann-Whitney p-values using leave-one-out cross validation. We thereby selected an average of 2293 features from the set of 3840 possible features (64 trimers x 60 GC intervals; Table 2 and Methods). These 2293 features included all 64 trimers and an average of 36 GC contents per trimer. To avoid information leakage, all feature selections were performed within each fold of cross-validation. This heuristic was named MendSeqS, for <u>M</u>odified <u>end</u>-based <u>Seq</u>uencing <u>S</u>ystem. For comparison, we selected 47 trimers using the same criteria (Mutual Information and Mann Whitney p-values) without considering GC content of the underlying fragments, referring to this conventional heuristic as EndSeqS (Table 3). We then used logistic regression to assign weights (coefficients) to each of the selected features in MendSeqS and EndSeqS. Logistic regression yielded a single score for each heuristic in each patient (Fig. 3A). The most obvious difference between the performance of MendSeqS and EndSeqS was the tighter distribution of scores in MendSeqS (Fig. 3A). This translated to a lower binary cross-entropy in MendSeqS (0.16) than in EndSeqS (0.21) (Methods). MendSeqS also was significantly more accurate than EndSeqS in ROC analysis (p-value of 0.015, Venkatraman’s test^22^, Fig. 3B). This improvement was particularly significant in the high specificity realm most important for earlier detection, where MendSeqS achieved a sensitivity of 95% (41/43) at 98% specificity whereas EndSeqS achieved a sensitivity of only 84% (36/43) (p-value of 0.024, Pepe’s test^23^, Fig. 3C).

**Figure 3.**
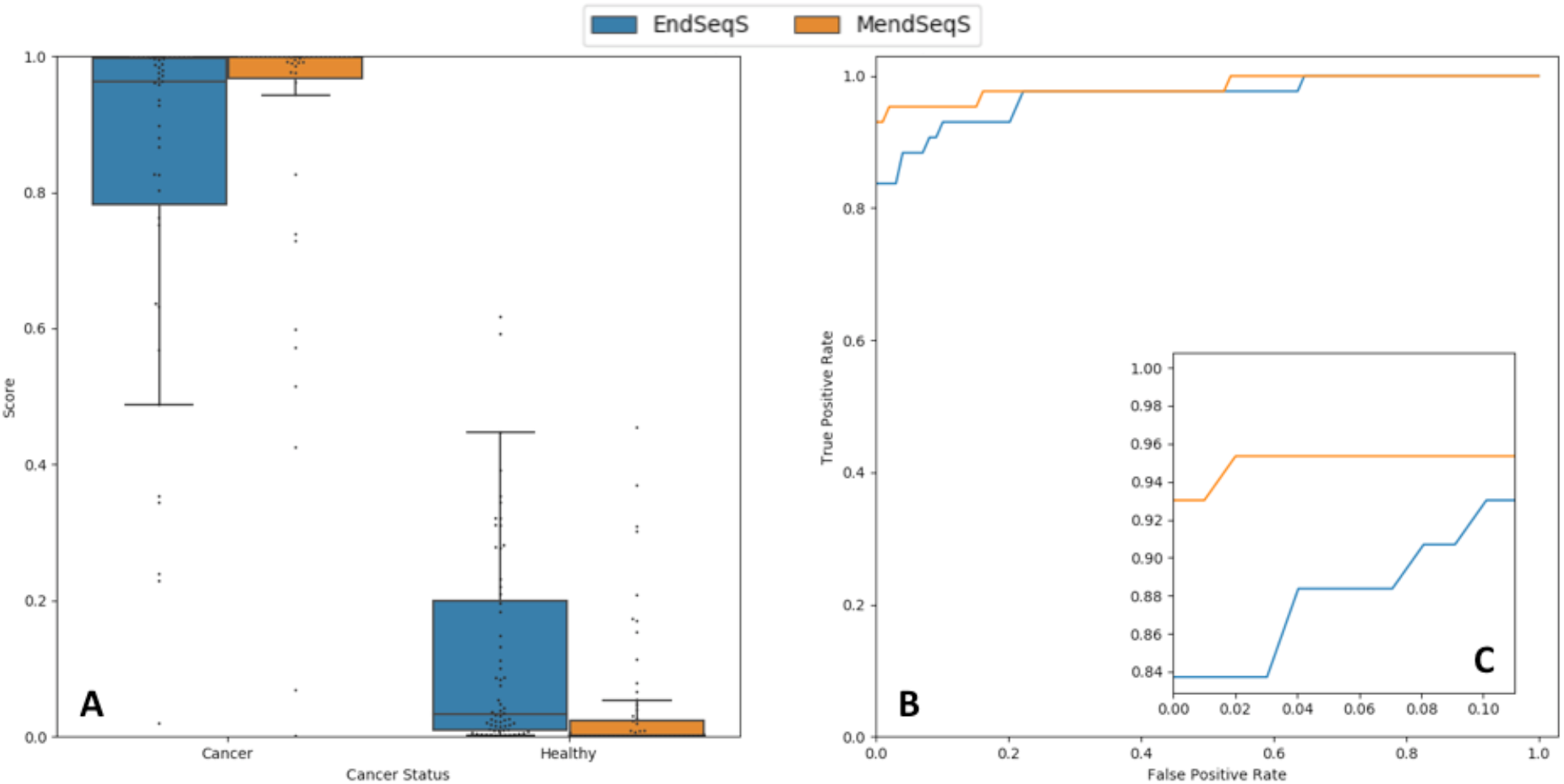
Performance of MendSeqS and EndSeqS on Cohort 1. (A) Box and swarm plots display the scores for MendSeqS and EndSeqS from leave one-out cross-validation. (B) Comparison of ROC curves from MendSeqS and EndSeqS during cross-validation (C) ROC at high specificity, representing the upper left part of (B).

#### Analysis of other WGS datasets

We then sought to see if MendSeqS could be applied to DNA samples that had been prepared, amplified, and sequenced in other laboratories. For this purpose, we employed 401 samples (193 with cancer, 208 without cancer) deposited in the FinaleDB public database^18^ (called Cohort 2,Table 4) from three separate studies, each using different technologies^9, 11, 24^. The cancer types represented in Cohort 2 were different than those in Cohort 1 (compare Table 1 with Table 4).

The first question addressed was whether the two basic observations that formed the rationale for MendSeqS were apparent in Cohort 2. With respect to Observation 1, the data in Figures 1E-H show a strong GC-dependence of the frequencies of the same four trimers illustrated in Figs 1A-D, and this was true for all 64 trimers (Supplementary Data 1).

With respect to Observation 2, the key question was whether MendSeqS could improve the classification of cancer samples in Cohort 2 over that achieved with EndSeqS. Because the sample types and methods used for DNA purification, library preparation, and sequencing analysis were heterogeneous in Cohort 2 and different than in Cohort 1, we derived new features and coefficients for Cohort 2 and evaluated them using leave one-out cross-validation (just as done for Cohort 1). For MendSeqs in Cohort 2, we selected an average of 2414 features (64 trimers x average of 38 GC contents per primer) from the set of 3840 possible features (Table 5). For EndSeqS, we selected all 51 using the same criteria (Table 6, Methods).

Dot plots of the scores of the four cancer types obtained with MendSeqS and EndSeqS are plotted in Fig. 4A, where the tighter distribution of MendSeqS scores was again observed. MendSeqS also was more accurate than EndSeqS in ROC analysis (p-value of 0.054, Venkatraman’s test^22^, Fig. 4B). This improvement was particularly significant in the high specificity realm most important for earlier detection, where MendSeqS achieved a sensitivity of 87% (168/193) at 98% specificity, whereas EndSeqS was less sensitive at the same specificity (68% [131/193], p-value of 0.0087, Pepe’s test^23^, Fig. 4C).

**Figure 4.**
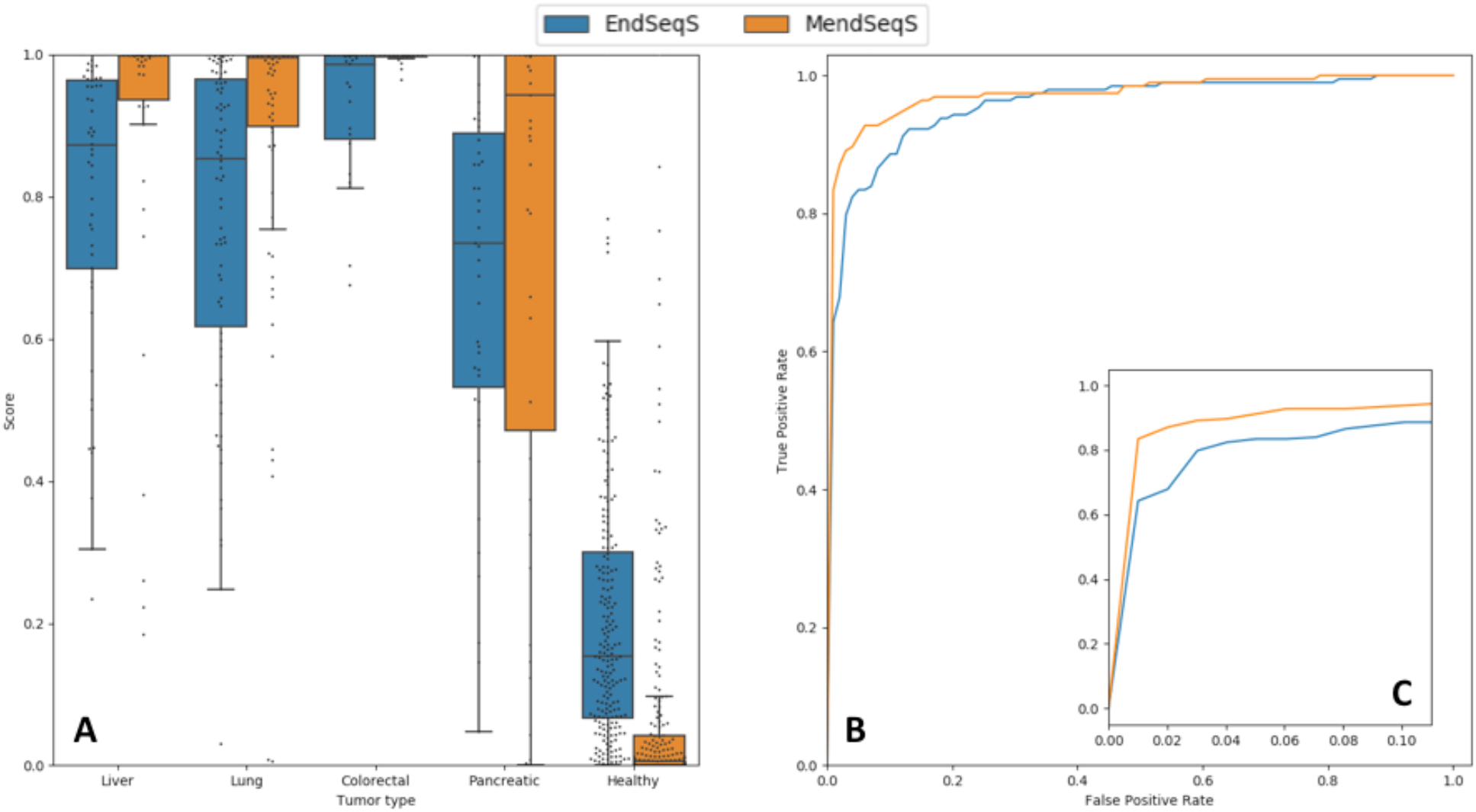
Performance of MendSeqS and EndSeqS on Cohort 2. (A) Scores from leave one-out cross-validation for four tumor types and healthy samples are shown.. (B) ROC curves displaying the performance across the 10 iterations of cross-validation.(C) ROC at high specificity, taken from the upper left part of (B).

## Discussion

The results described above demonstrate that the distributions of trimers at the 5’ ends of cfDNA fragments are influenced by the GC content of the fragment from which those ends were derived. Moreover, taking this GC dependency into account can magnify the differences in trimer frequency between cancer patients and healthy individuals, resulting in improved classification performance.

The GC dependence of trimers was observed in samples from both cancer patients and healthy controls. The similarity of the trends as seen in cell-free DNA and *in silico* fragmentation experiments indicate that these trends reflect the influence of local GC content on the frequency of each end-motif (Fig. 1I-L; Methods). Moreover, while the overall shapes of the curves relating to end-motif frequency and fragment GC content are observed in experimental data (Cohorts 1 and 3), and in silico fragmentation, their magnitude is different. Note, for example, that the y-axes are different in Fig 1A, 1E, and 1I. Previous studies have shown that the distribution of fragment-end motifs are influenced by the activity of sequence specific nucleases such as DNASE1, DNASE1L3, and DFFB^25-26^ as well as chromatin organization, with DNA wrapped around nucleosomes less susceptible to cleavage than internucleosomal regions^11, 27^. Our results illustrate that the frequency of end-motifs is also heavily influenced by GC content of the region itself, implying that phenomena such as PCR biases and copy number changes may add significant noise to the overall end-motif frequency. Through the binning of end-motifs by GC content, it is possible to disaggregate the signal derived from fragments of varying GC content, allowing MendSeqS to overcome these confounders.

Several prior studies have documented the utility of cancer-specific differences in trimers or other motifs at the ends of cfDNA fragments for cancer detection^13, 17, 19-20^. MendSeqS amplifies these differences, as documented by higher Z-scores, p-values, and mutual information when GC content of fragments are considered (Supplementary Table 2). There are at least three possible explanations for these cancer-specific differences. The activities of sequence-specific nucleases that produce cfDNA fragments in neoplastic cells may not be identical to those in non-neoplastic cells. For example, DFFB, associated with certain forms of cell death, could be more active in a subset of neoplastic cells^3, 25-26, 28^. Second, DNA from neoplastic cells could be more fragmented as a result of unrepaired DNA damage by exogenous or endogenous sources of DNA damage (e.g. chemical insults, radiation, free radicals, topological changes) which may lead to changes in the distribution of end-motifs. ^8, 13, 20^. Third, differences in chromatin structure between neoplastic and non-neoplastic cells are known to alter the fragmentation patterns of cell-free DNA and could explain both cell-type specific and cancer-specific changes ^7-8, 11, 14, 29-31^. Further research will be required to determine the relative importance of these three potential explanations.

Note that in the proposed explanations above we are *assuming* that the cancer-specific differences in trimers are the result of contributions of cfDNA derived from neoplastic cells. In fact, neither our data nor prior evidence provide unequivocal evidence that the fragments whose ends give rise to the cancer-specific signal are actually derived from cancer cells. They could be derived from non-cancer cells of the organ giving rise to the cancer, or from leukocytes or other cells that are simply associated with cancer.

Our study of course has limitations. Among them, our cohorts were relatively small so the confidence limits in the estimates of sensitivity and specificity derived from the ROC curves are relatively wide. Second, in the ideal situation, the selected features and the logistic regression coefficients associated with each feature should be broadly applicable to all cohorts. However, when we applied the features and coefficients derived from the analysis of data generated in other laboratories (Cohort 2) to those generated in our laboratory (Cohort 1), the MendSeqS sensitivity at 95% specificity dropped from 92% to 23.1%. This drop in sensitivity was also observed in EndSeqS, dropping from 89% to 18.1% when features from Cohort 1 rather than Cohort 2 were used. Therefore, the decreased performance was not likely to be a result of the new heuristics employed in MendSeqS. The most obvious basis for the difference in performance is that the sample preparation, DNA library construction, and sequencing methods – factors that are known to influence characteristics of cell-free DNA^32-35^ – used in Cohort 2 were different than those used to in Cohort 1. The enzymes used to make WGS libraries often employ nucleases and polymerases that can alter the bases at the ends of fragments in preparation for ligation, so this explanation is plausible. If true, it would suggest that the parameters for GC adjustment used in a training set should be derived from control libraries made identically to those used in the validation set. However, it is also possible that these performance differences were representative of differences between tumor types in Cohort 2 (Liver, Lung, Pancreas, and Colorectal) compared to Cohort 1 (predominately Colorectal), or to other confounders. Regardless, studies with more patients, and with different types of library constructions on the same patients, should be able to address this issue in the future.

In summary, we demonstrate that the frequency and cancer-specificity of a cfDNA fragment end-motif are correlated with the GC content of the fragment from which the was derived. We anticipate that the principles on which MendSeqS is based can be incorporated into the analysis of cfDNA in general and can serve as an adjunct to other assays performed on the same cfDNA, such as mutations, methylation, fragment length, and chromatin accessibility.

## Methods

### Plasma Collection and DNA Collection

This study was approved by the Institutional Review Boards for Human Research at participating institutions in compliance with the Health Insurance Portability and Accountability Act. All the participants provided written informed consent in accordance with the principles of the Declaration of Helsinki. DNA was purified from an average of 1 to 10 mL of plasma using either a QIASymphony circulating DNA kit (cat # 1091063) or a BioChain Cell-free DNA Extraction kit (Cat # K5011625).

### Library Preparation and Sequencing

All libraries were prepared as described in ref. ^36^. Barcoded libraries were sequenced using 75 bp paired-end runs (150 cycles) on either Illumina HiSeq 4000 or Novaseq 6000 platforms to an average depth of 17 million molecules. Adapters and UMIs were trimmed using cutadapt^37^ and trimmed sequences were aligned to the hg19 genome using bowtie2^38^ in paired-end mode. Reads were filtered for a MAPQ>1 and duplicates were removed using unique molecular identifiers (UMIs). Paired-end reads that did not have proper orientation or did not have paired-end support were removed.

### Fragment-End Analysis

Fragmentation data was collected from filtered SAM files. For each pair of reads the fragment start, end, and strand alignment was determined. Bedtools^39^ was then used to sort fragments and extract the full insert sequence using the hg19 reference genome. GC content for each insert was then calculated using all bases of the aligned sequence.

For every unique molecule the first (5’) and last (3’) nucleotides of the fragment sequence were evaluated and the frequency of each motif at each end was determined. Our library preparation either degrades the 3’ end of the original DNA duplex fragment when there is a 3’-overhang or fills-in a 5’ overhang with the compliment of the 3’ strand. We therefore used the end-motif (trimer) itself when the sequence read aligned to the reference strand of the genome, and the reverse compliment of the trimer when the sequence read aligned with the reverse compliment of the reference strand.

We binned each fragment based on the GC content of the sequence of the entire aligned read; when only part of a read aligned to the reference genome, it was discarded. GC bins extended from 20% to 21% GC, 21% to 22% GC, etc. up to 79 to 80% GC. Thus there were 60 possible bins and 64 possible trimers, for a total of 3840 possible features used for MendSeqS.

### In silico fragmentation of the hg19 genome

We extracted fragments from 74 healthy controls and placed them randomly throughout the hg19 genome using the bedtools random function^39^. For example, if sample A had 2000 fragments of length X we placed 2000 fragments of length X randomly throughout the hg19 genome.

### Analysis of FinaleDB samples

Fragmentation data was downloaded for 352 patients from the FinaleDB database^18^. Data files were downloaded in sorted tsv format and contained fragment-end positioning (chr,start,end) and strand alignment. Fragment-end analysis was performed as described above.

### Classification Algorithm

For each potential feature, we used the data within the training set (all but one of the samples) to create a StandardScaler model (sklearn) to standardize features by removing the mean and scaling to unit variance. Next, we evaluated (1) the mutual information between feature values and cancer status and (2) the Mann-Whitney p-value for feature values of healthy controls *vs*. cancer samples (Tables 2, 3, 5, and 6). To remove uninformative features, we filtered for features that had either (a) mutual information greater than 0.05 or (b) a p-value that remained statistically significant (i.e., < E-5) after Bonferonni correction (α=0.05). Logistic regression was then used to determine the coefficient of each feature that surpassed these thresholds. Finally, these coefficients were used to score the left out sample in each fold of cross-validation. Note that both feature selection and coefficient determination were determined in each fold of cross-validation to avoid data leakage during feature selection^21^. After completing all folds, we calculated the binary cross-entropy, AUROC and sensitivity at 98% specificity. This workflow was identical for both MendSeqS and EndSeqS.

### Code Availability

Scripts for analyzing bed files and evaluating logistic regressions are available under a GNU 3.0 public license at https://github.com/sdcurtis/LudwigCenterBaltimore/tree/main

### Data availability

The sequencing data generated in this study can be obtained from the European Genome–phenome Archive (accession number EGAS00001006418)

## Supporting information

MendSeqS Tables

MendSeqS Supplementary Figures

## Data Availability

The sequencing data generated in this study can be obtained from the European Genome phenome Archive (accession number EGAS00001006418)

https://github.com/sdcurtis/LudwigCenterBaltimore/tree/main

## Conflicts of Interest

BV, KWK, & NP are founders of Thrive Earlier Detection, an Exact Sciences Company. KWK, NP, & CD are consultants to Thrive Earlier Detection. BV, KWK, NP, SZ, and CD hold equity in Exact Sciences. BV, KWK, NP, and SZ, are founders of or consultants to and own equity in ManaT Bio., Haystack Oncology, Neophore, CAGE Pharma and Personal Genome Diagnostics. NP is consultant to Vidium. PG and JT are consultants to Haystack Oncology. BV is a consultant to and holds equity in Catalio Capital Management SZ has a research agreement with BioMed Valley Discoveries, Inc. CB is a consultant to Depuy-Synthes, Bionaut Labs, Haystack Oncology and Galectin Therapeutics. CB is a co-founder of OrisDx. The companies named above, as well as other companies, have licensed previously described technologies related to the work described in this paper from Johns Hopkins University. BV, KWK, NP, JDC, and CD are inventors on some of these technologies. Licenses to these technologies are or will be associated with equity or royalty payments to the inventors as well as to Johns Hopkins University. Patent applications on the work described in this paper may be filed by Johns Hopkins University. The terms of all these arrangements are being managed by Johns Hopkins University in accordance with its conflict of interest policies.

## Funding

This work was supported by The Lustgarten Foundation for Pancreatic Cancer Research, The Virginia and D.K. Ludwig Fund for Cancer Research, The Conrad N. Hilton Foundation, The Sol Goldman Charitable Foundation, and National Institutes of Health grants (T32 GM008752, U01 CA200469, U01 CA62924, T32 GM136577, U01 CA06973, T32 GM135083, and T32 GM007814).

## Notes

### Author Declarations

Approval was granted by the institutional Review Board of the Johns Hopkins Medical Institutions.

