## Supplementary material for "identifying cancer patients from GC-patterned fragment ends of cell-free DNA": MendSeqS Supplementary Figures

### Slide 1
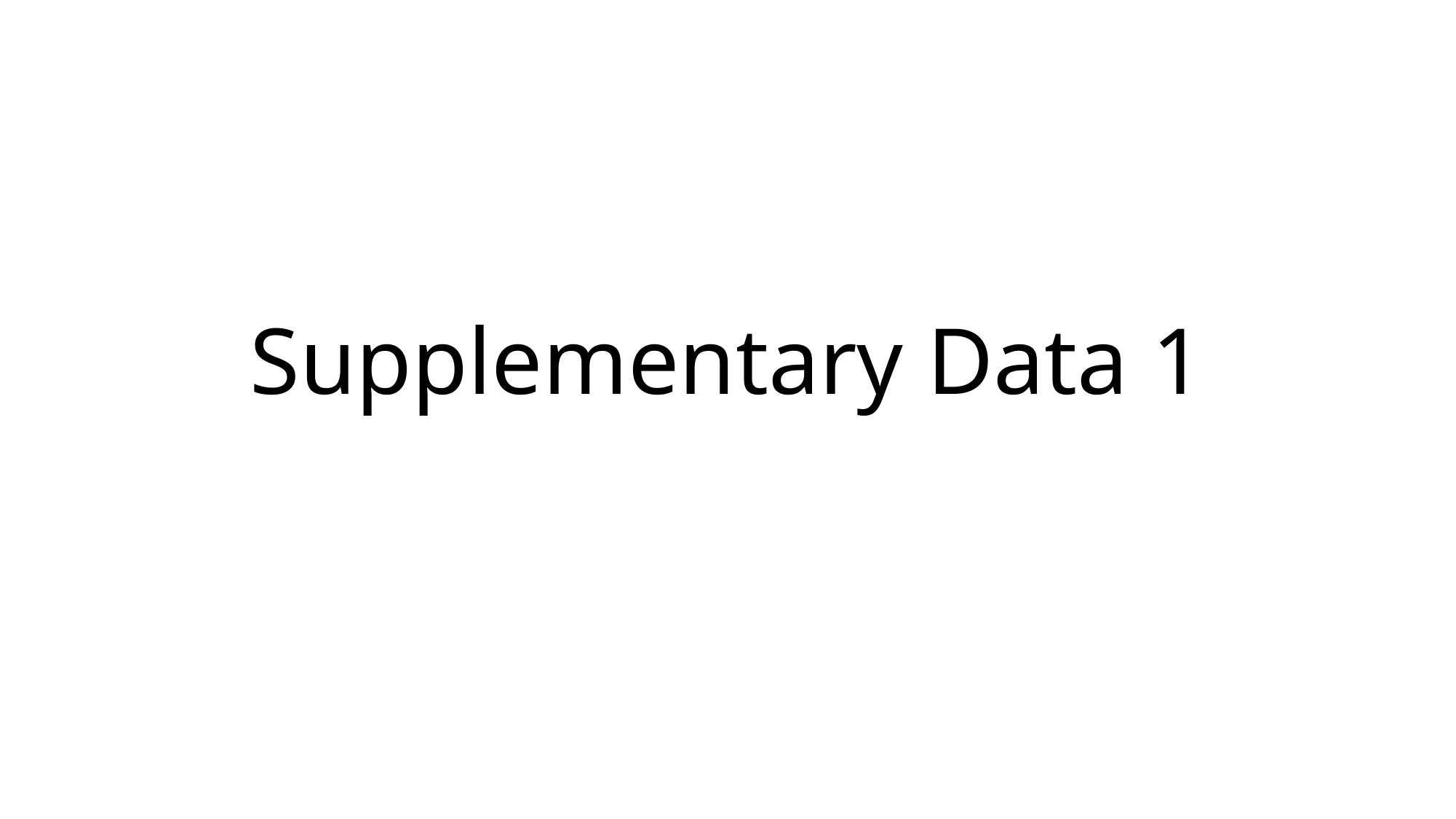

#### Supplementary Data 1

### Slide 2
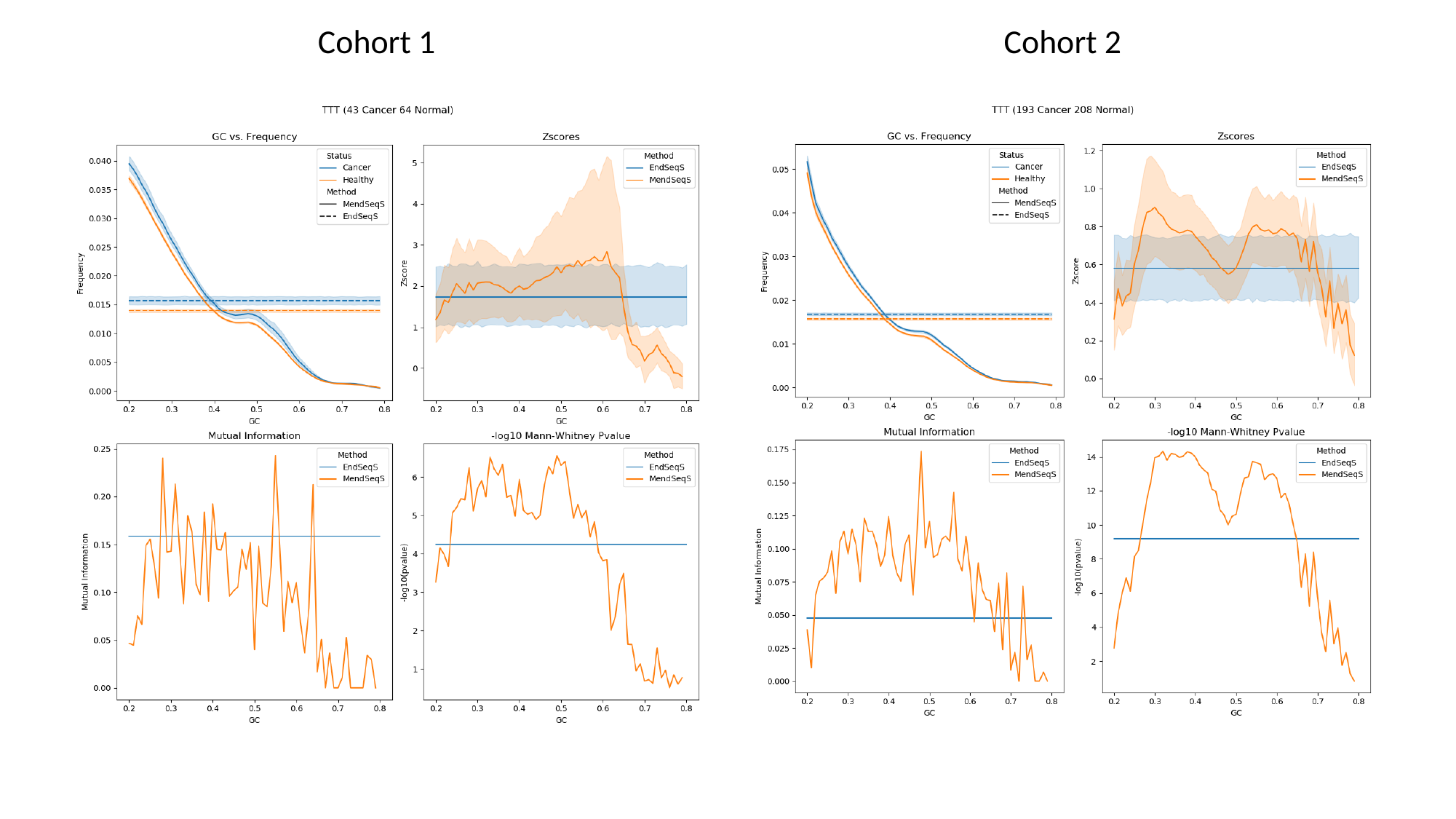

Cohort 2
Cohort 1

### Slide 3
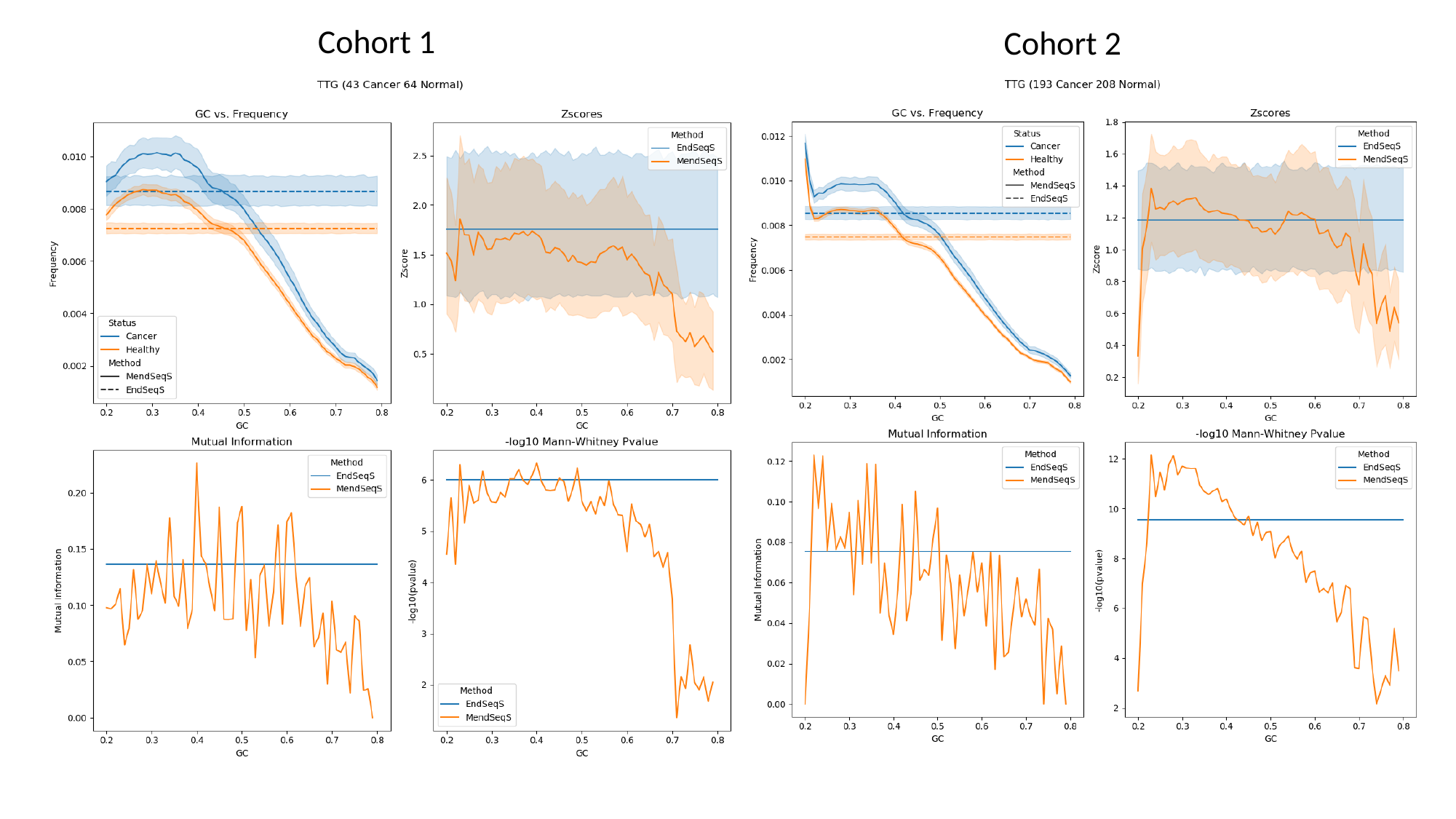

Cohort 1
Cohort 2

### Slide 4
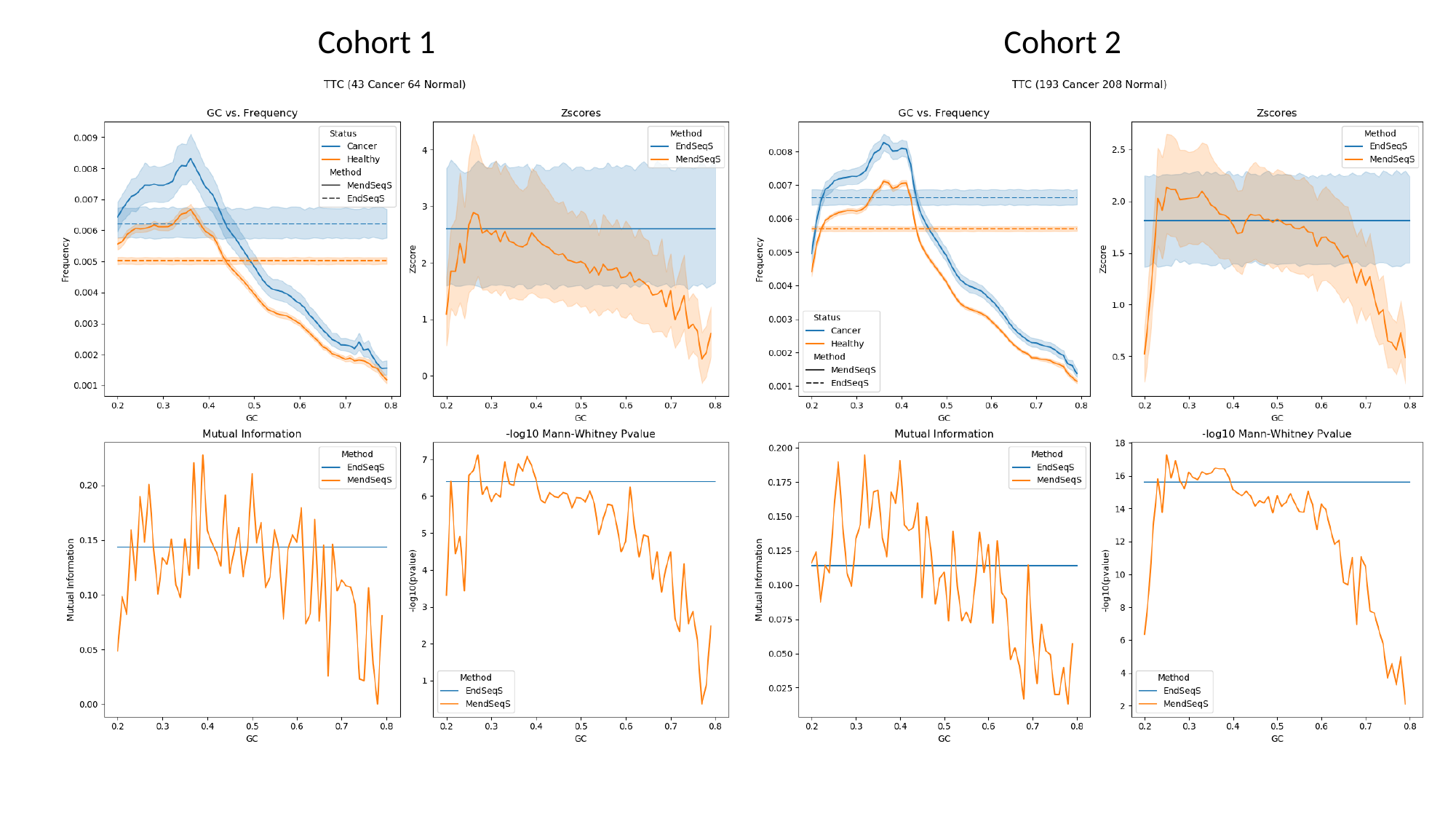

Cohort 2
Cohort 1

### Slide 5
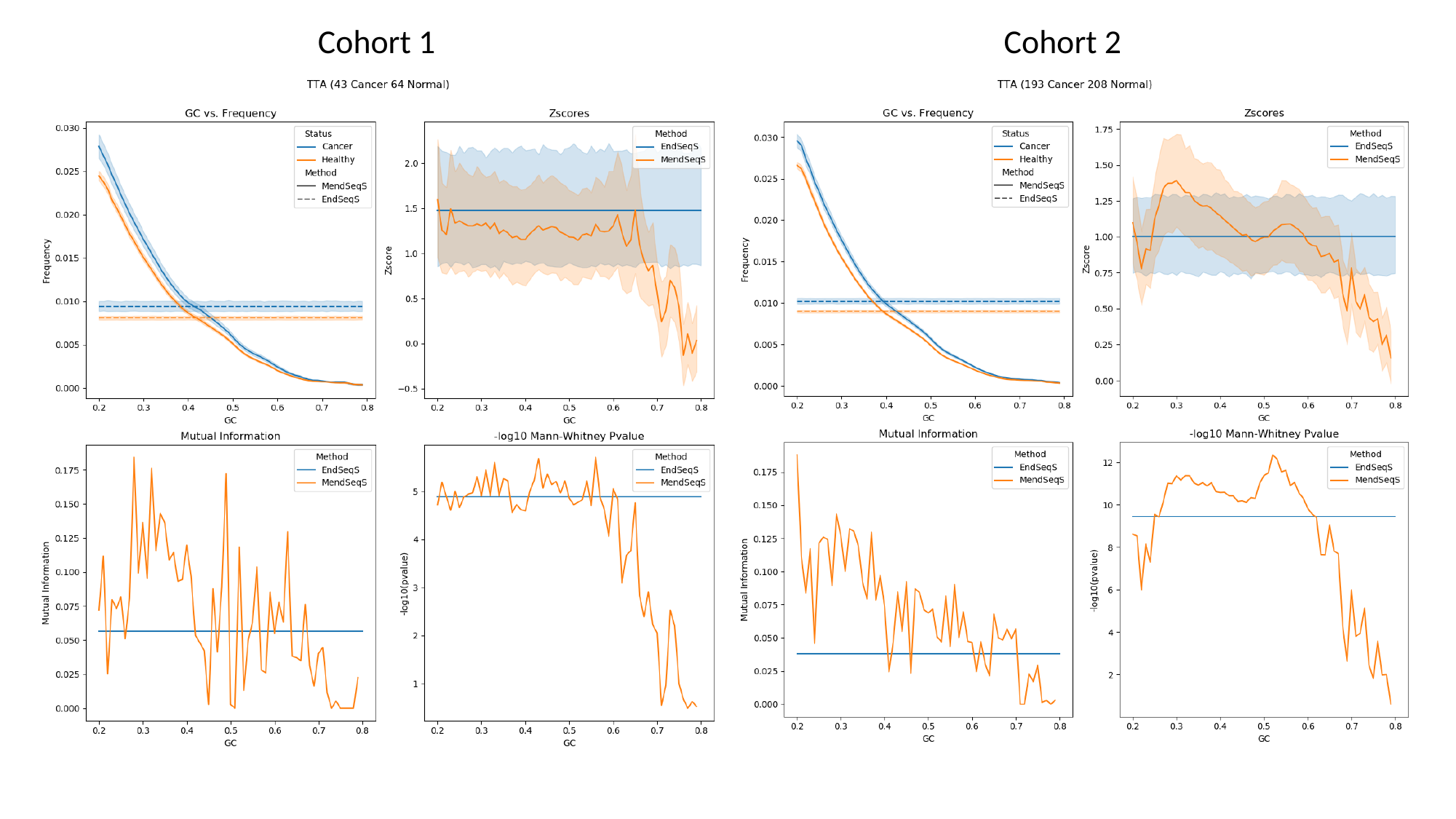

Cohort 2
Cohort 1

### Slide 6
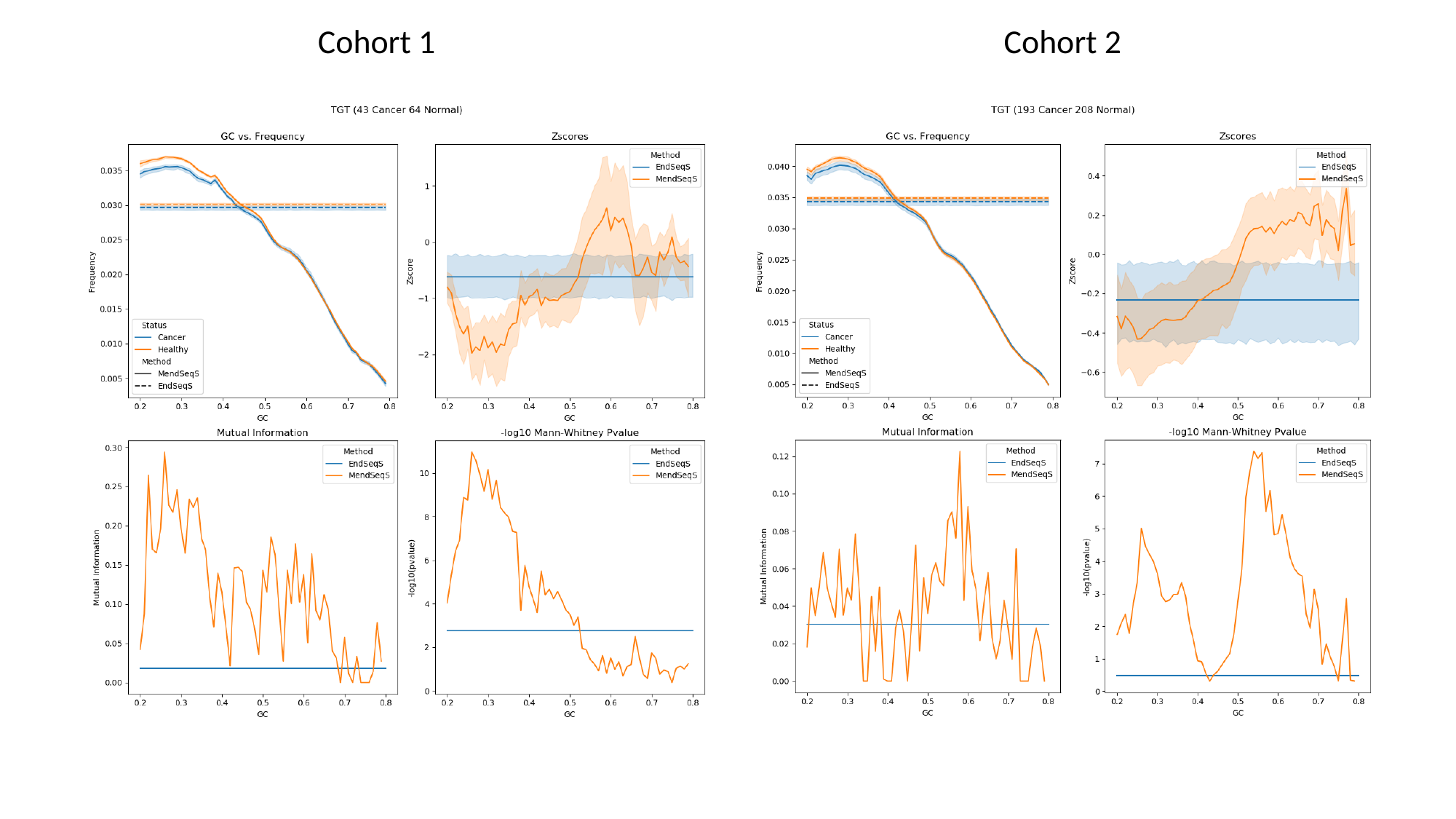

Cohort 2
Cohort 1

### Slide 7
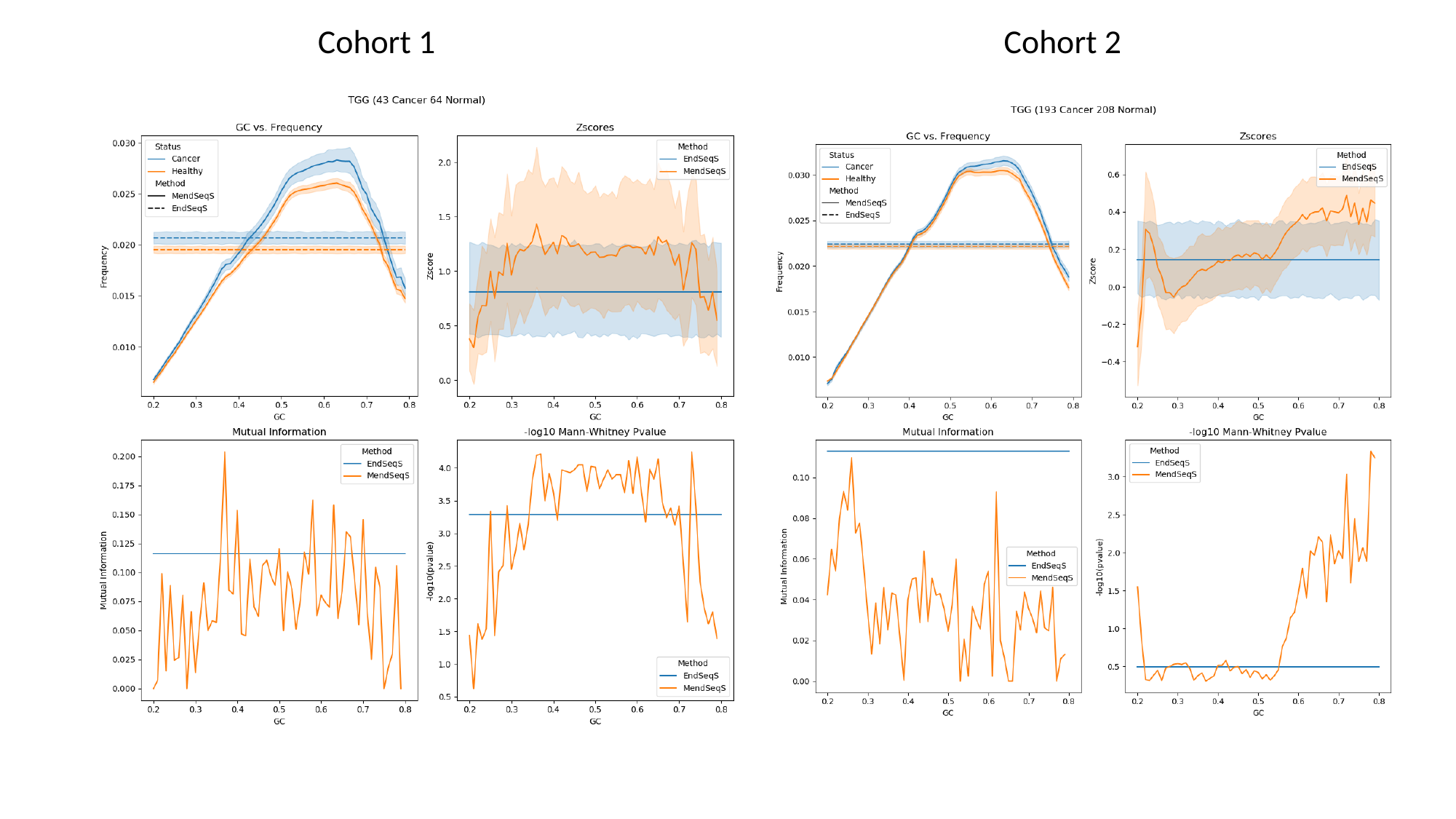

Cohort 2
Cohort 1

### Slide 8
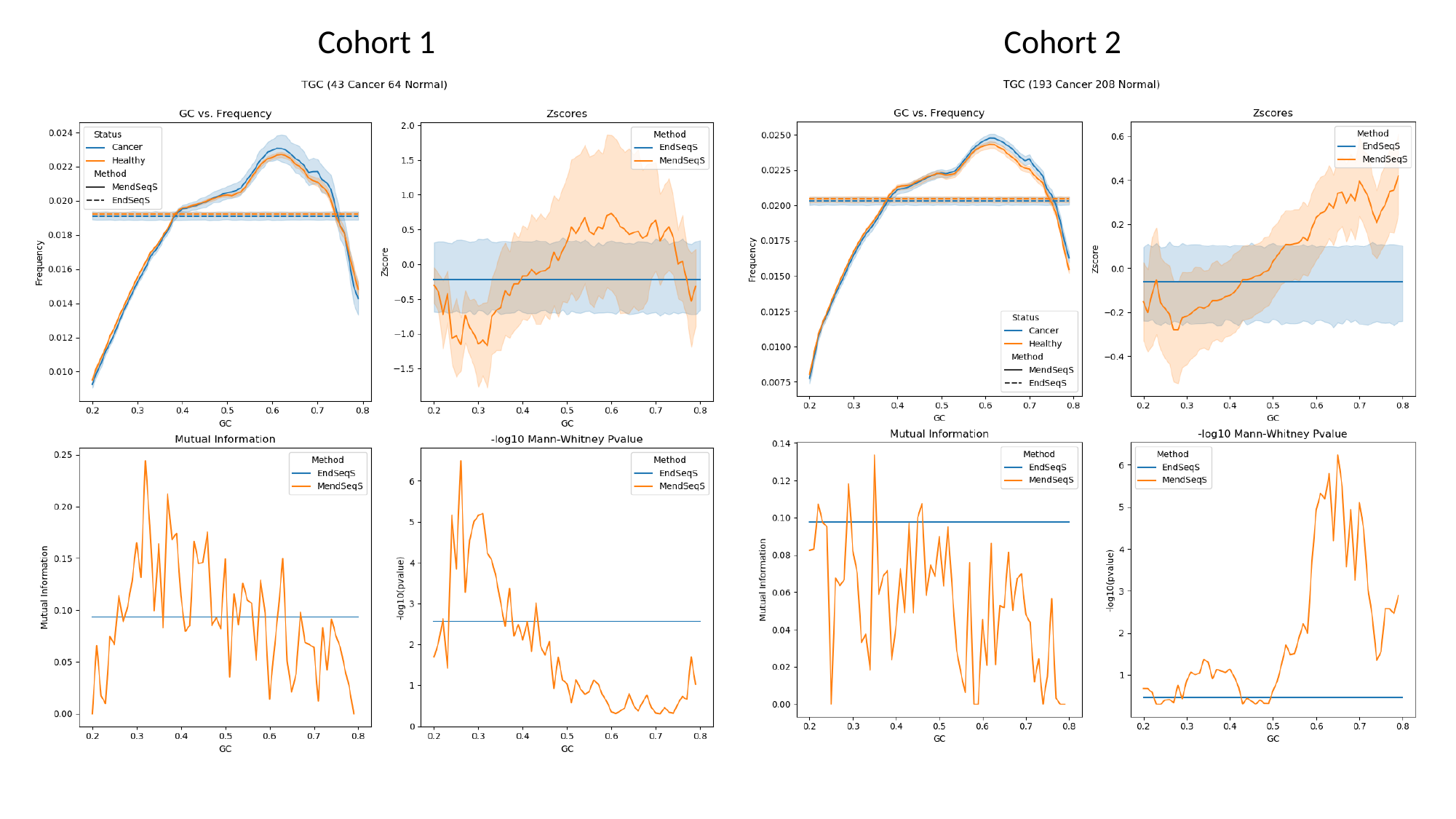

Cohort 2
Cohort 1

### Slide 9
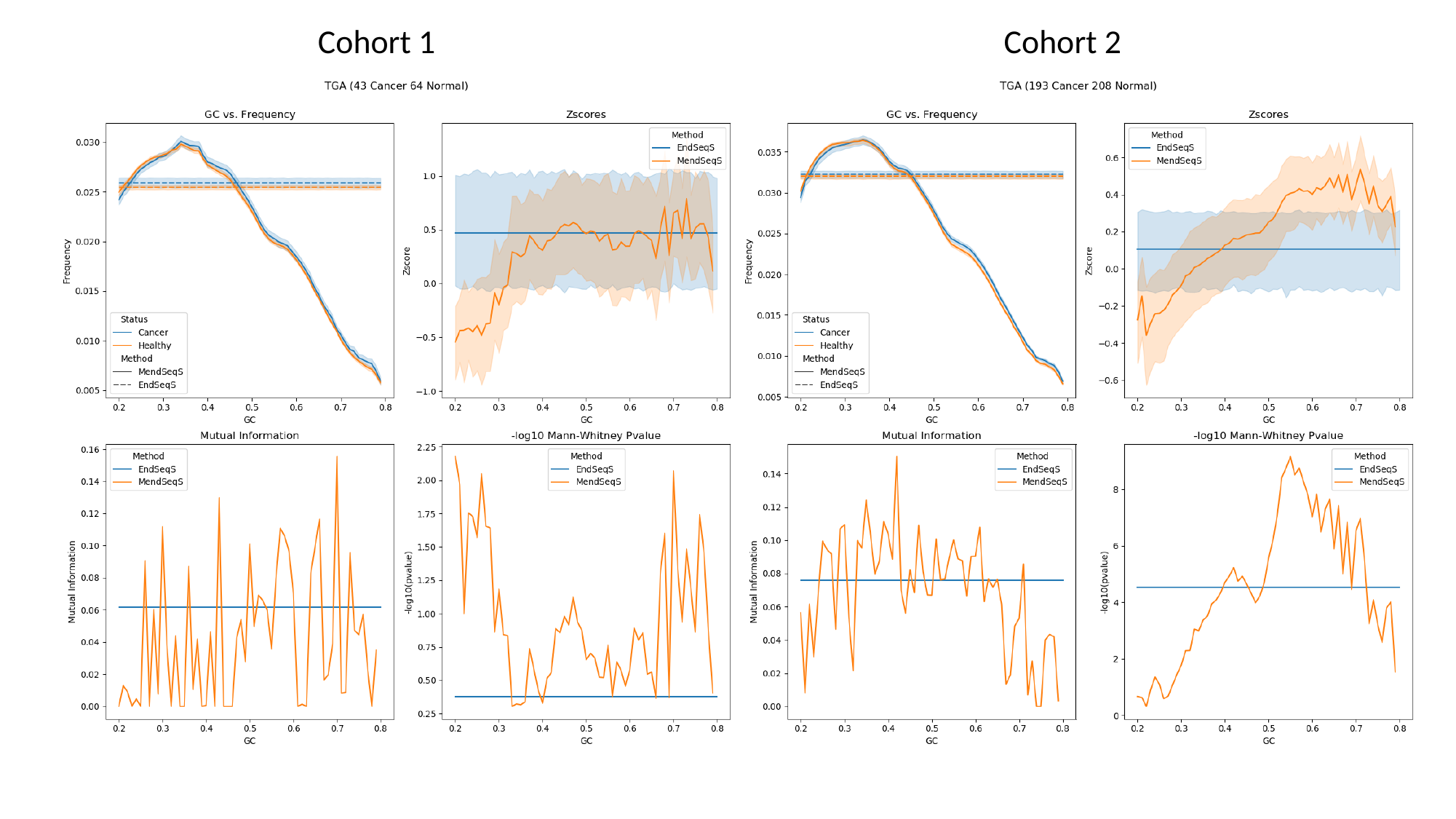

Cohort 2
Cohort 1

### Slide 10
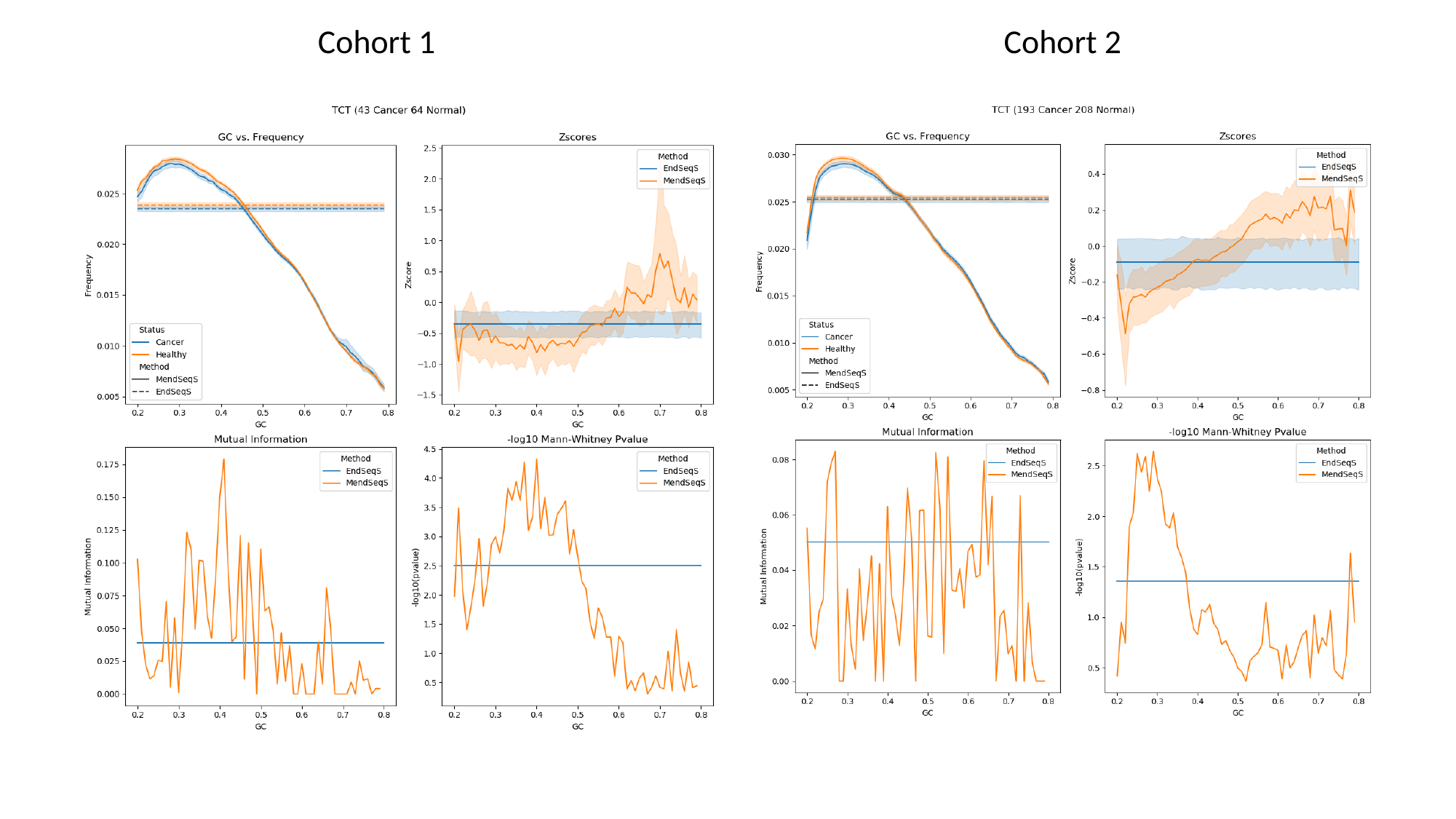

Cohort 2
Cohort 1

### Slide 11
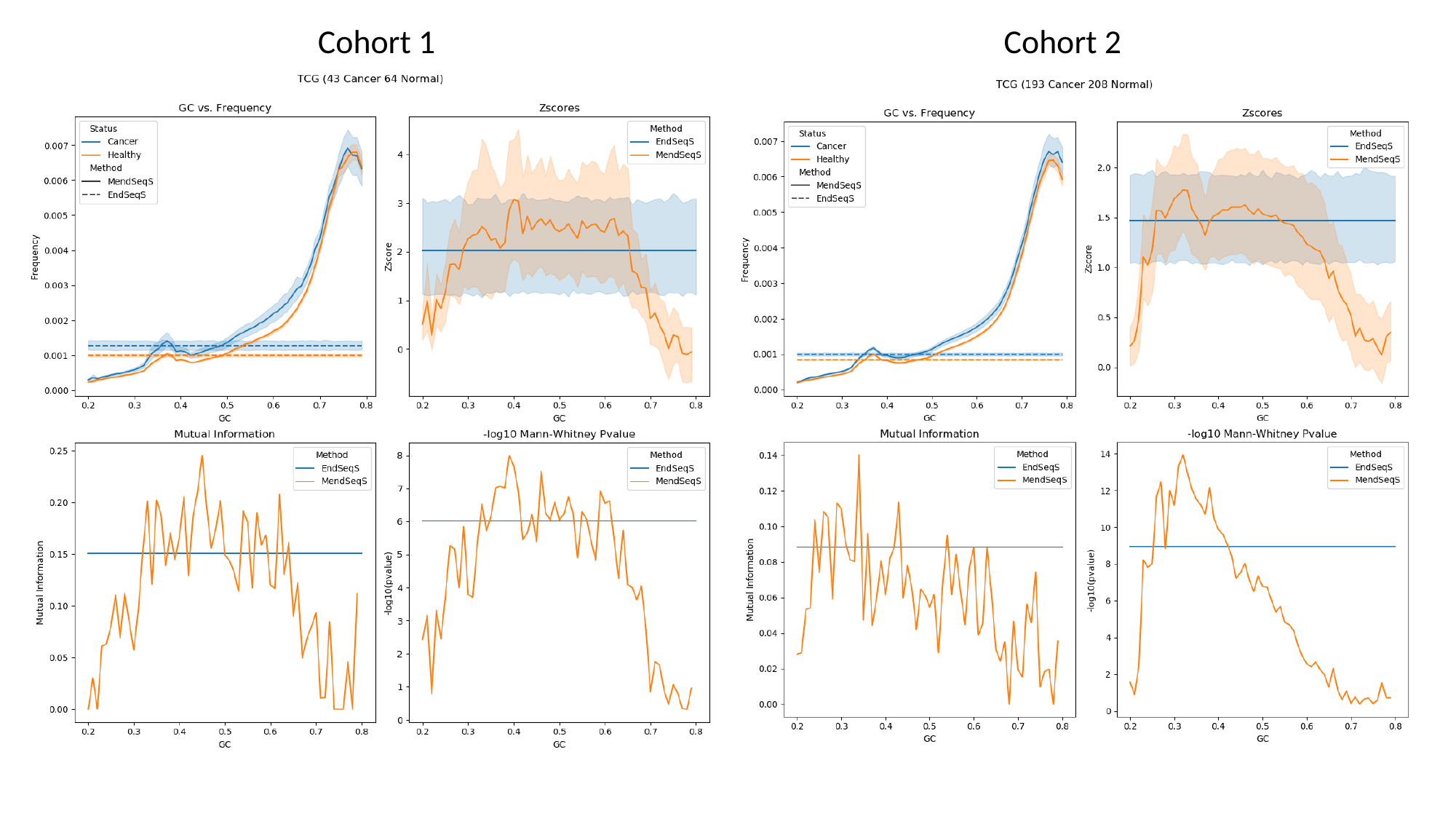

Cohort 2
Cohort 1

### Slide 12
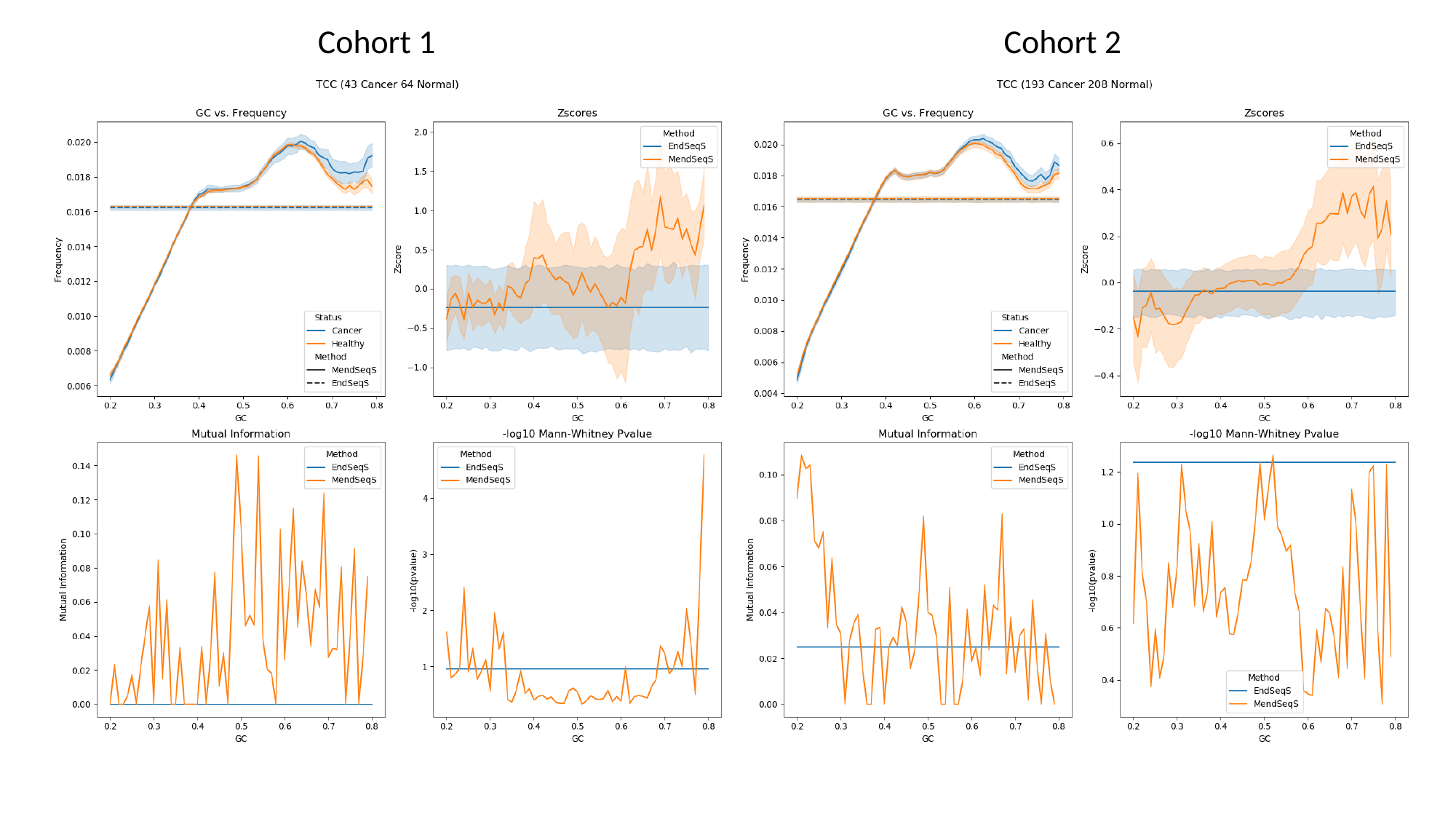

Cohort 2
Cohort 1

### Slide 13
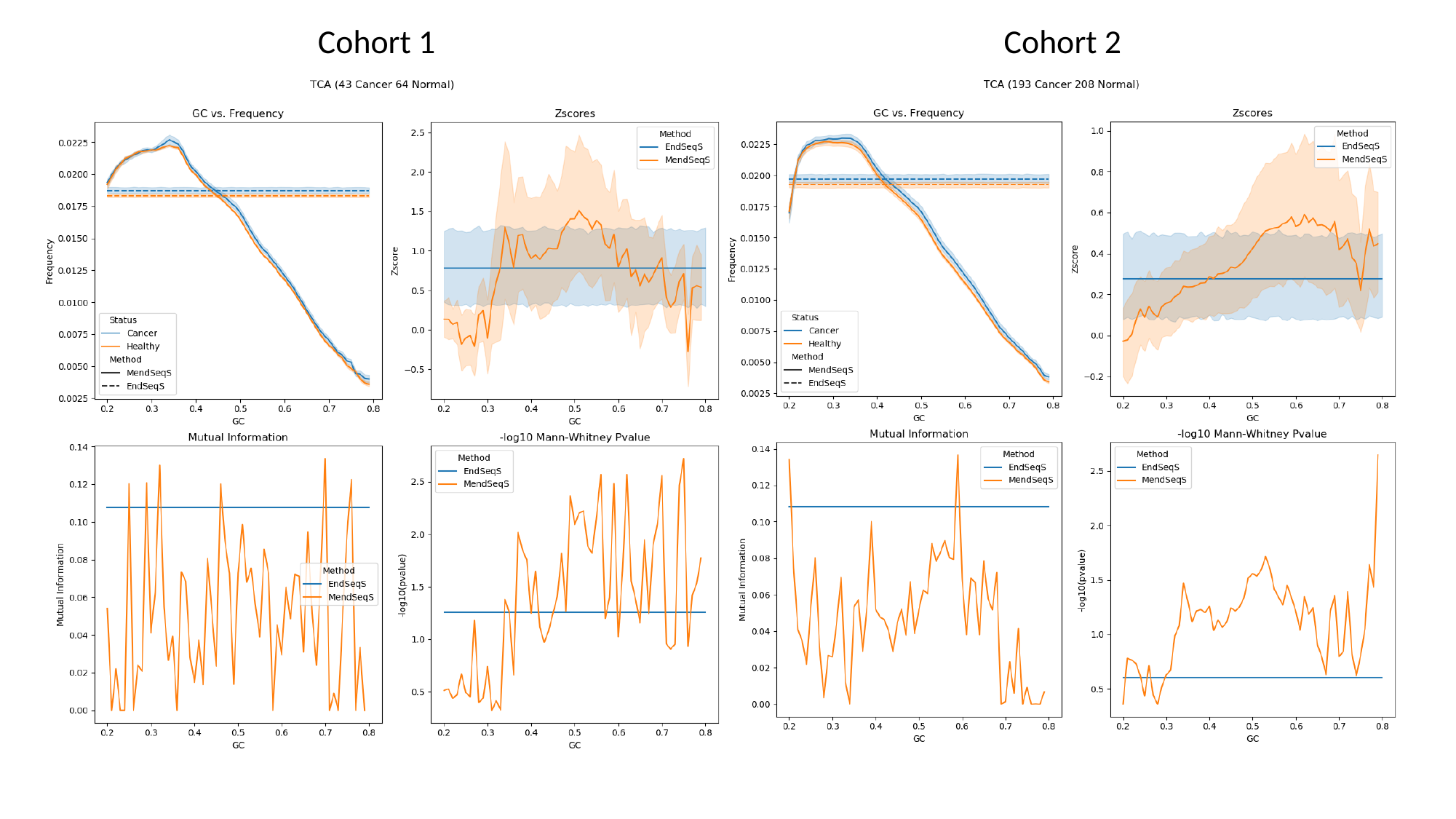

Cohort 2
Cohort 1

### Slide 14
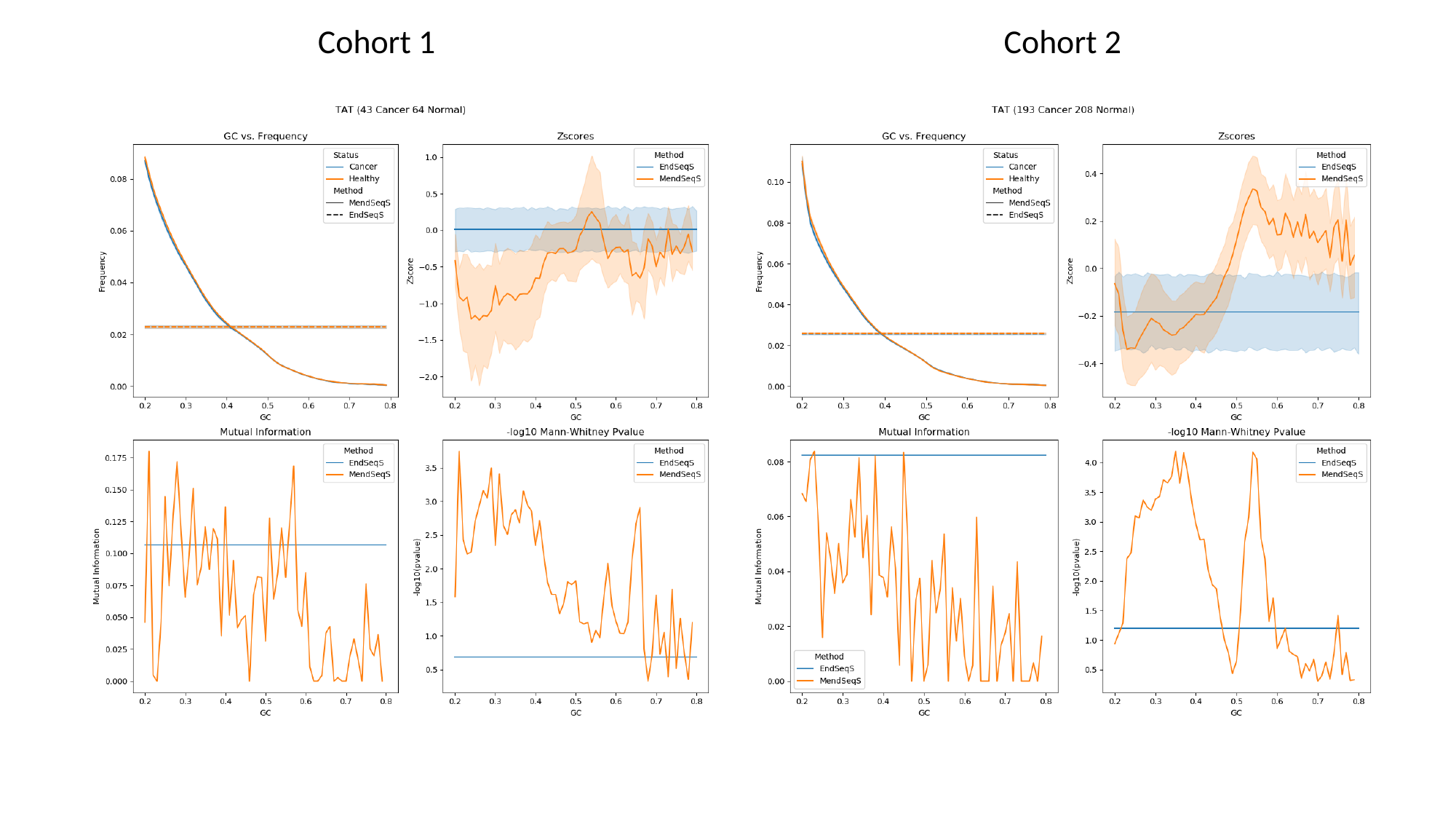

Cohort 2
Cohort 1

### Slide 15
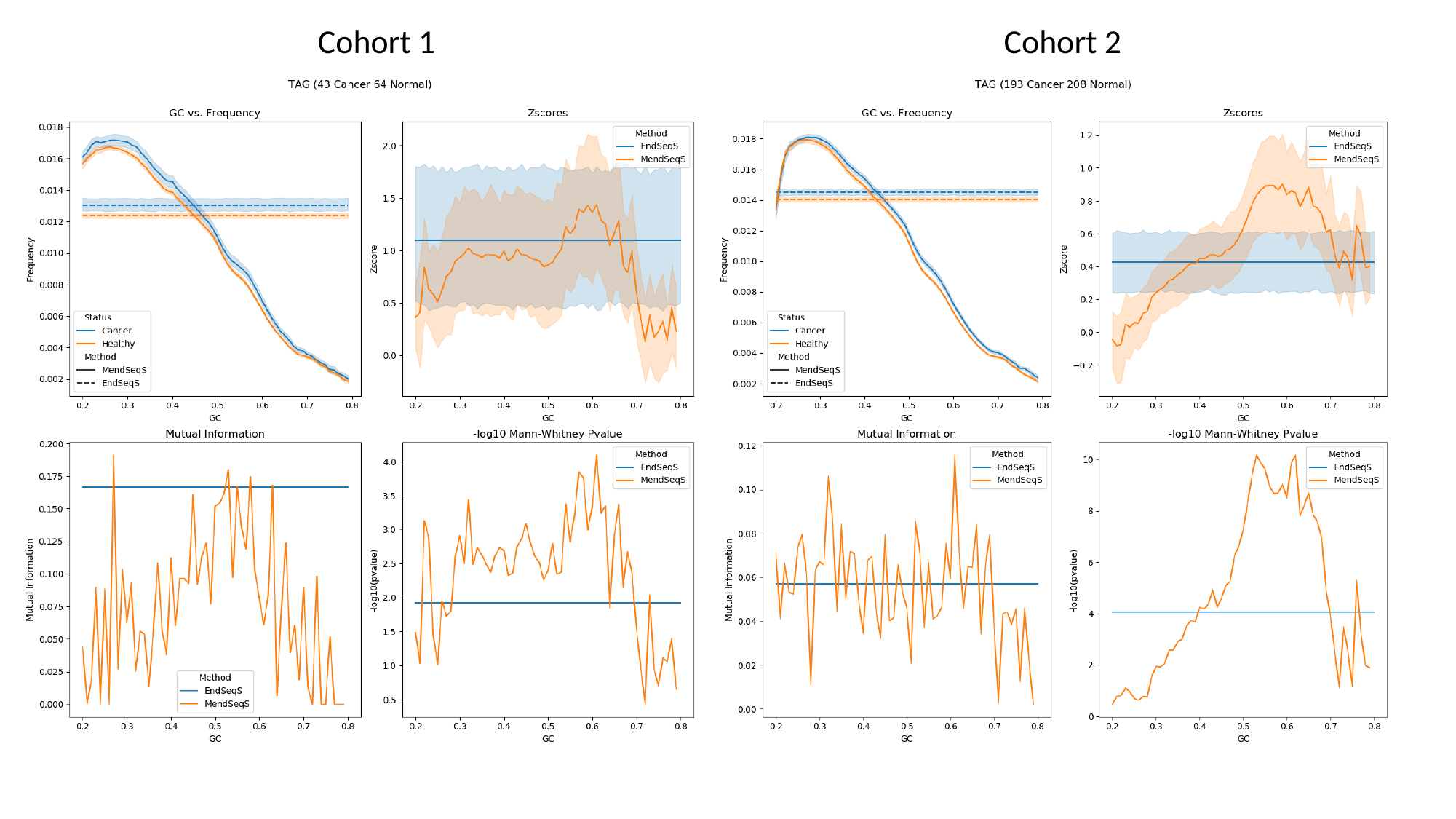

Cohort 2
Cohort 1

### Slide 16
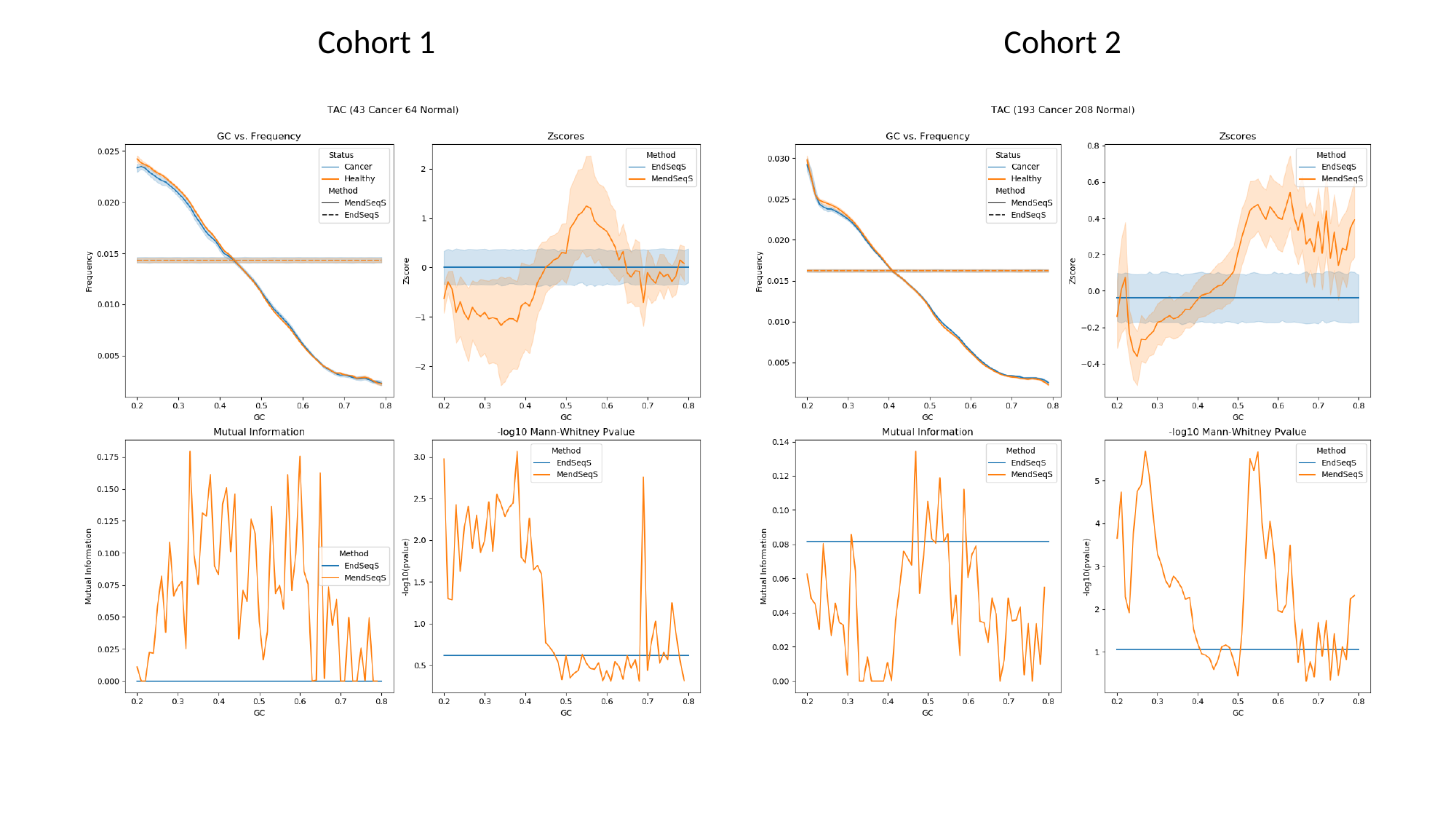

Cohort 2
Cohort 1

### Slide 17
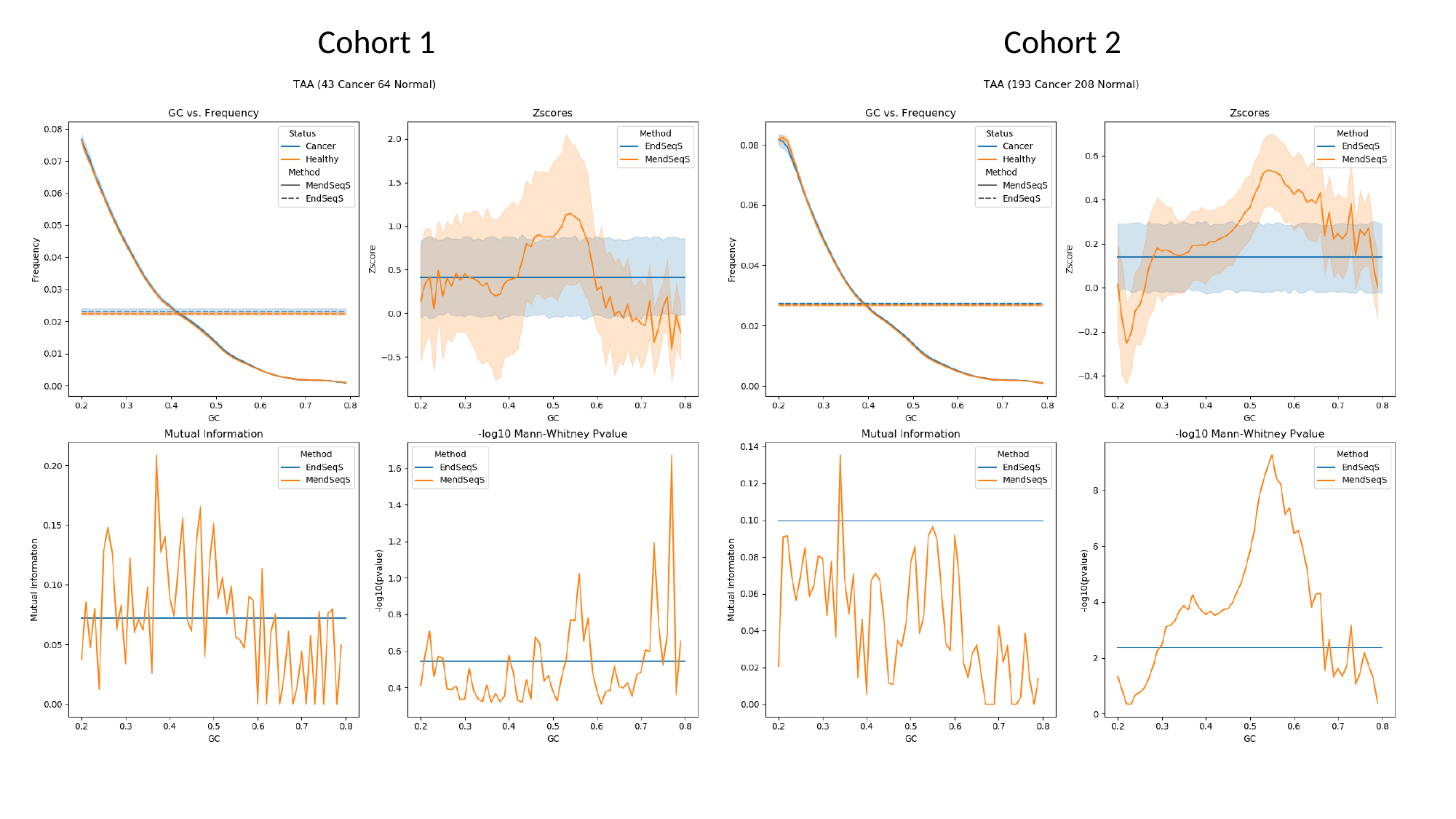

Cohort 2
Cohort 1

### Slide 18
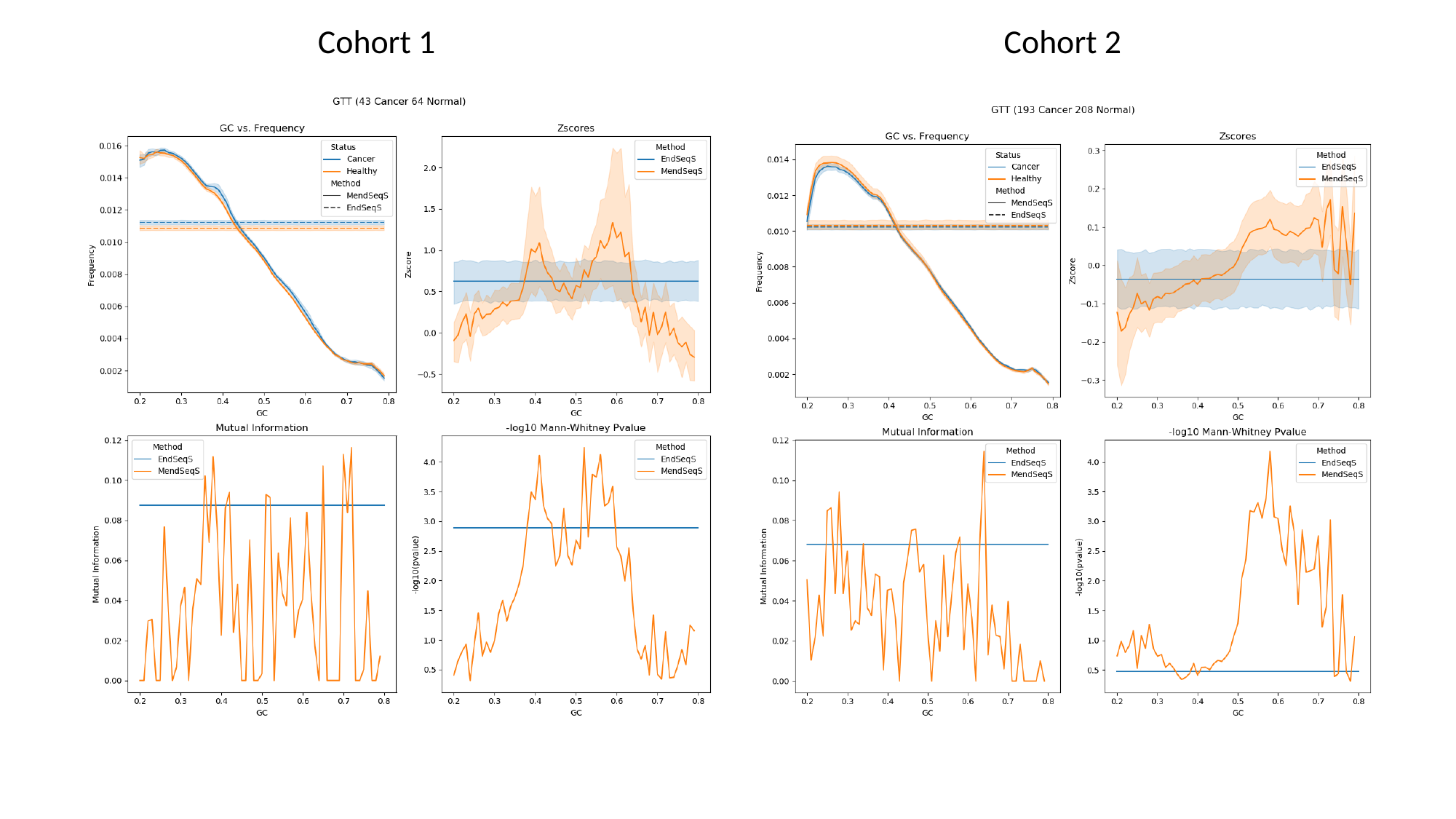

Cohort 2
Cohort 1

### Slide 19
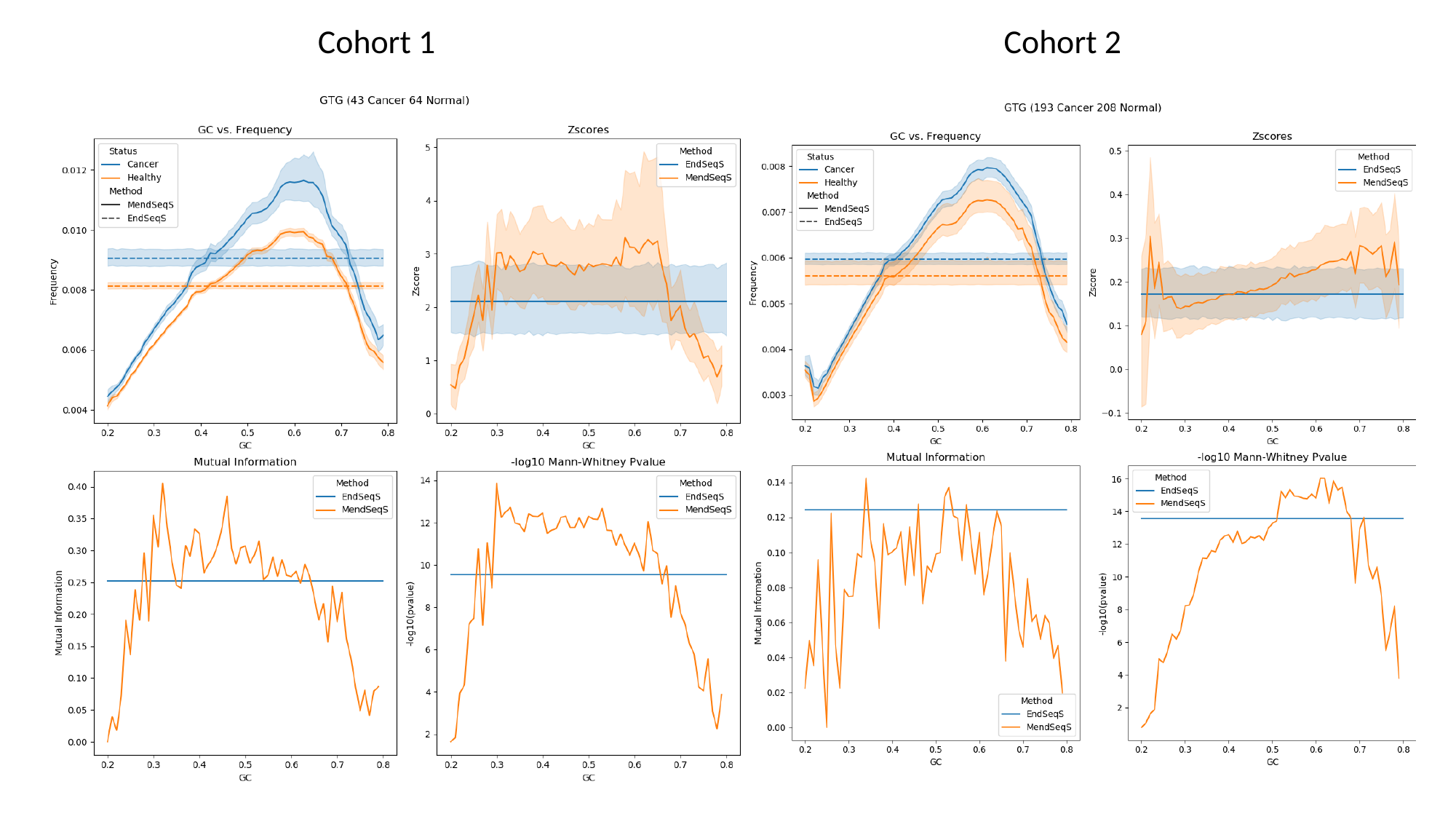

Cohort 2
Cohort 1

### Slide 20
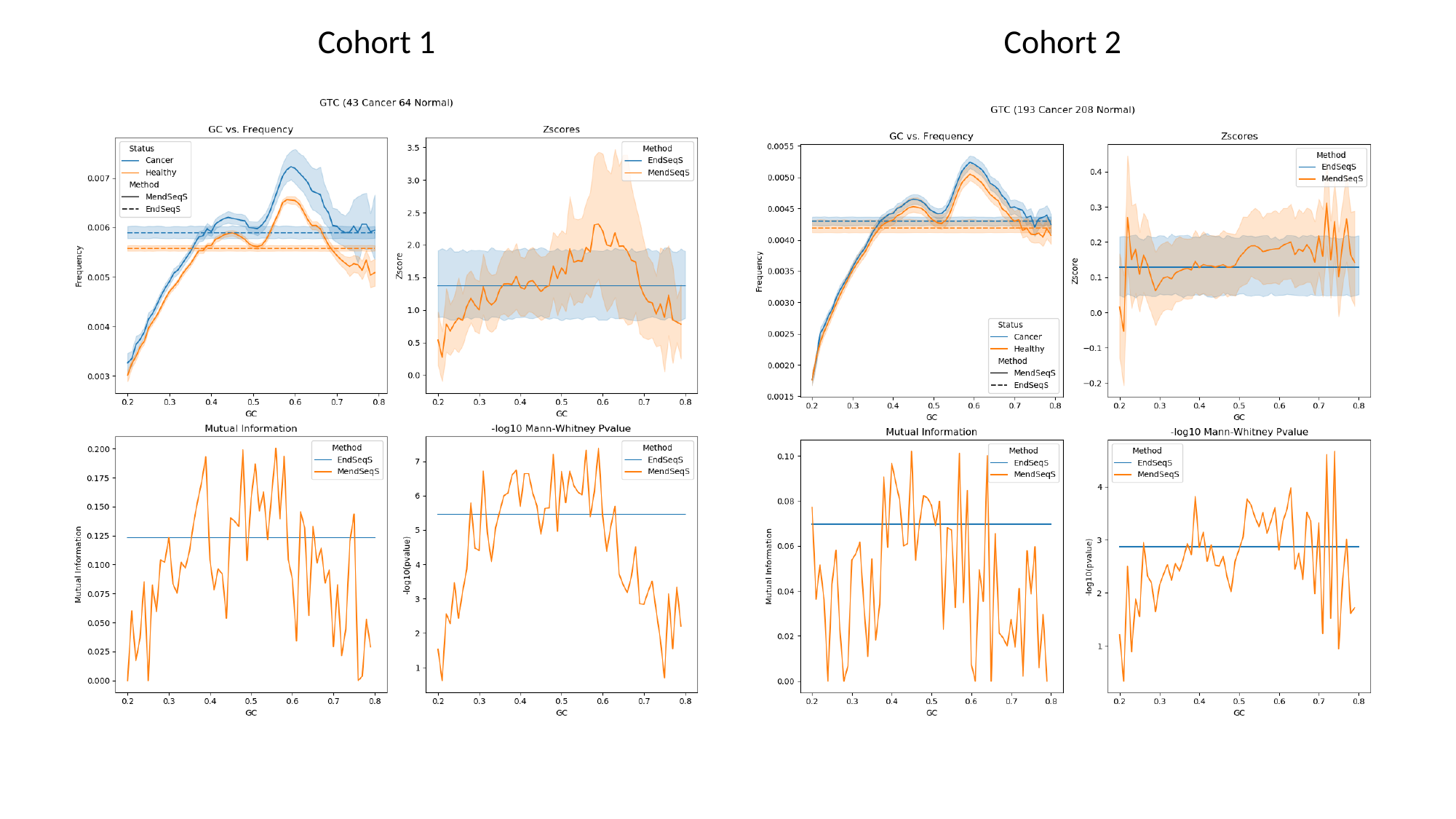

Cohort 2
Cohort 1

### Slide 21
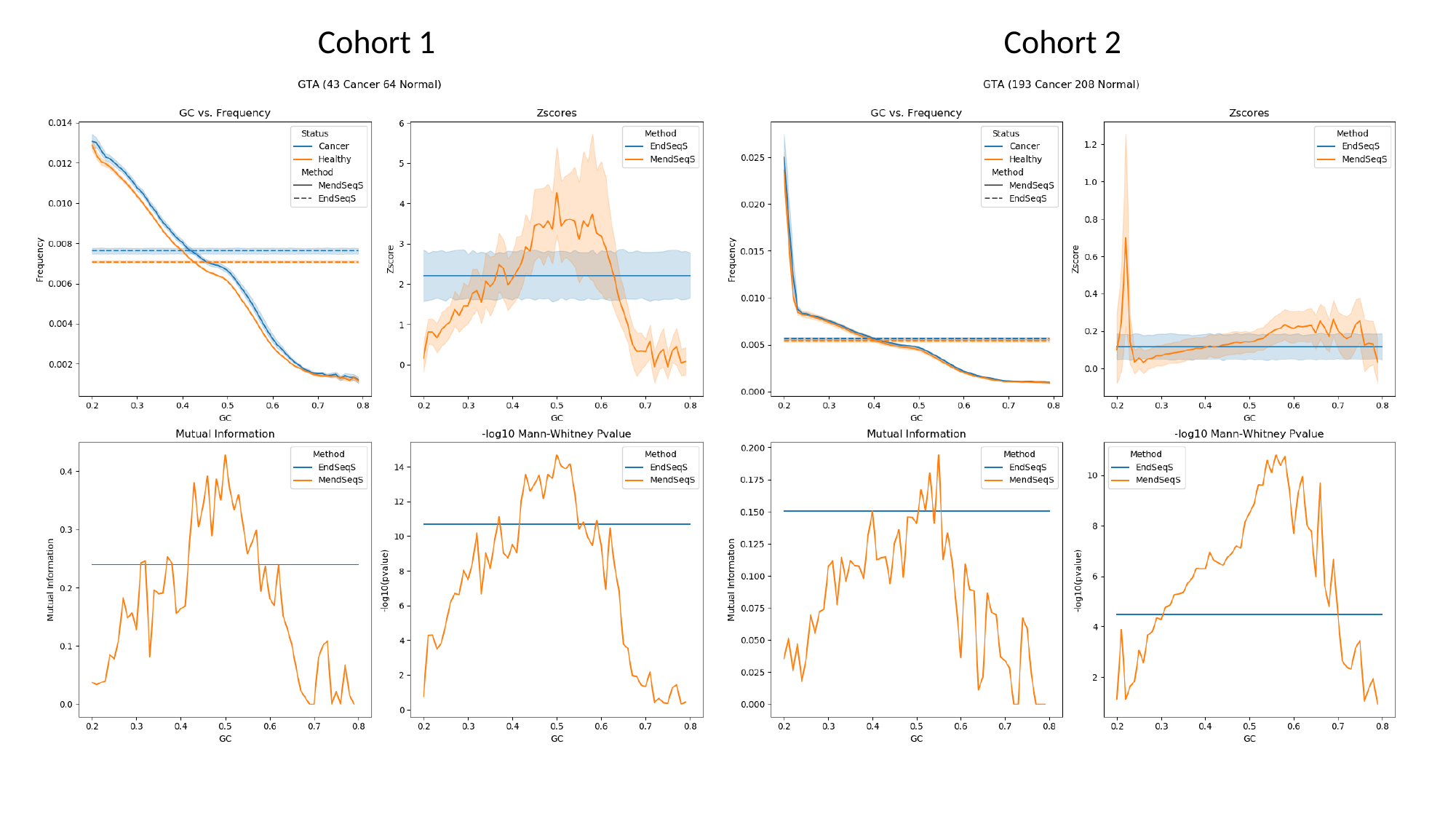

Cohort 2
Cohort 1

### Slide 22
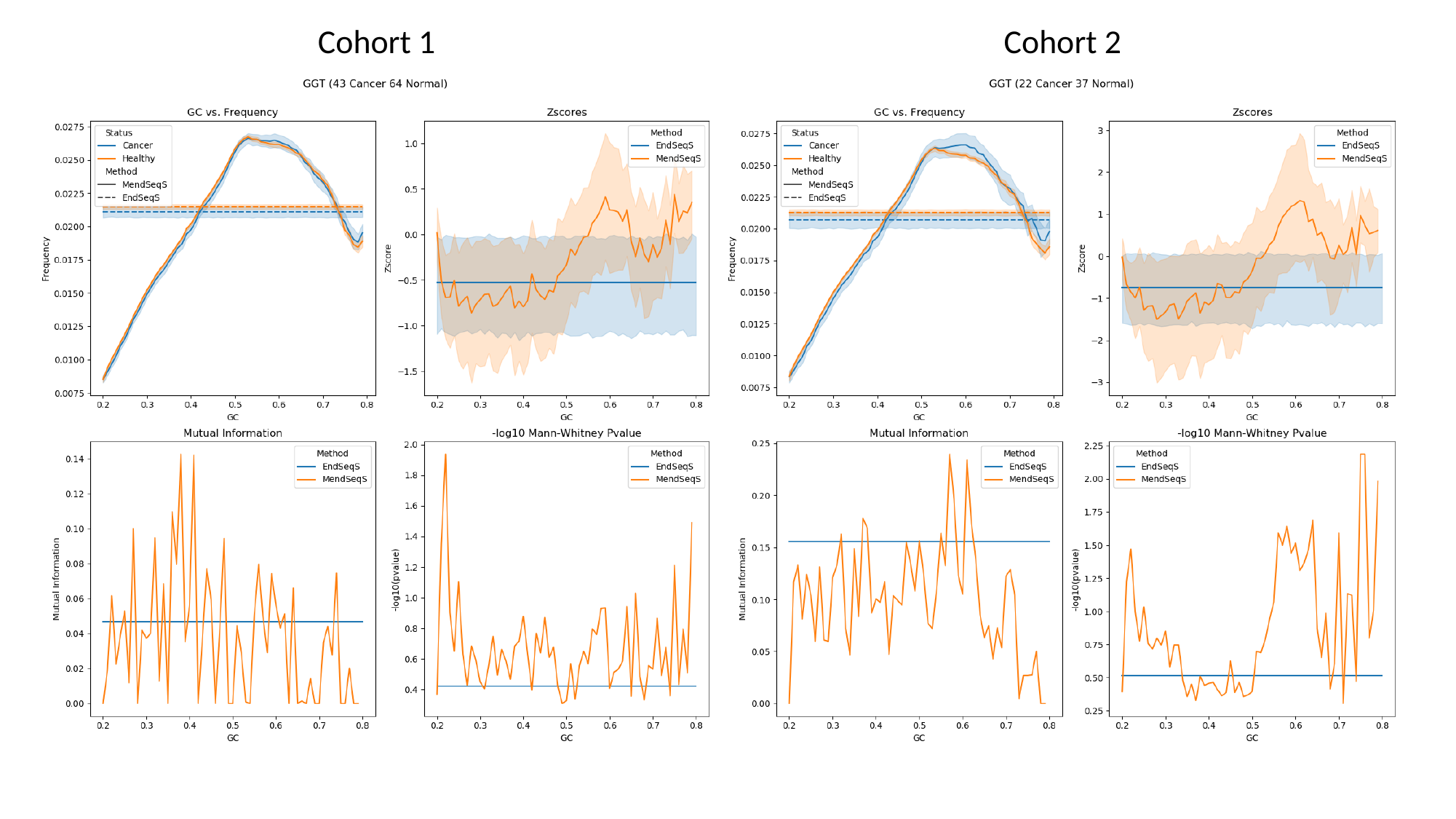

Cohort 2
Cohort 1

### Slide 23
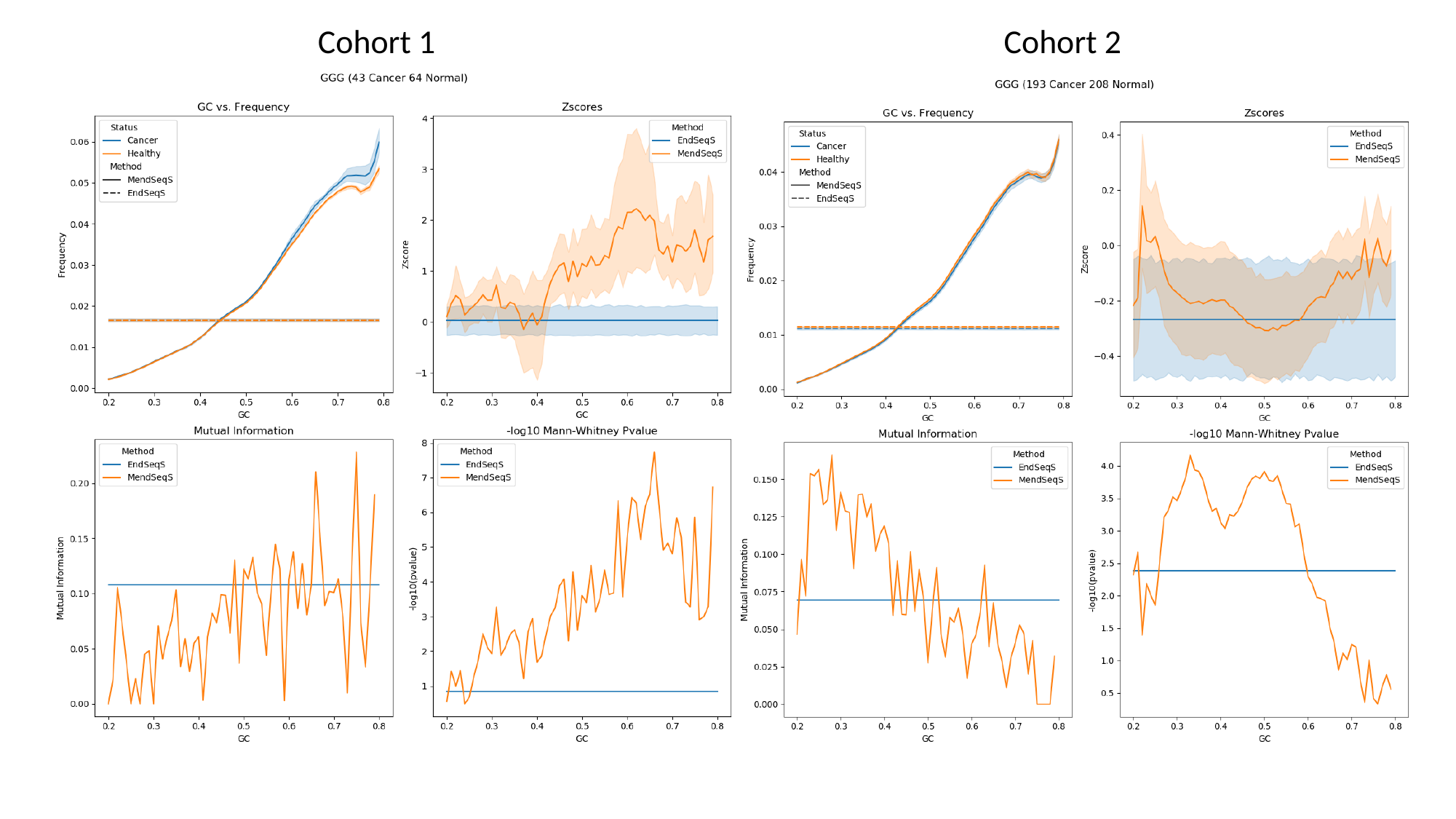

Cohort 2
Cohort 1

### Slide 24
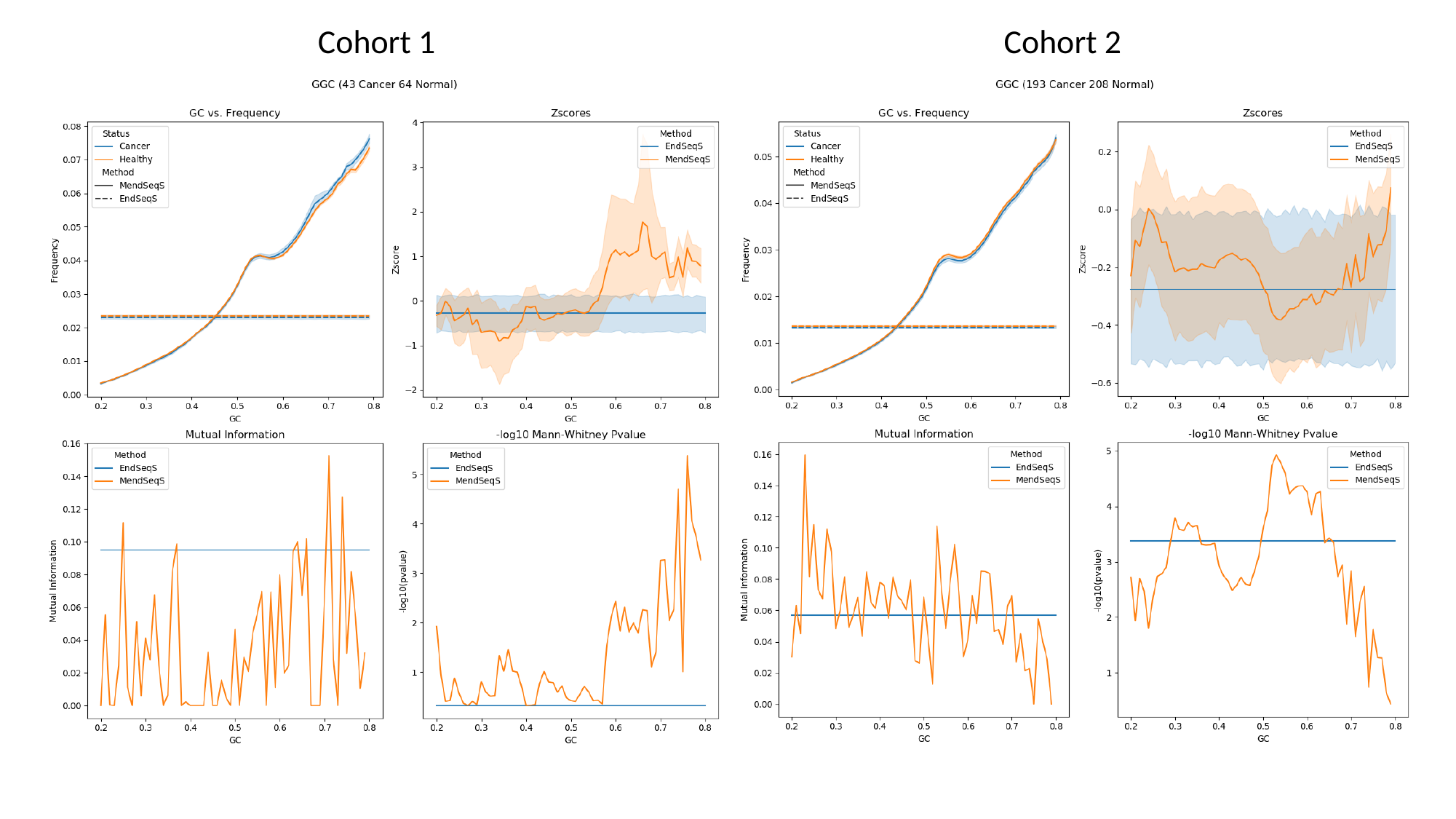

Cohort 2
Cohort 1

### Slide 25
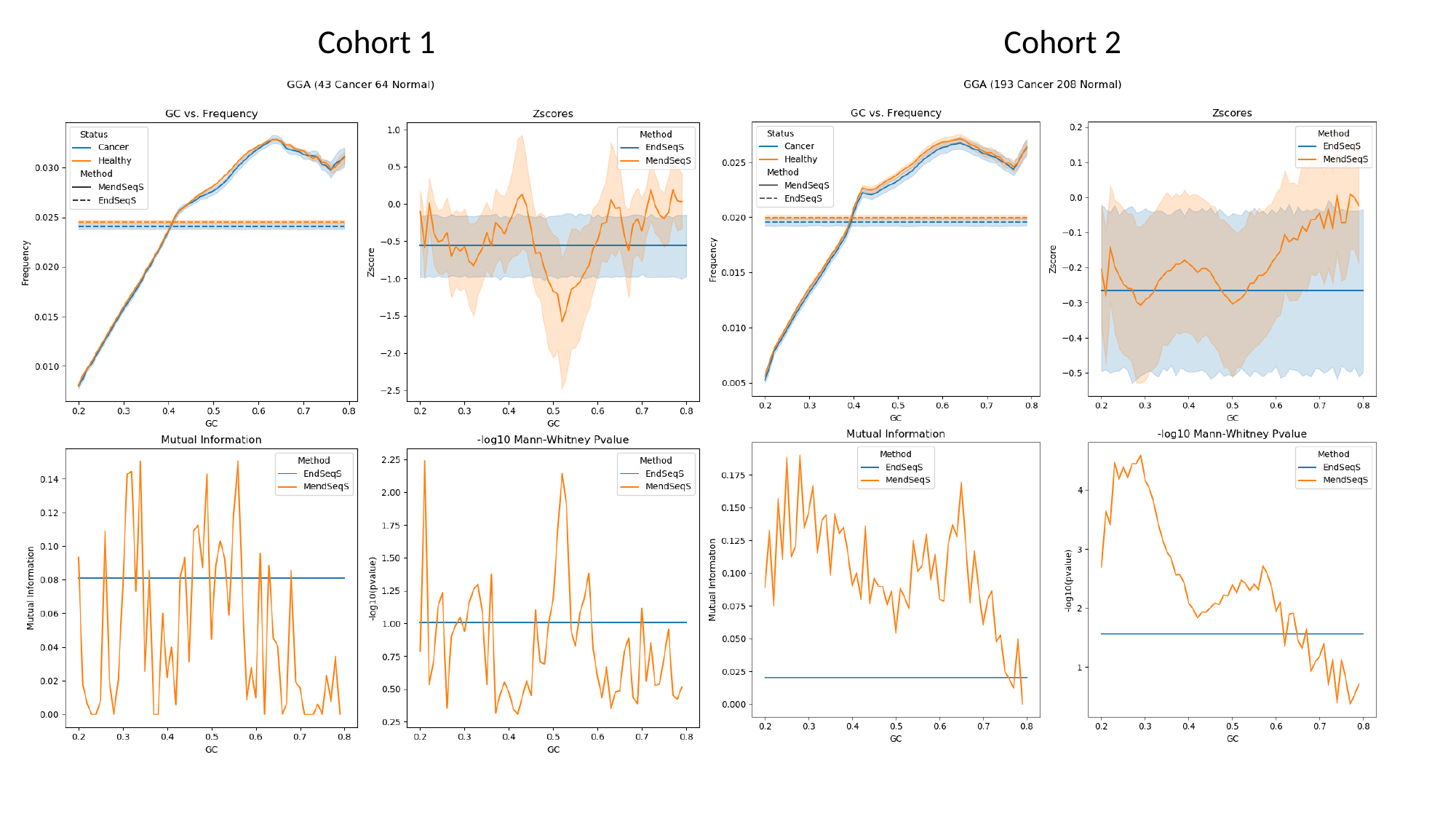

Cohort 2
Cohort 1

### Slide 26
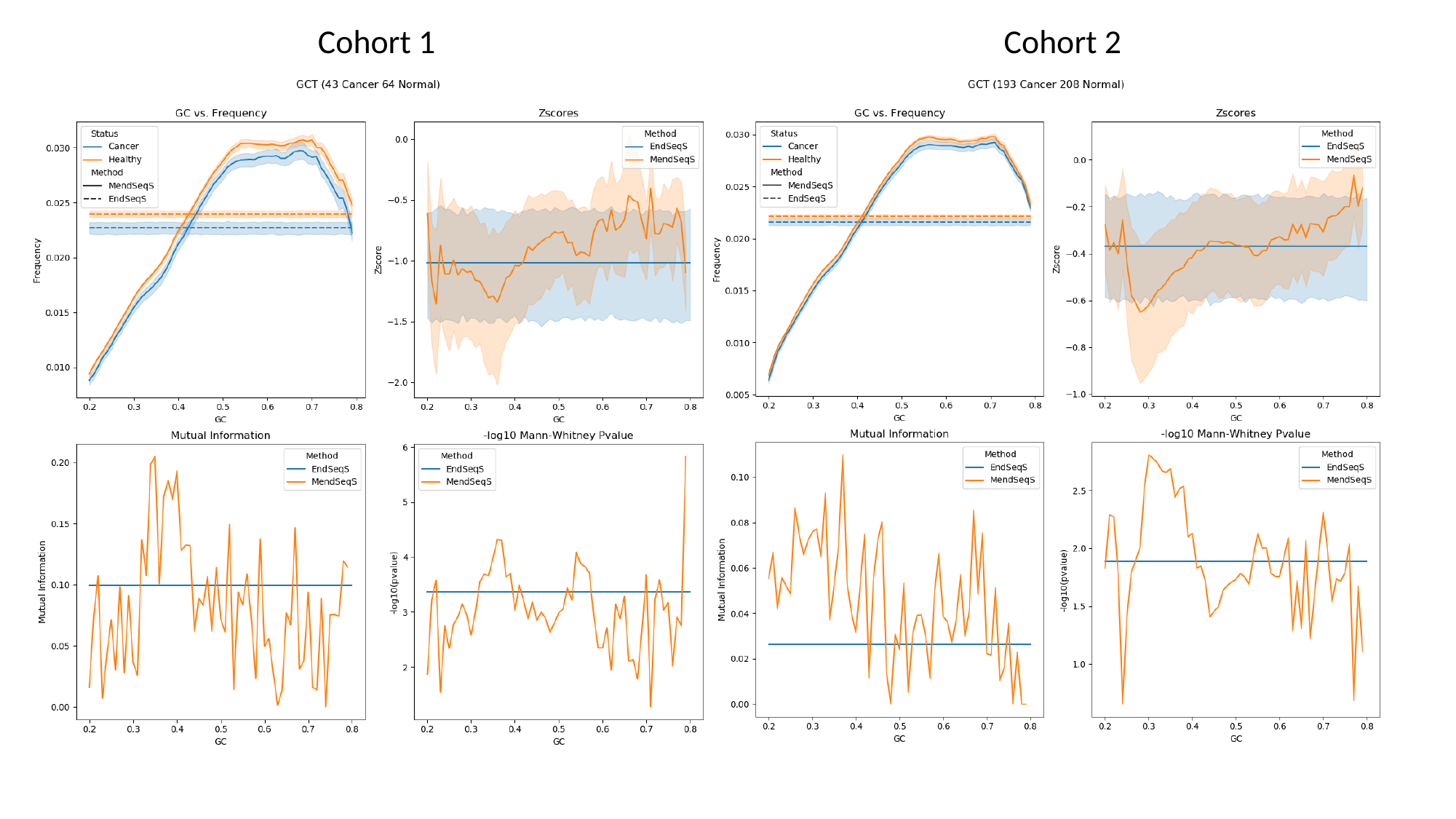

Cohort 2
Cohort 1

### Slide 27
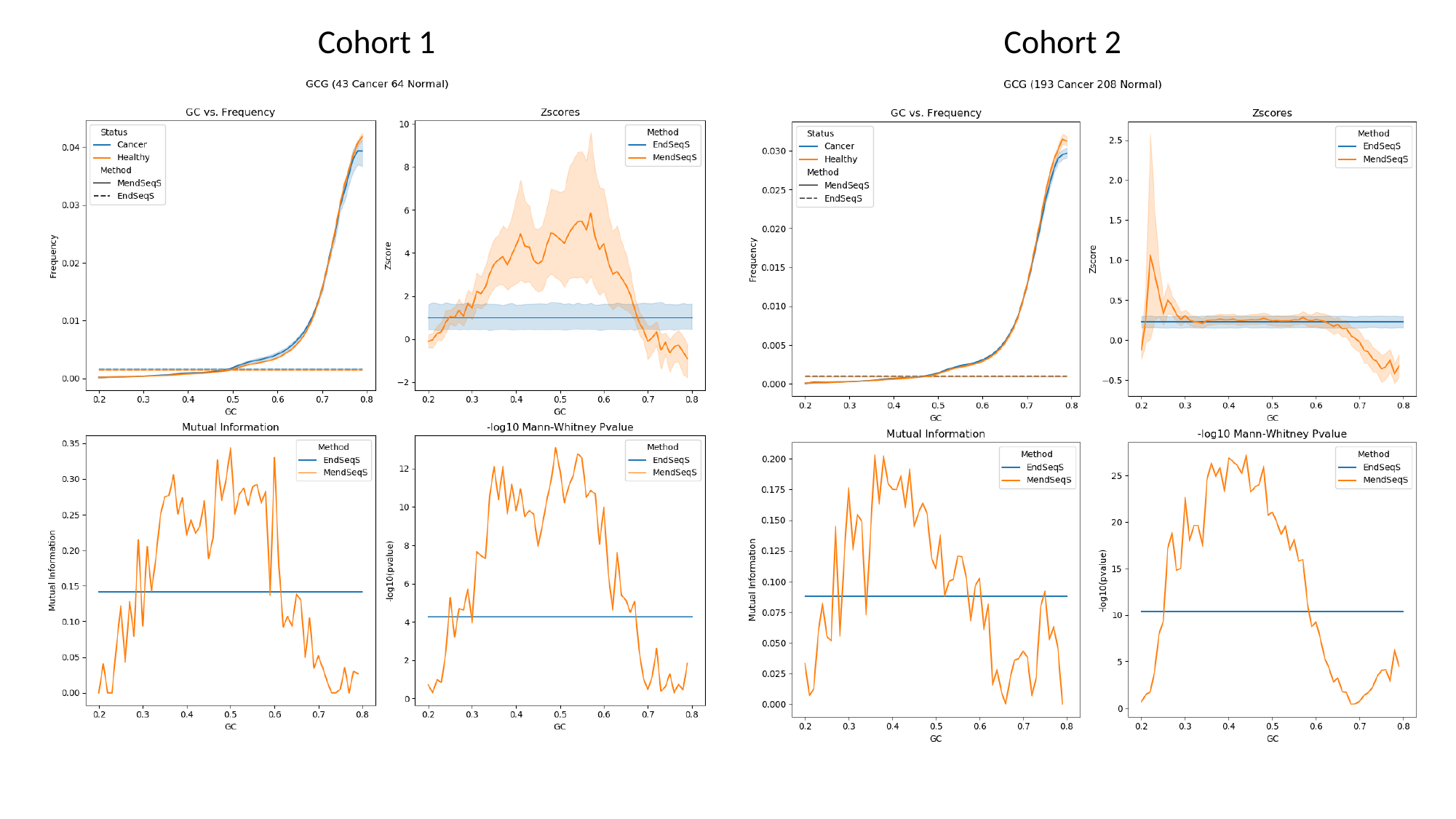

Cohort 2
Cohort 1

### Slide 28
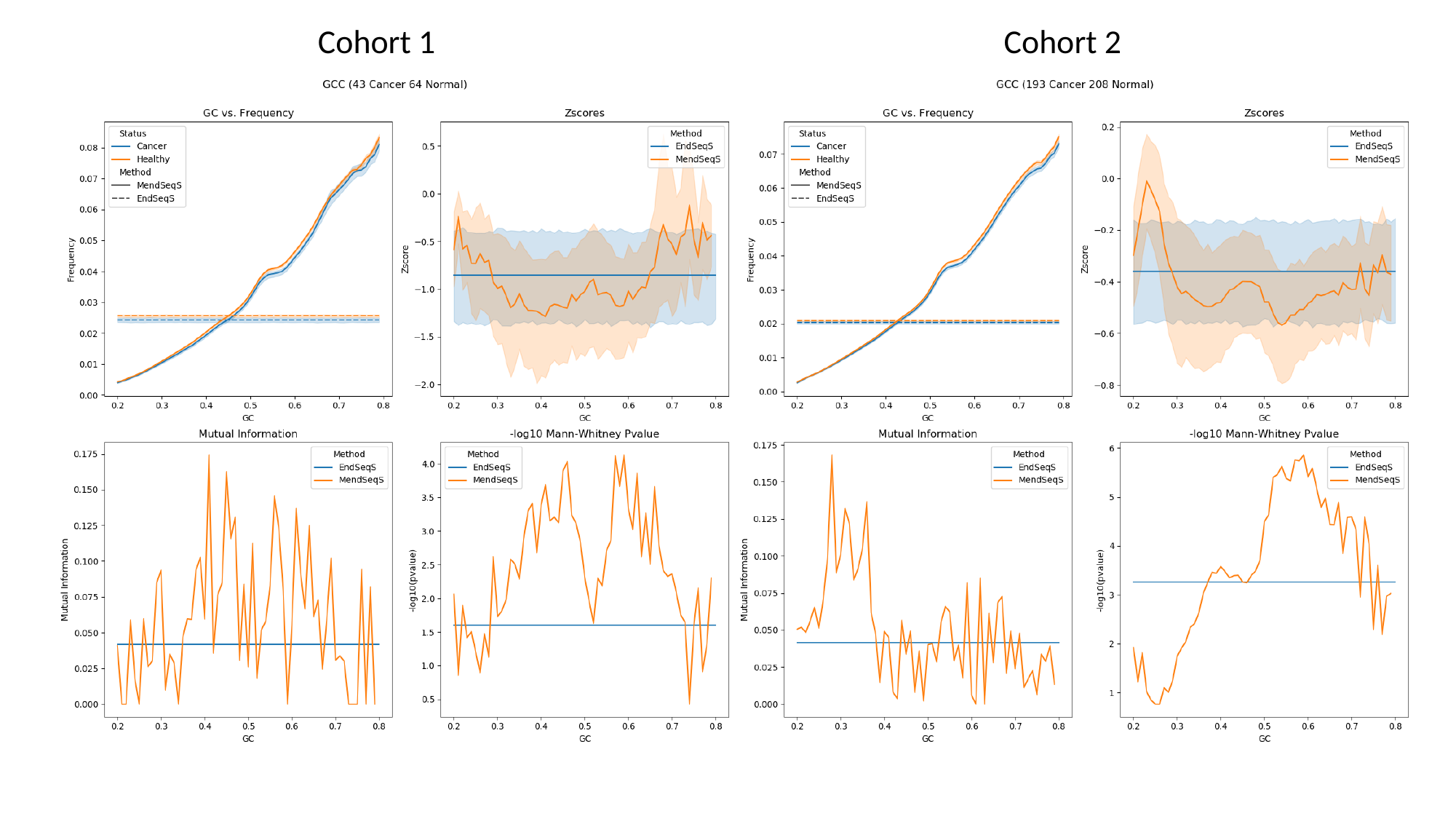

Cohort 2
Cohort 1

### Slide 29
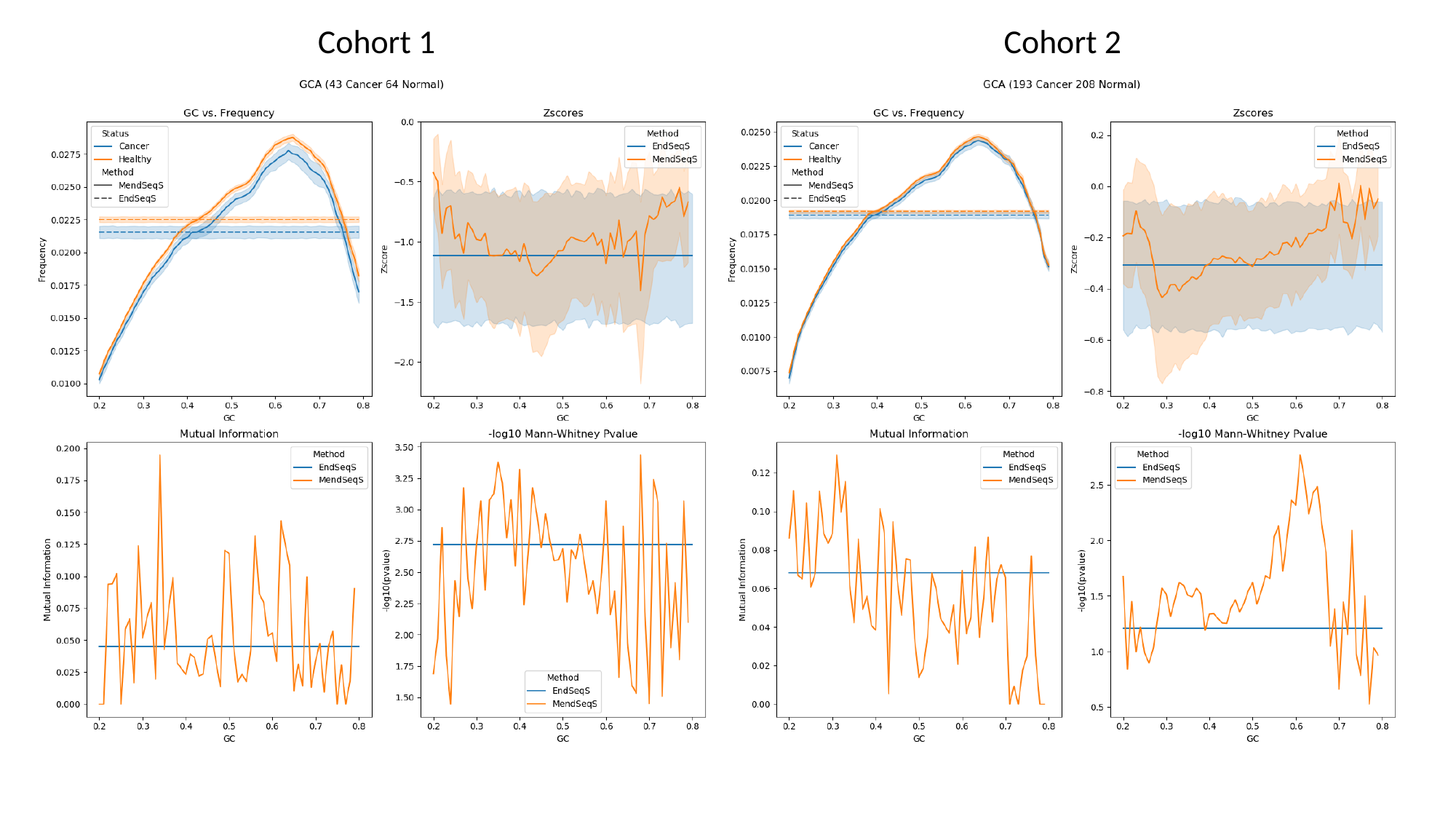

Cohort 2
Cohort 1

### Slide 30
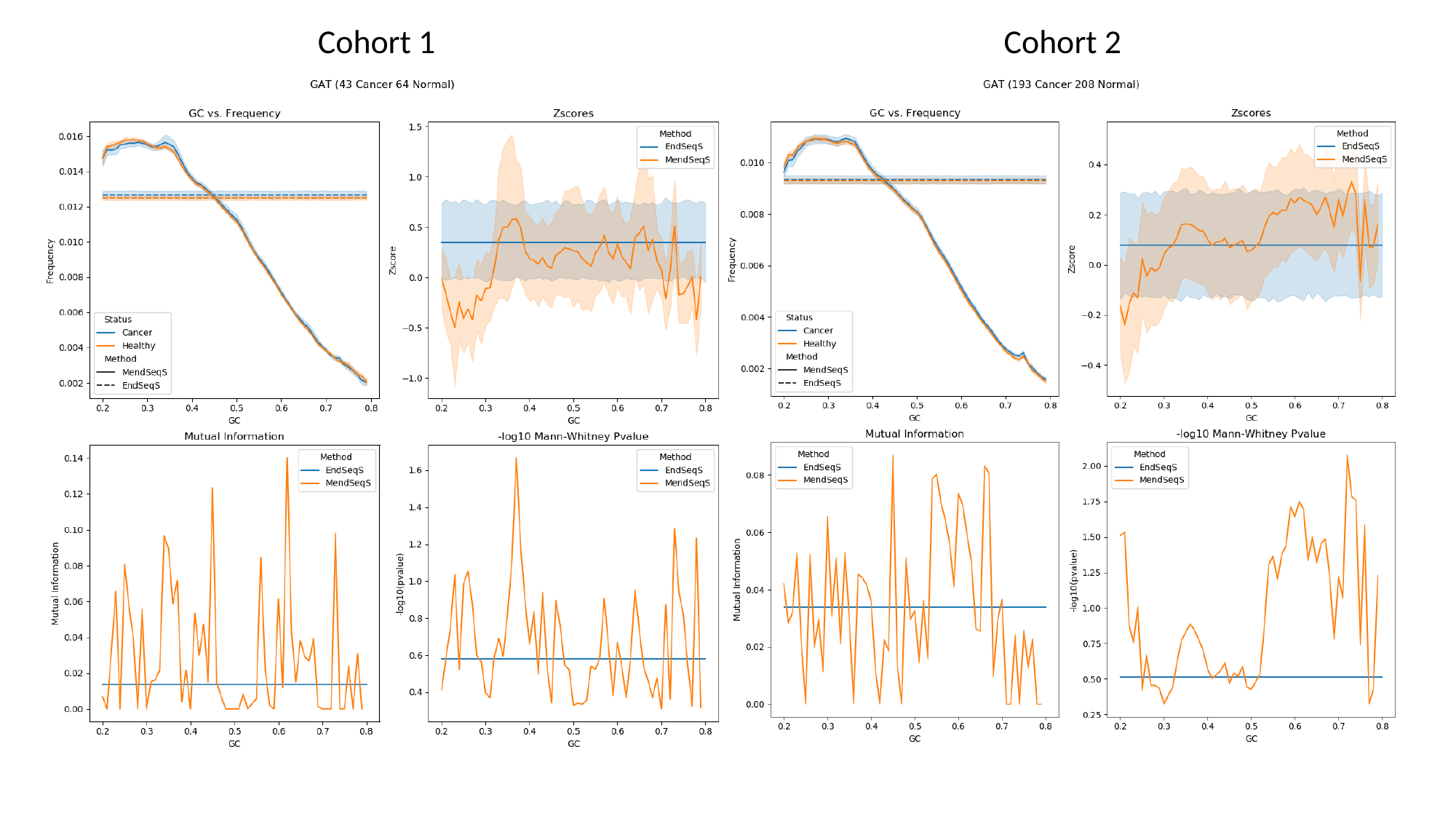

Cohort 2
Cohort 1

### Slide 31
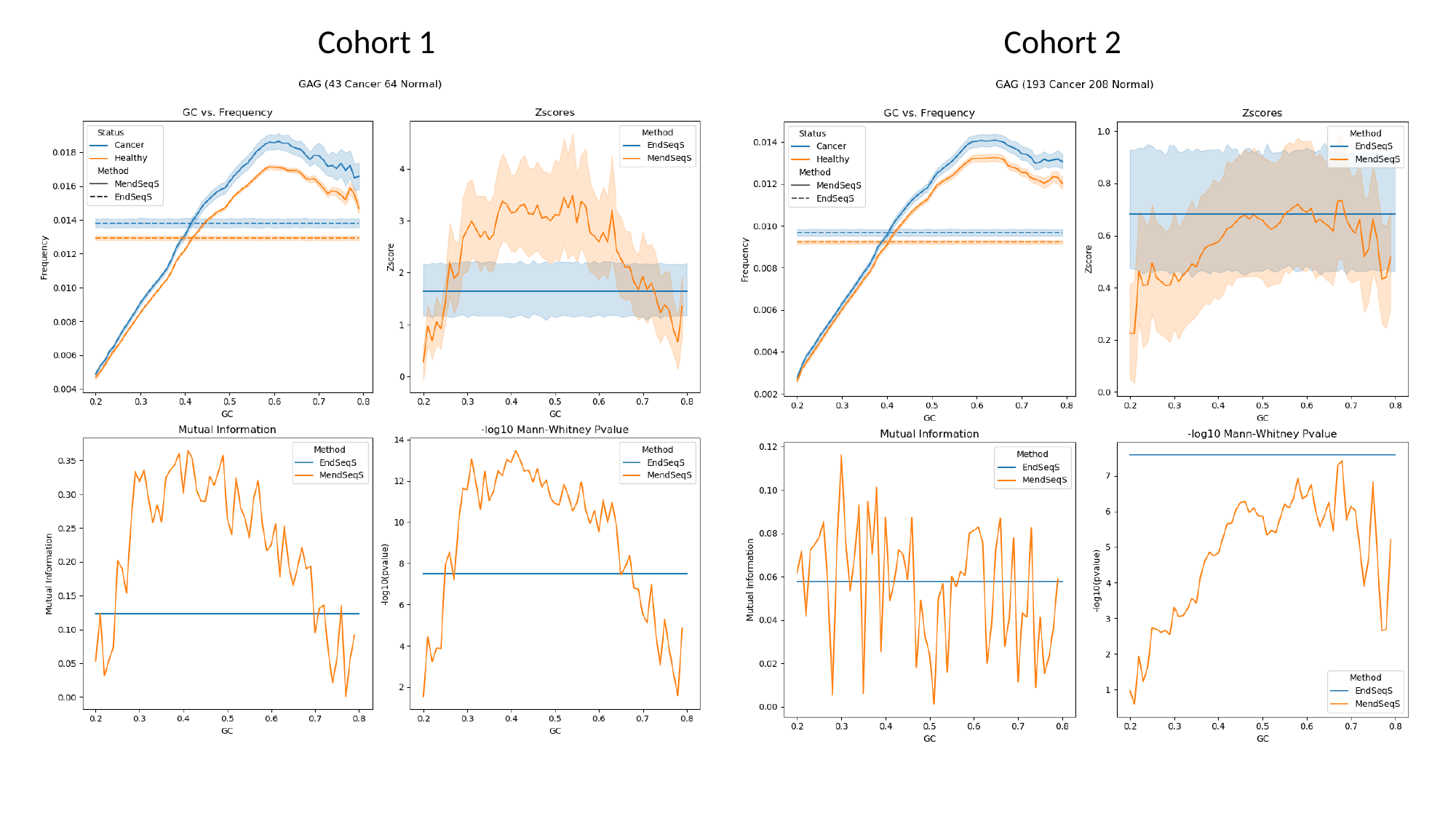

Cohort 2
Cohort 1

### Slide 32
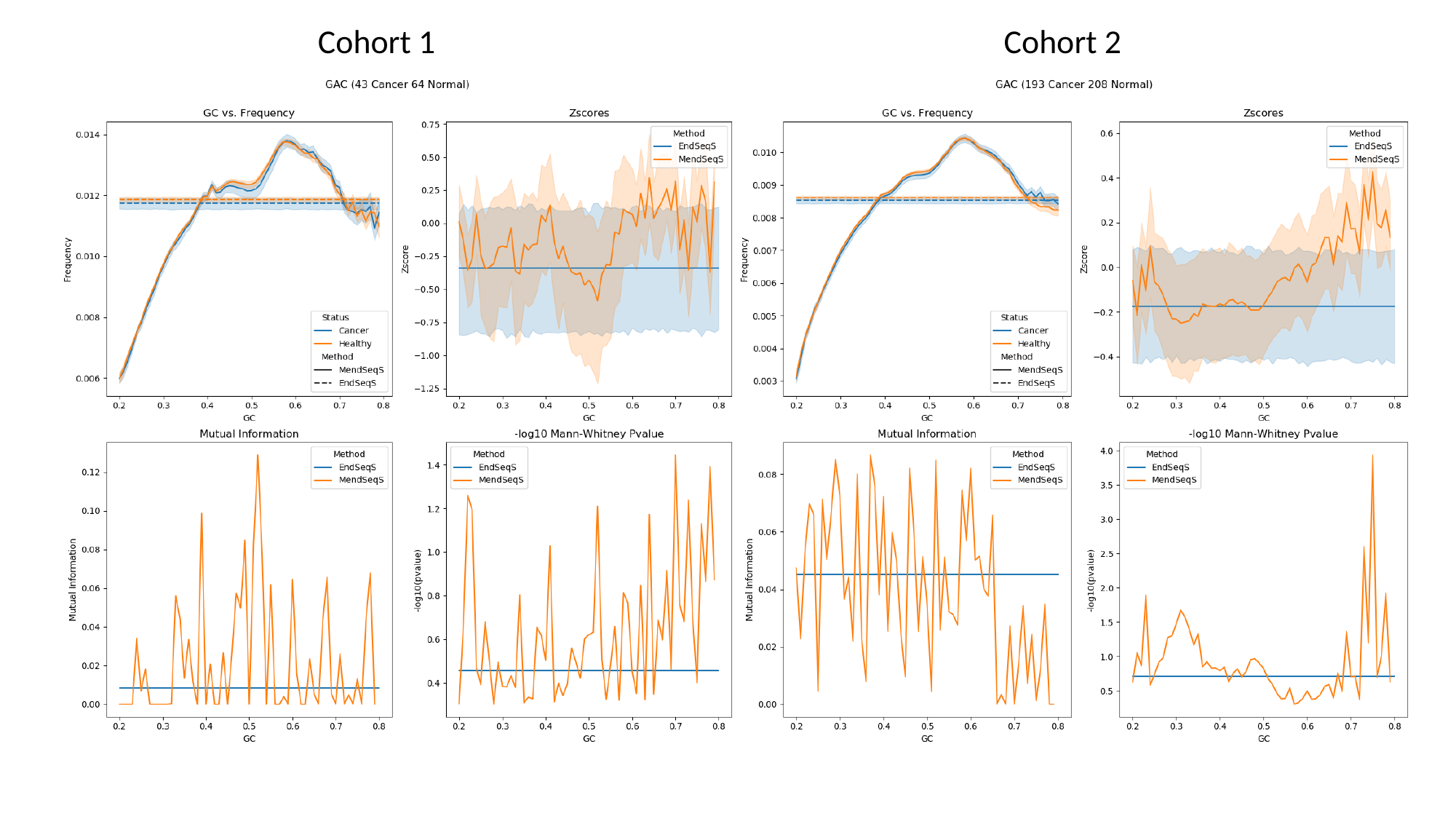

Cohort 2
Cohort 1

### Slide 33
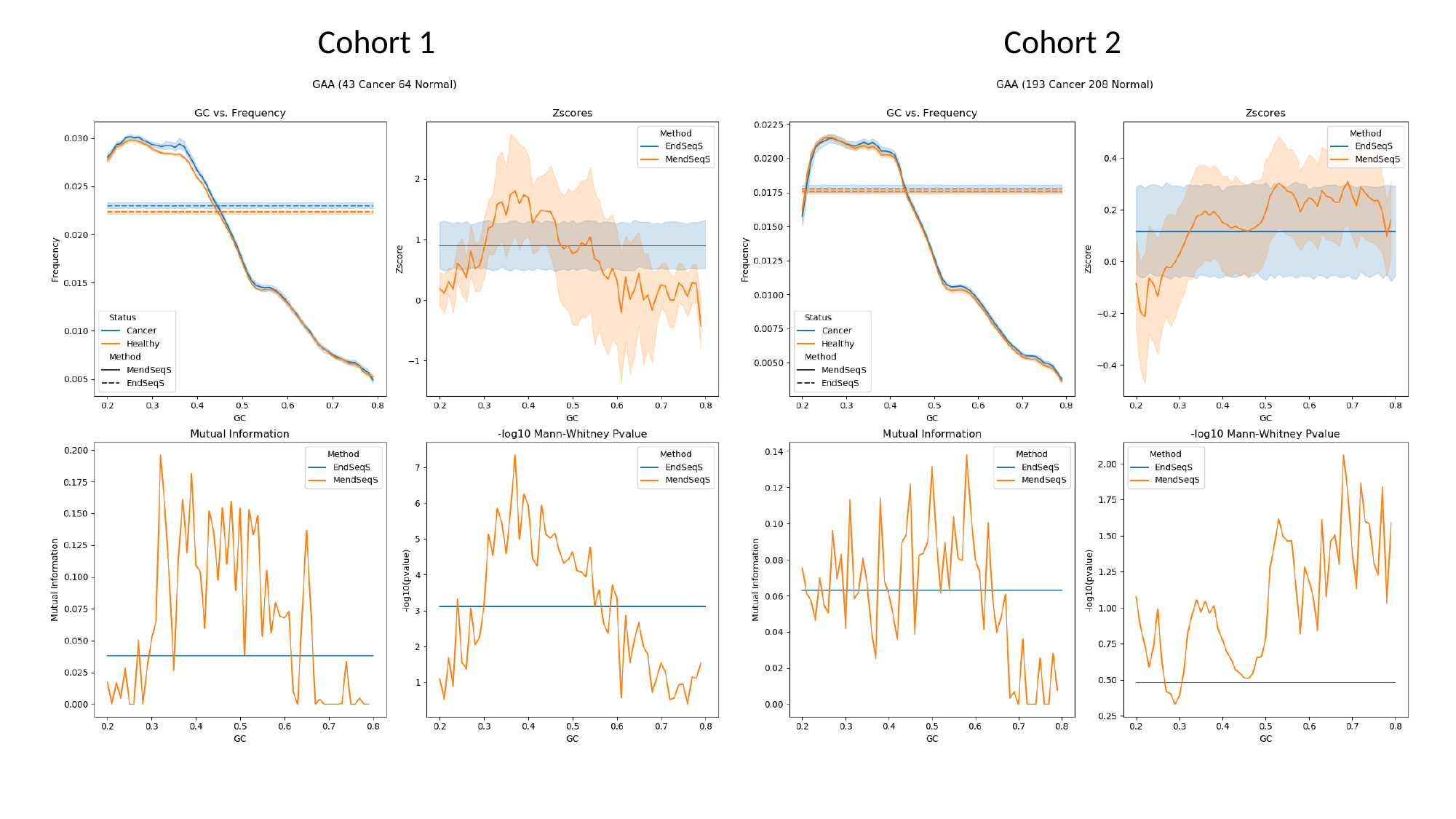

Cohort 2
Cohort 1

### Slide 34
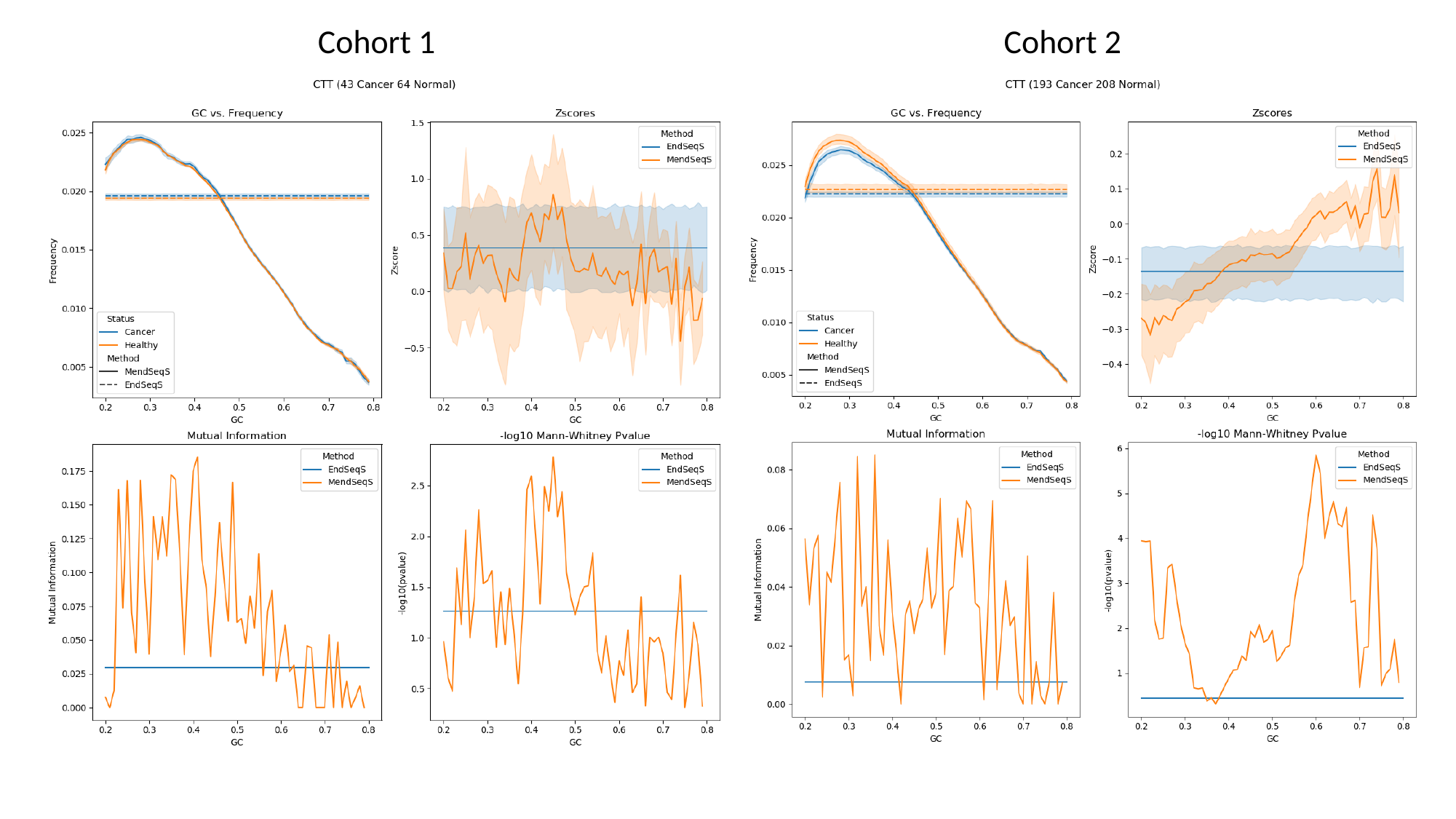

Cohort 2
Cohort 1

### Slide 35
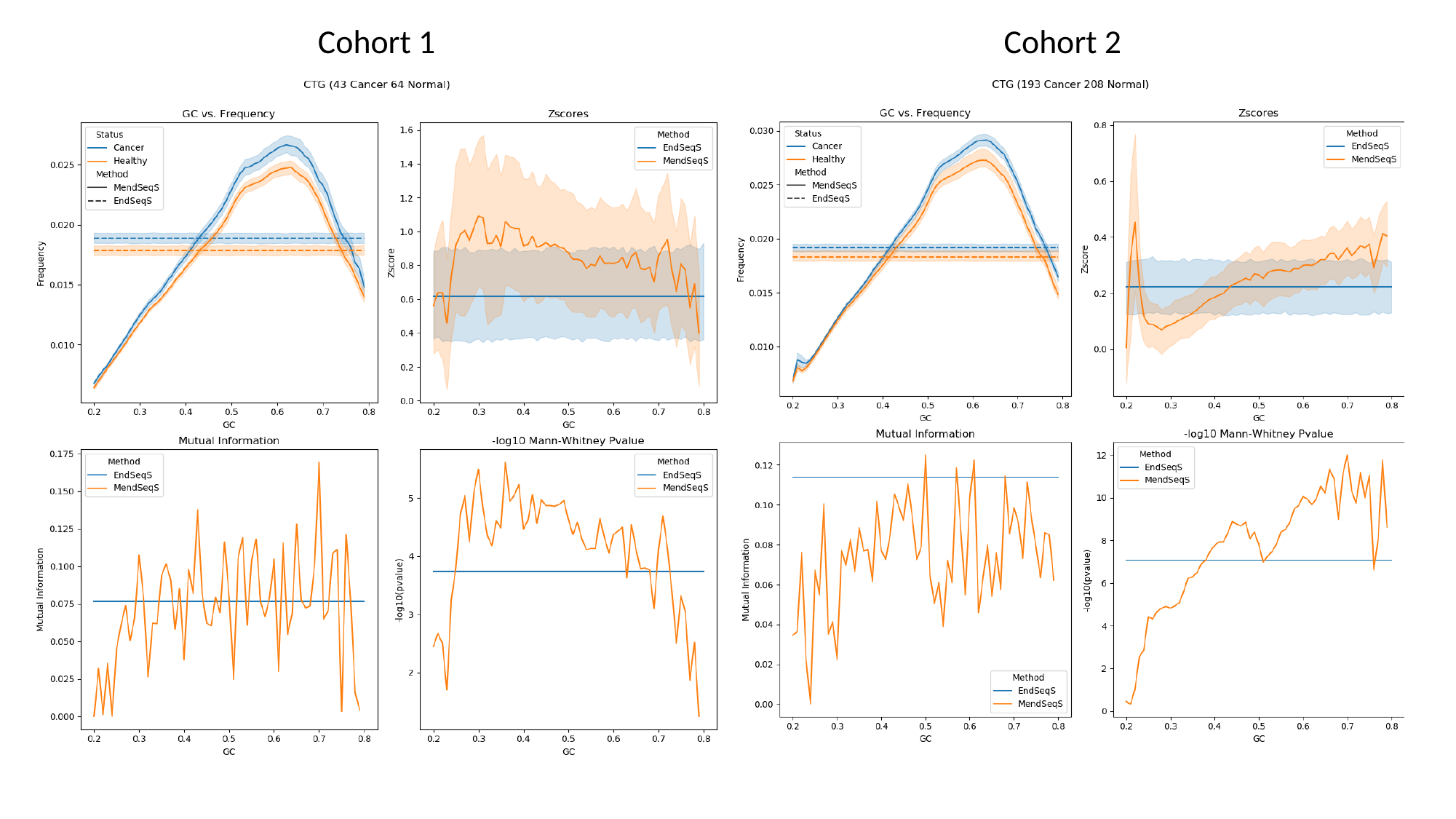

Cohort 2
Cohort 1

### Slide 36
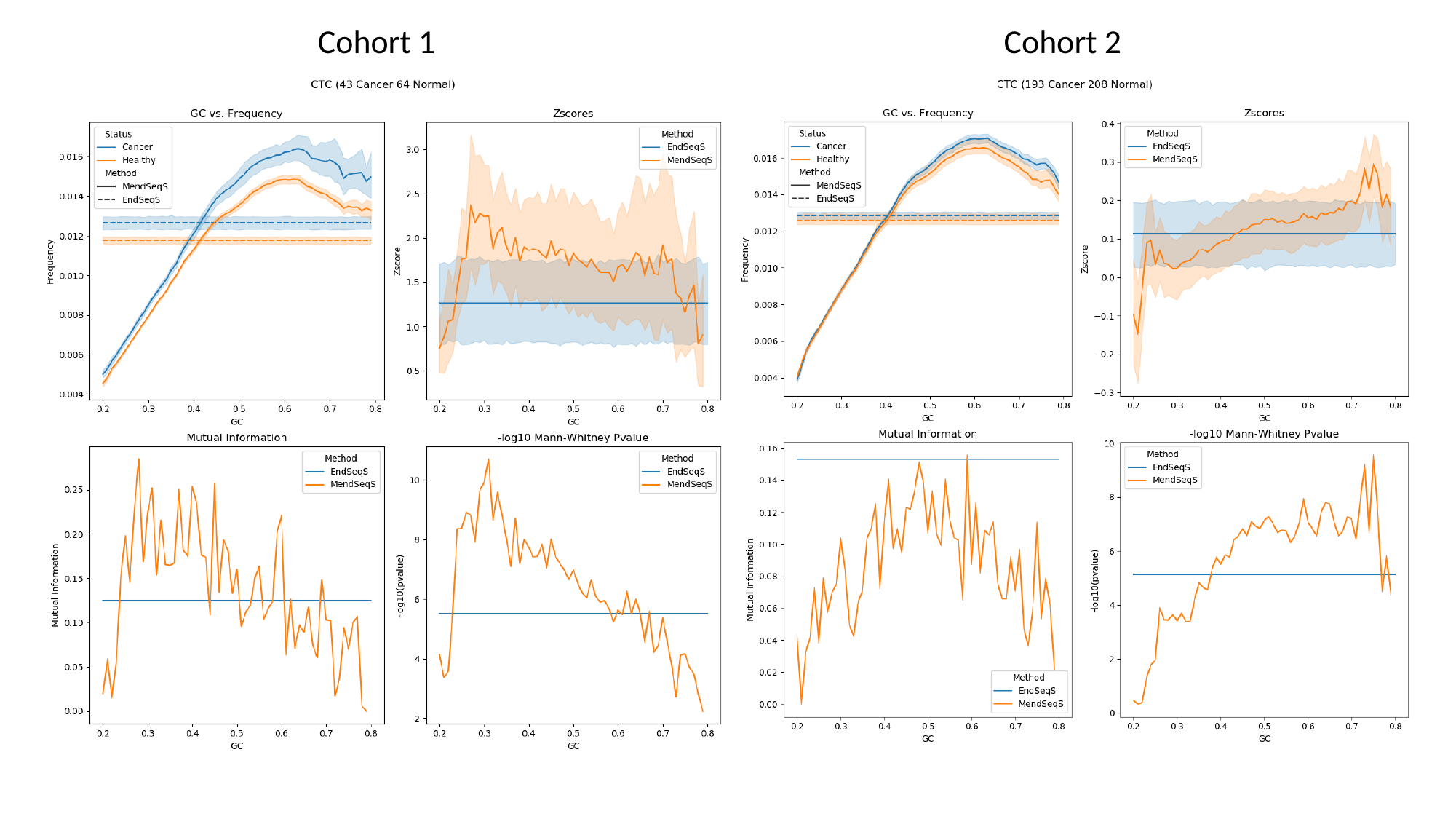

Cohort 2
Cohort 1

### Slide 37
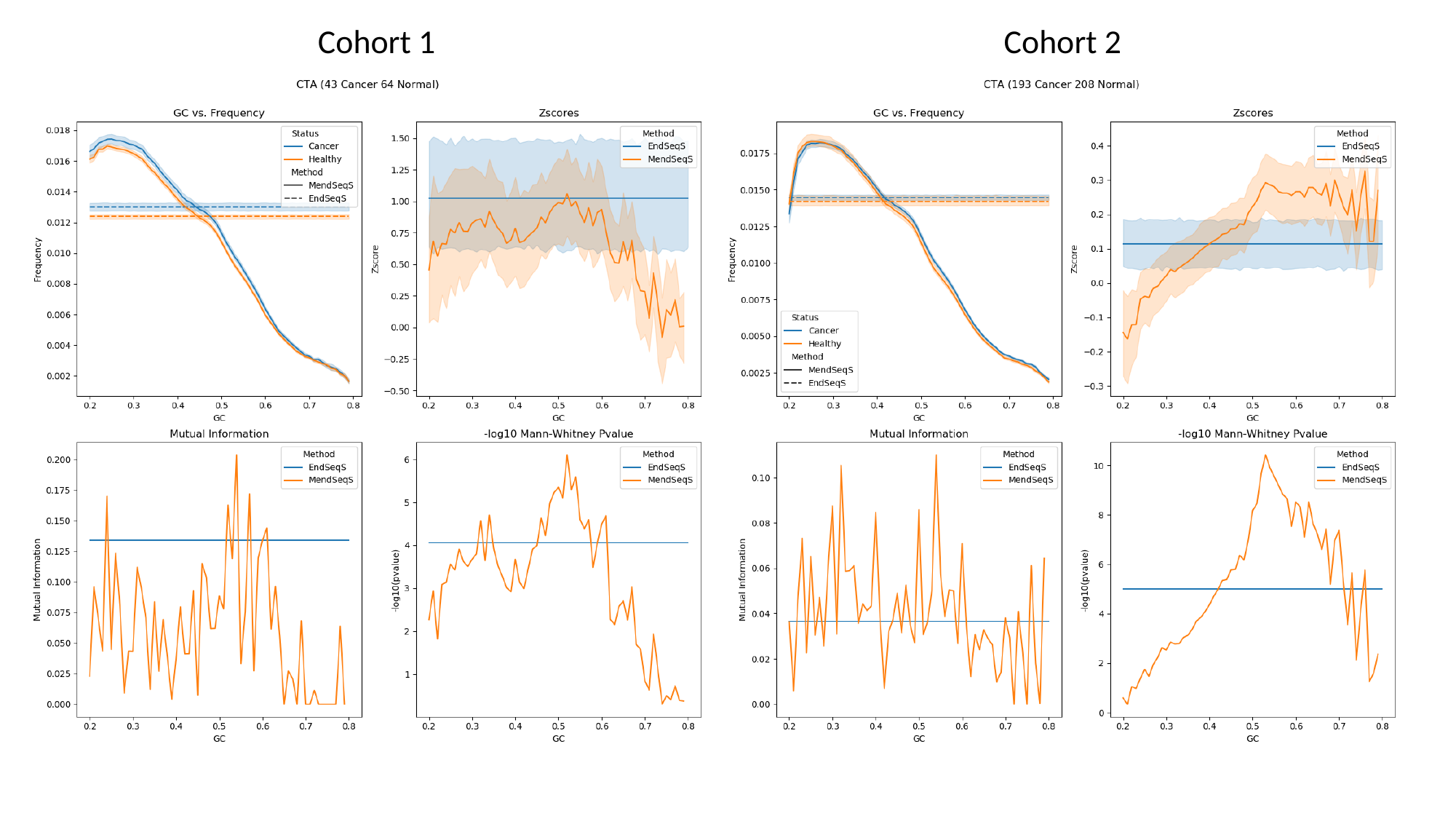

Cohort 2
Cohort 1

### Slide 38
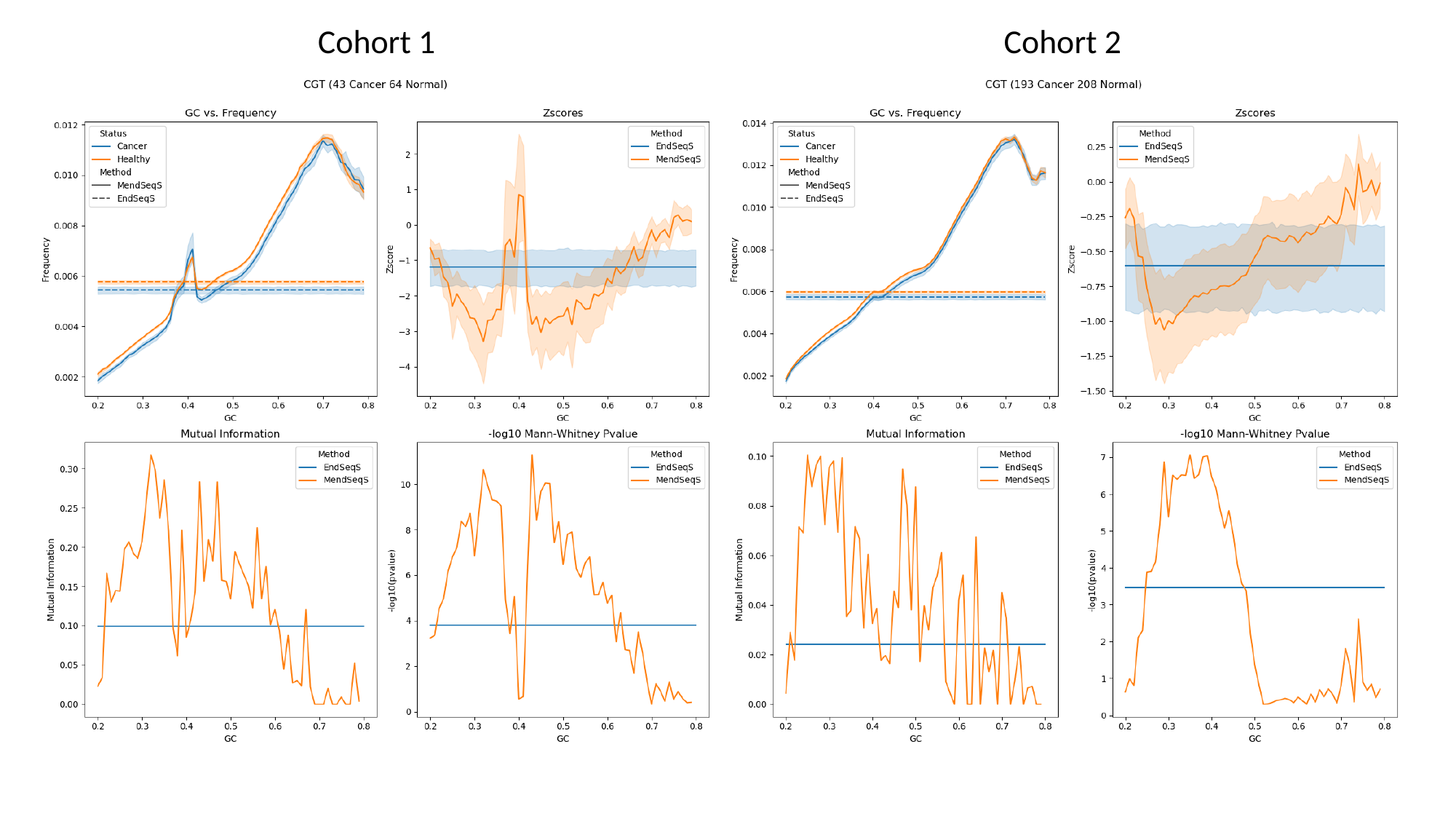

Cohort 2
Cohort 1

### Slide 39
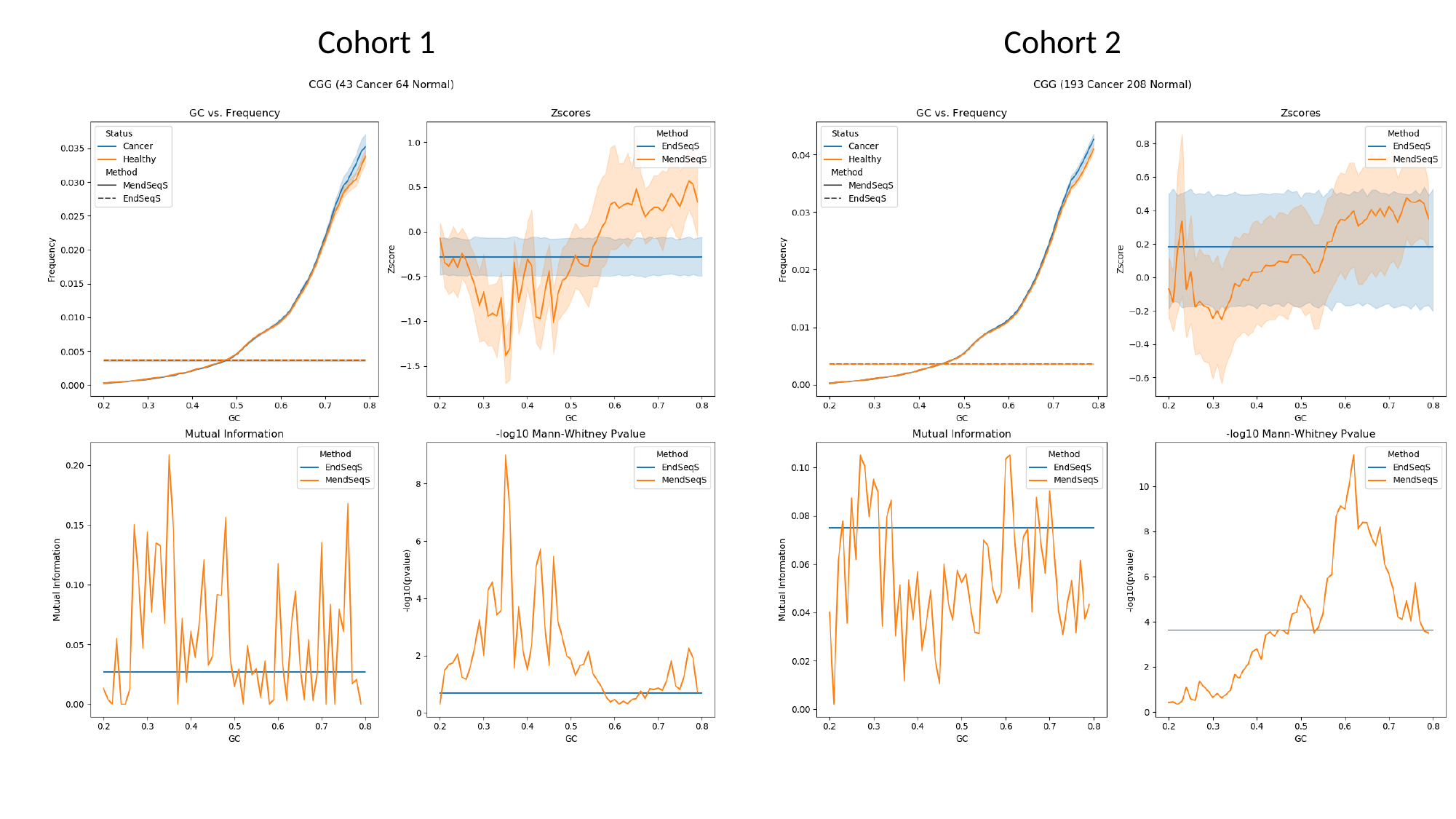

Cohort 2
Cohort 1

### Slide 40
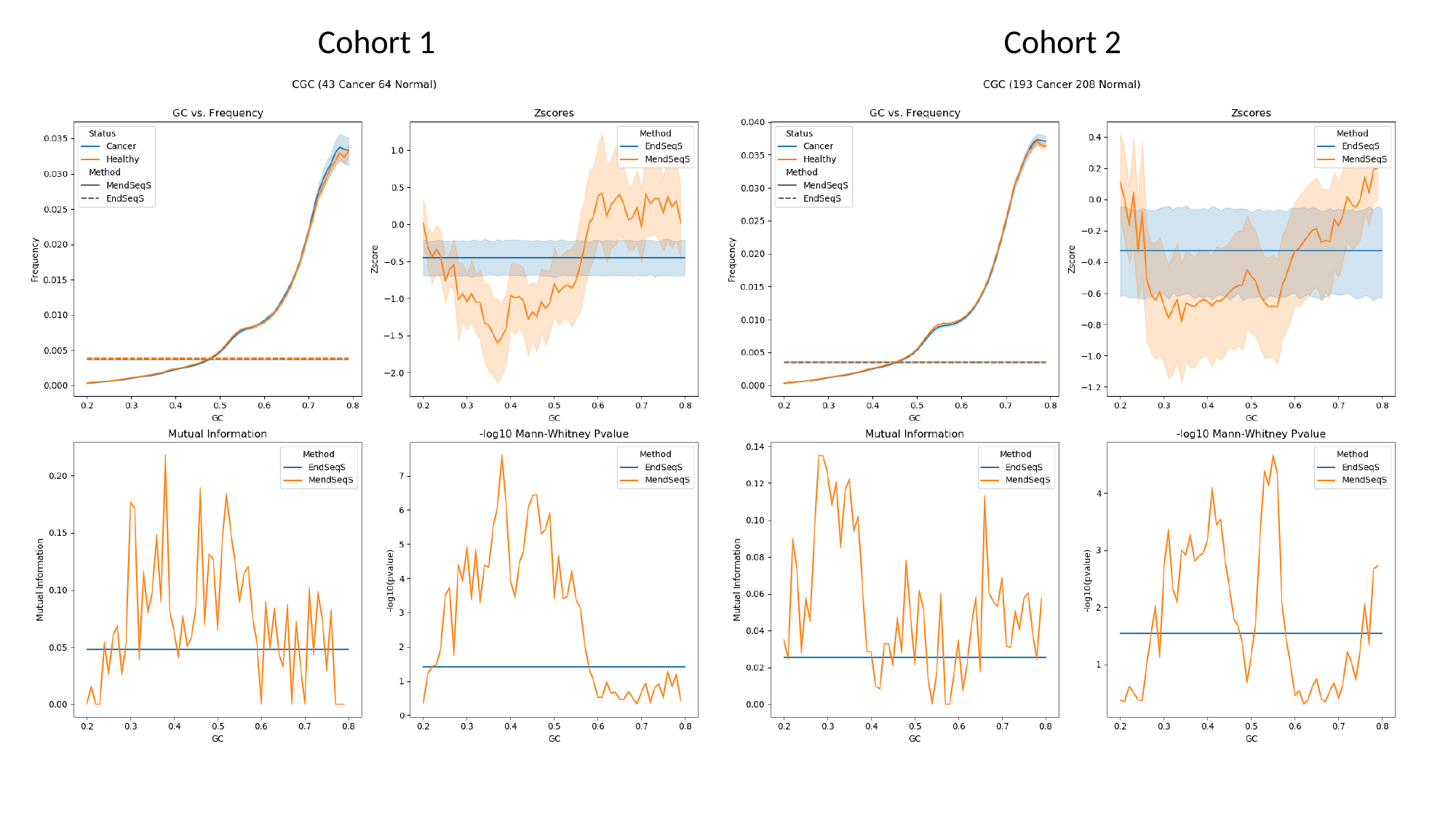

Cohort 2
Cohort 1

### Slide 41
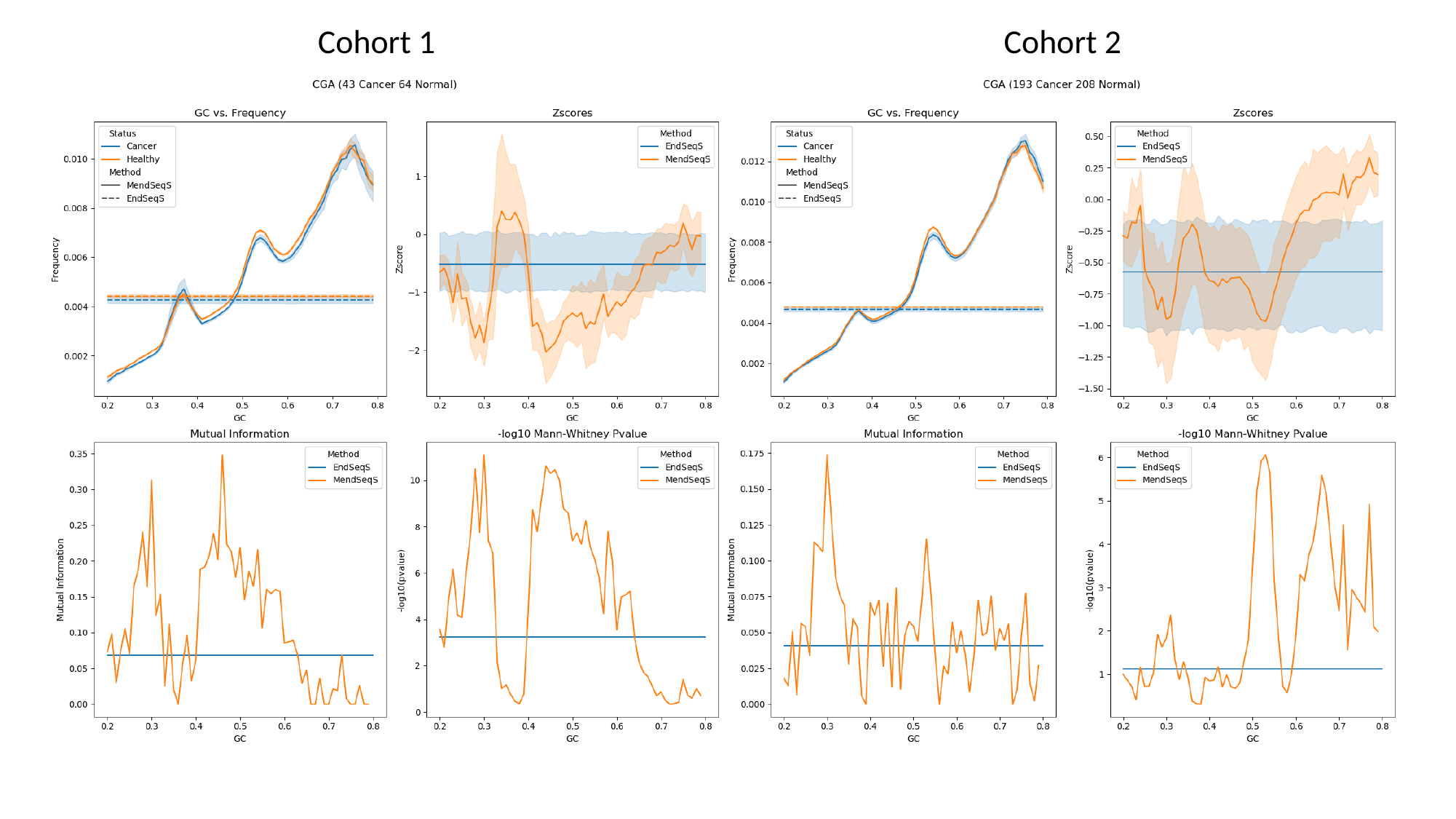

Cohort 2
Cohort 1

### Slide 42
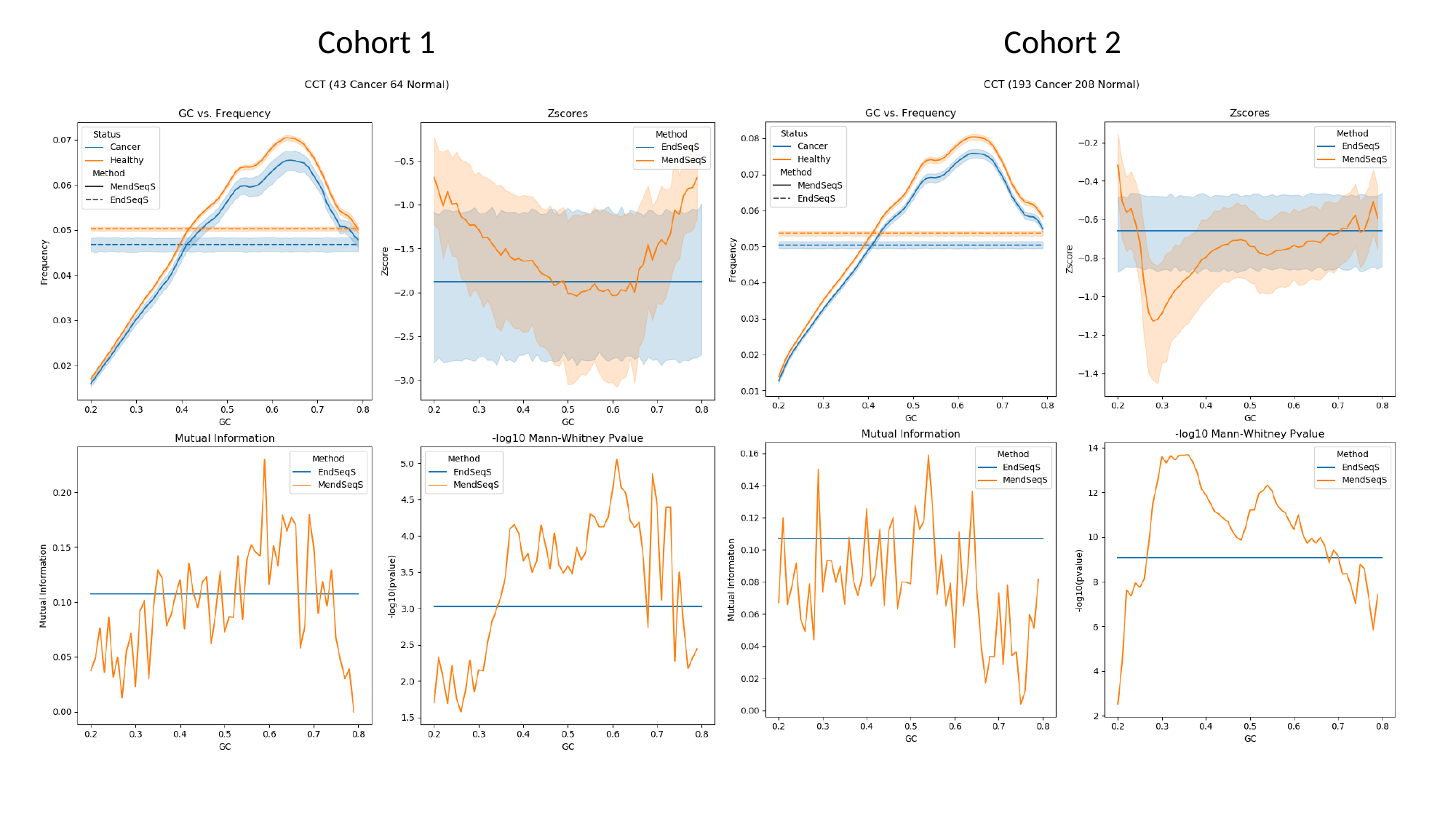

Cohort 2
Cohort 1

### Slide 43
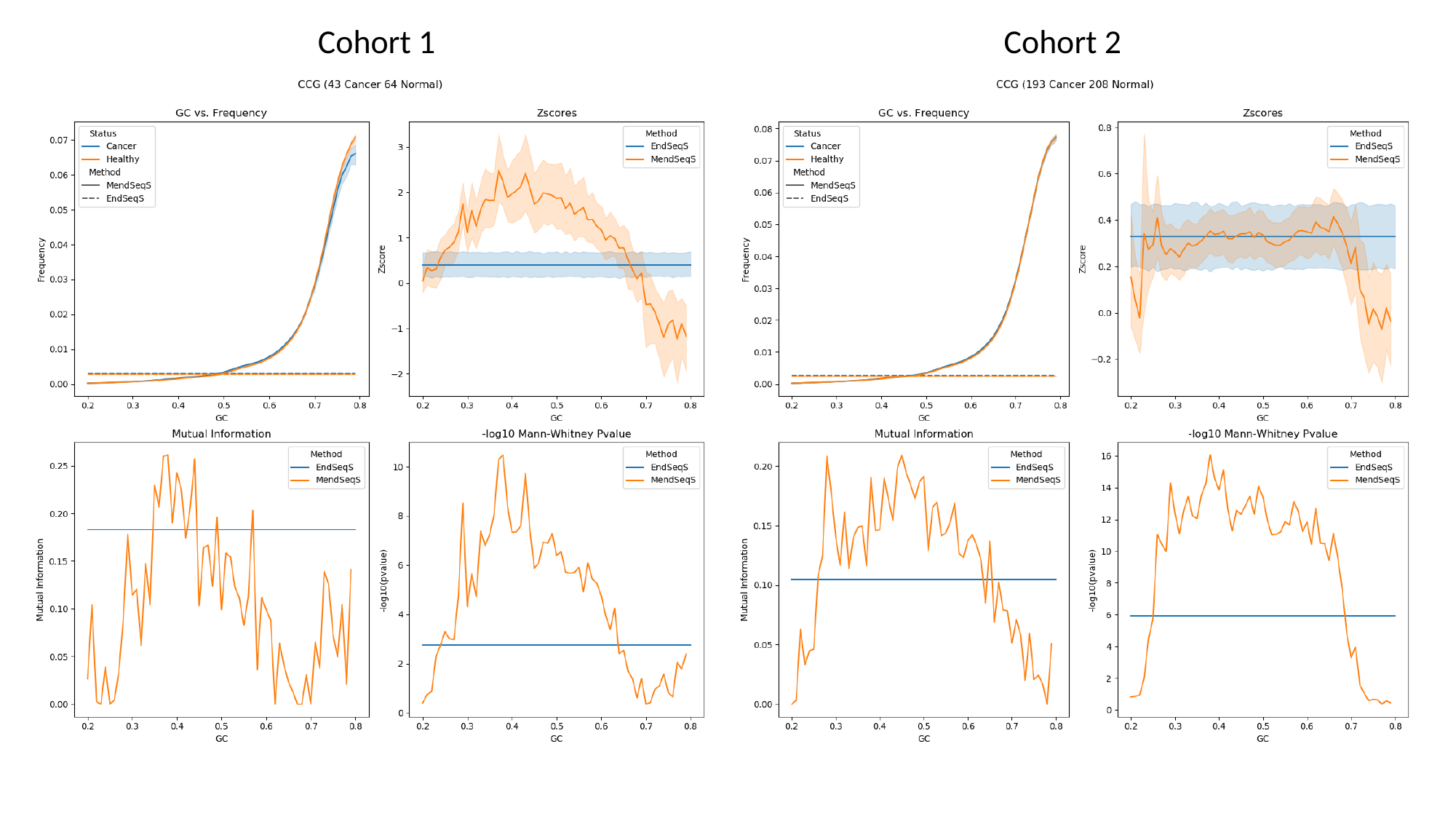

Cohort 2
Cohort 1

### Slide 44
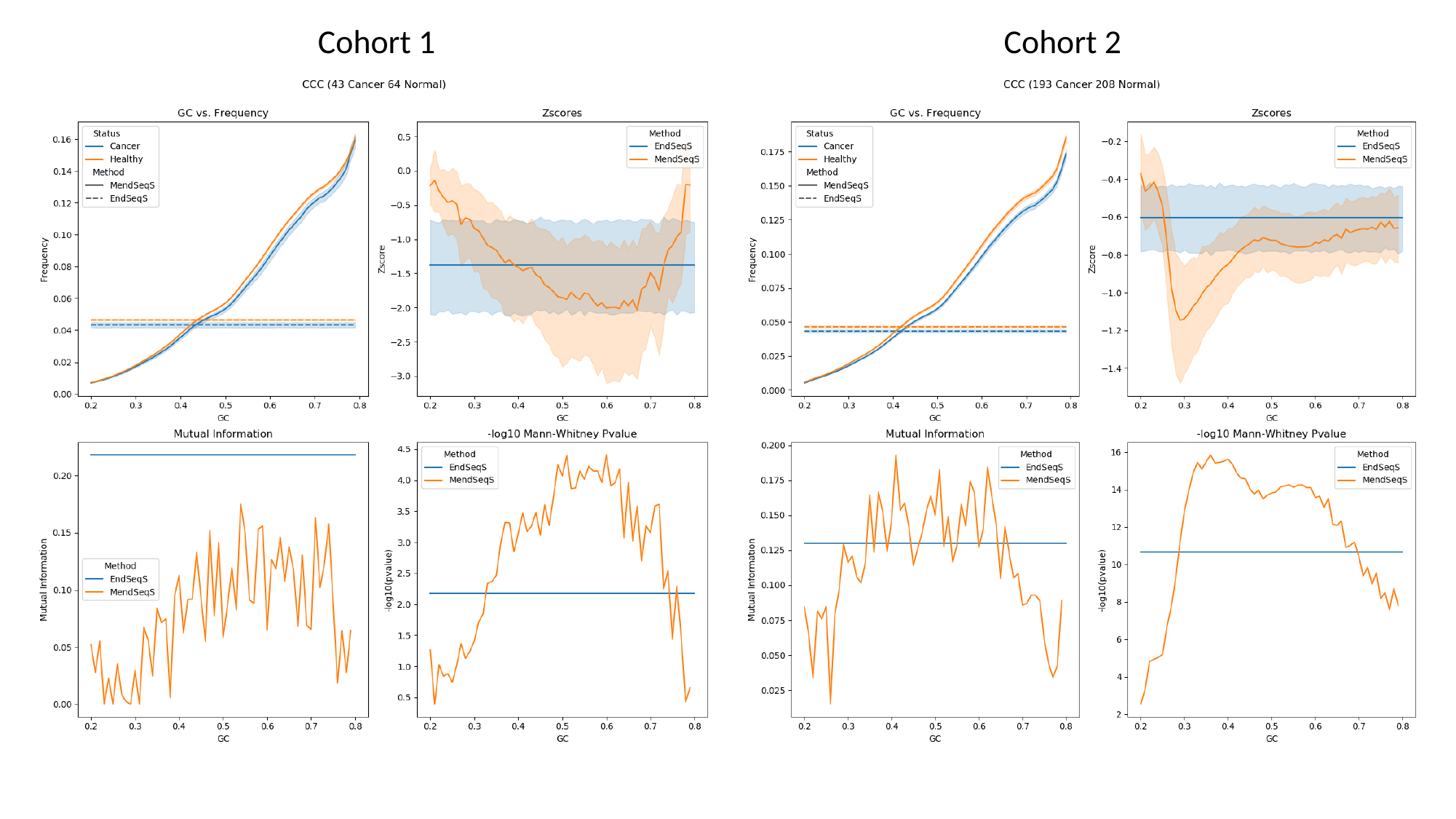

Cohort 2
Cohort 1

### Slide 45
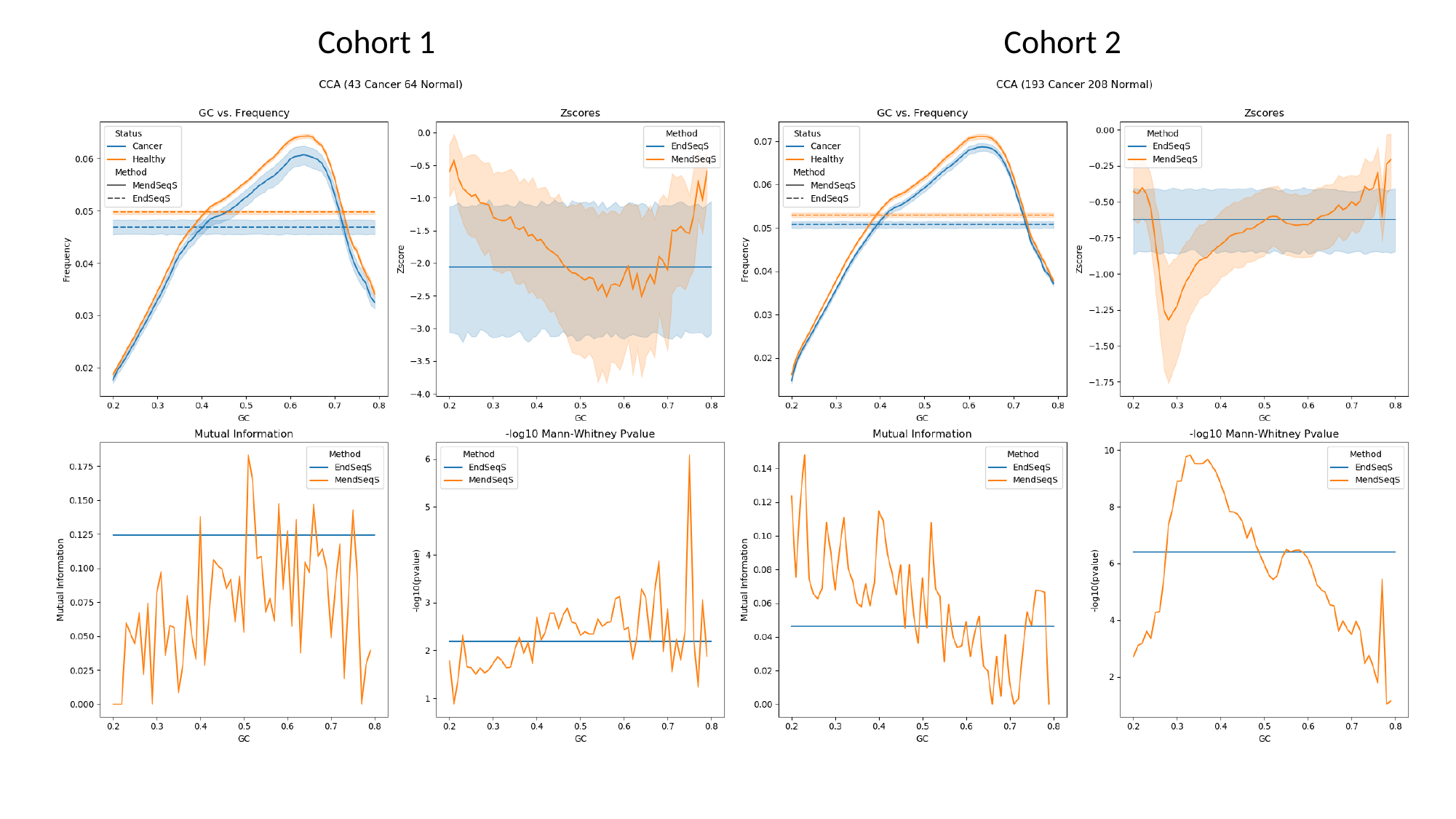

Cohort 2
Cohort 1

### Slide 46
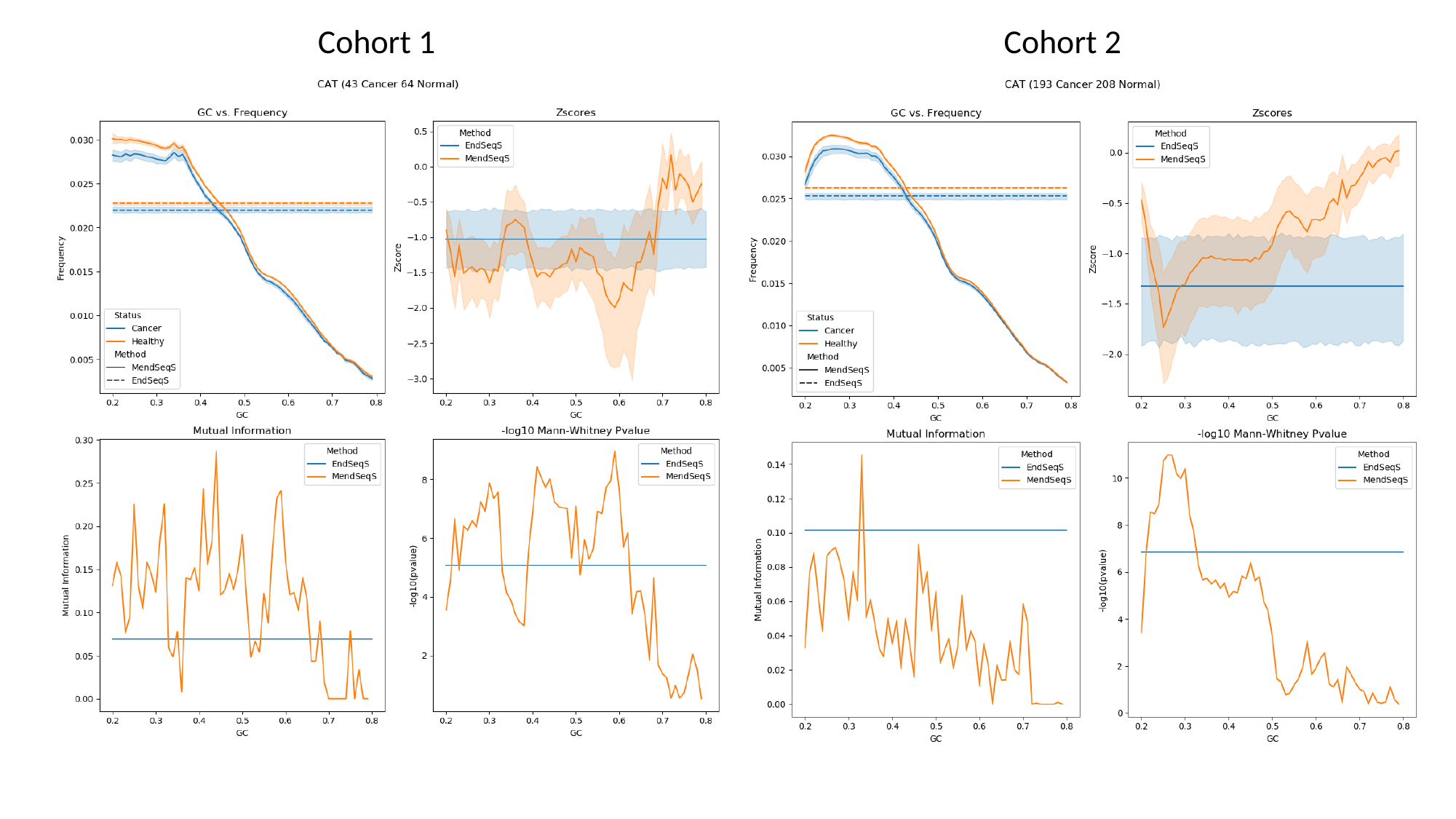

Cohort 2
Cohort 1

### Slide 47
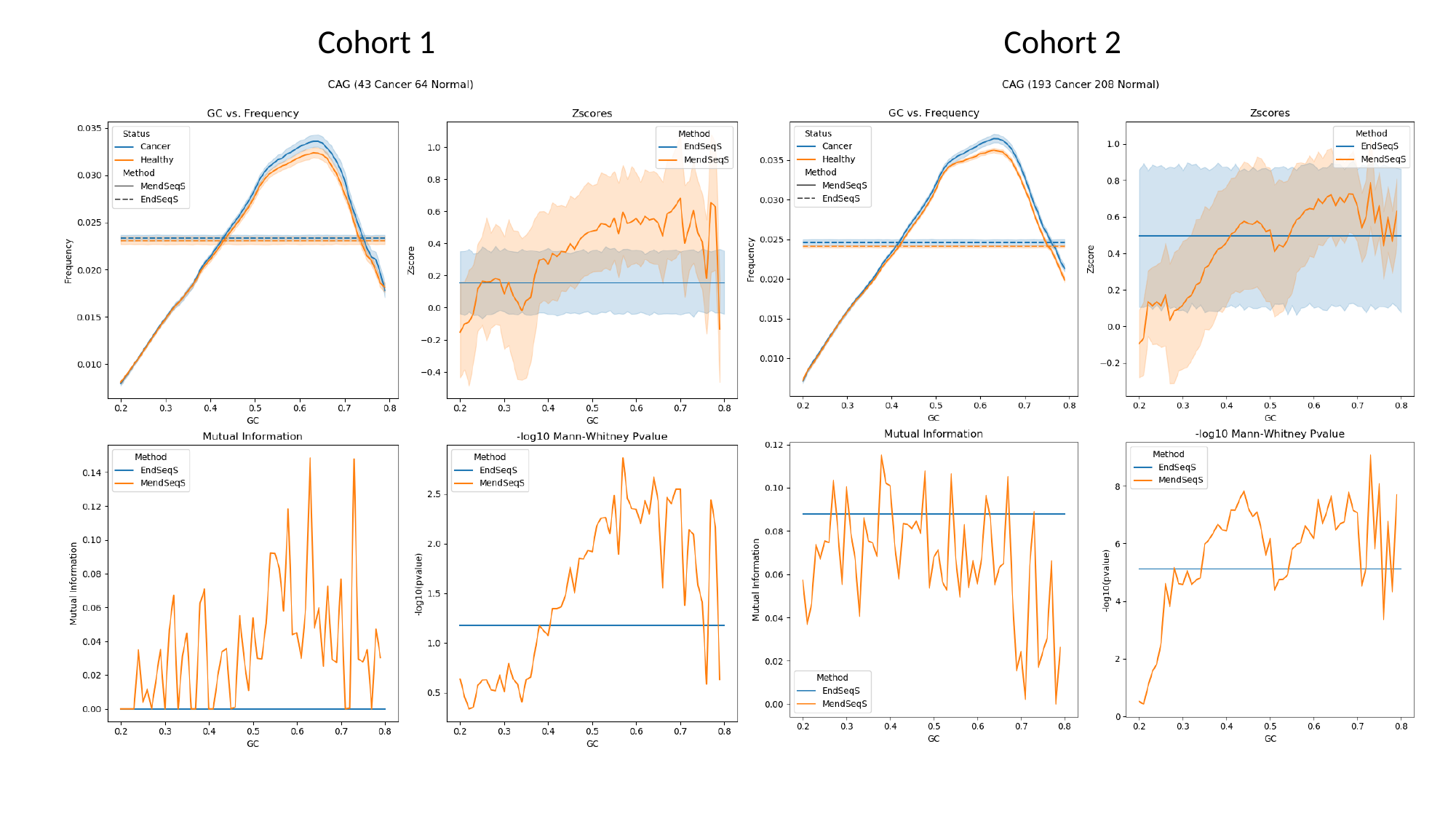

Cohort 2
Cohort 1

### Slide 48
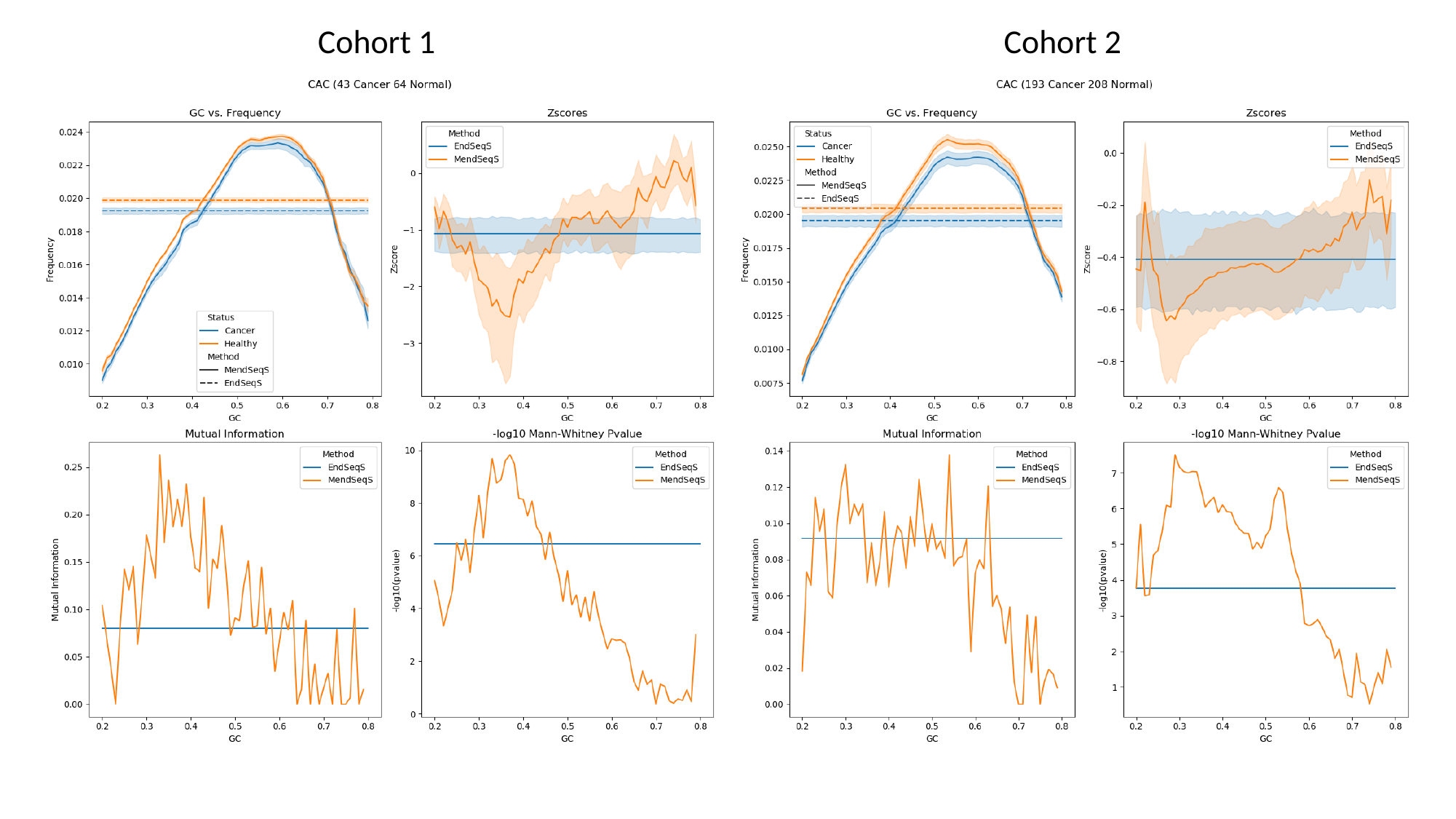

Cohort 2
Cohort 1

### Slide 49
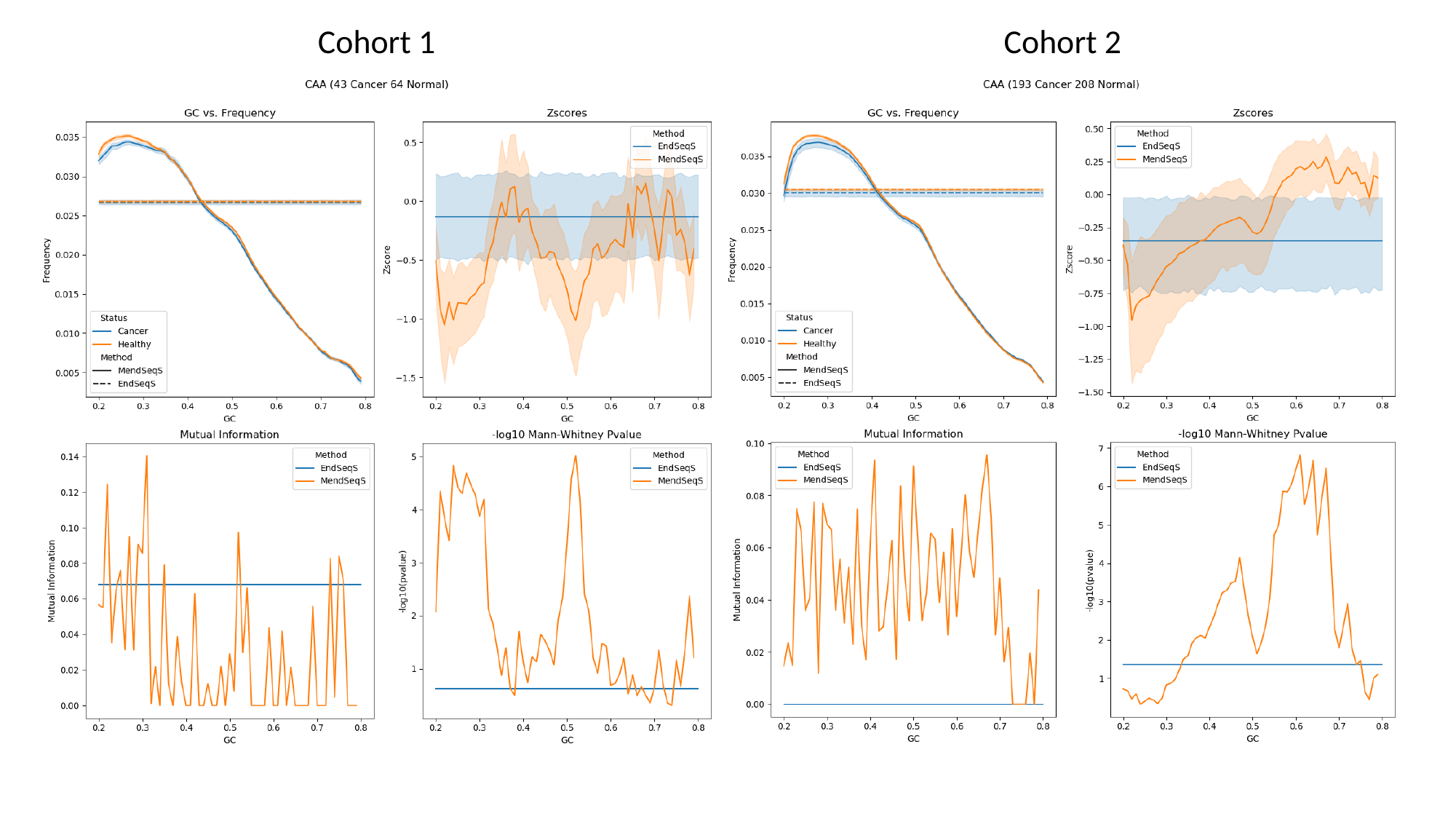

Cohort 2
Cohort 1

### Slide 50
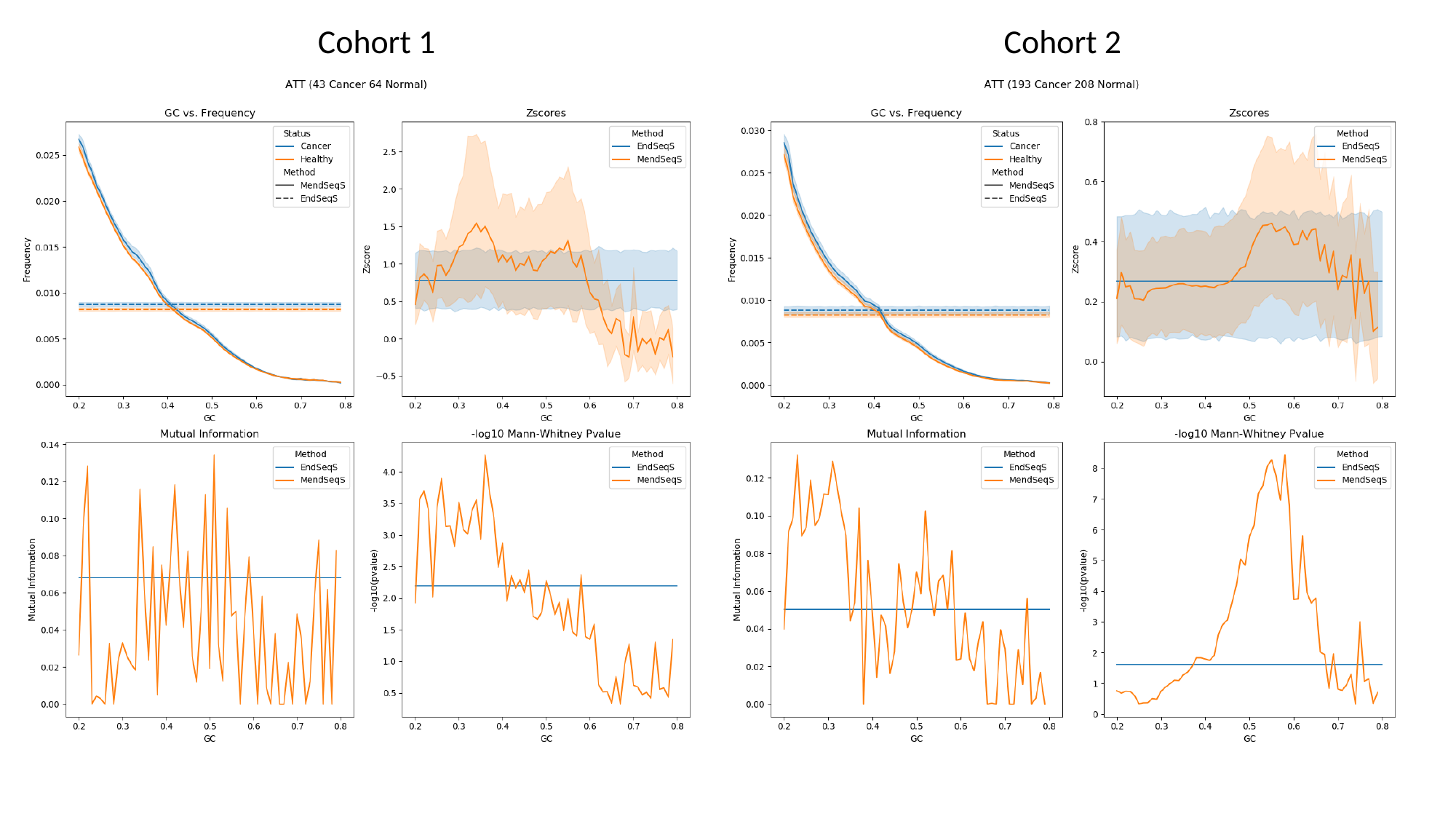

Cohort 2
Cohort 1

### Slide 51
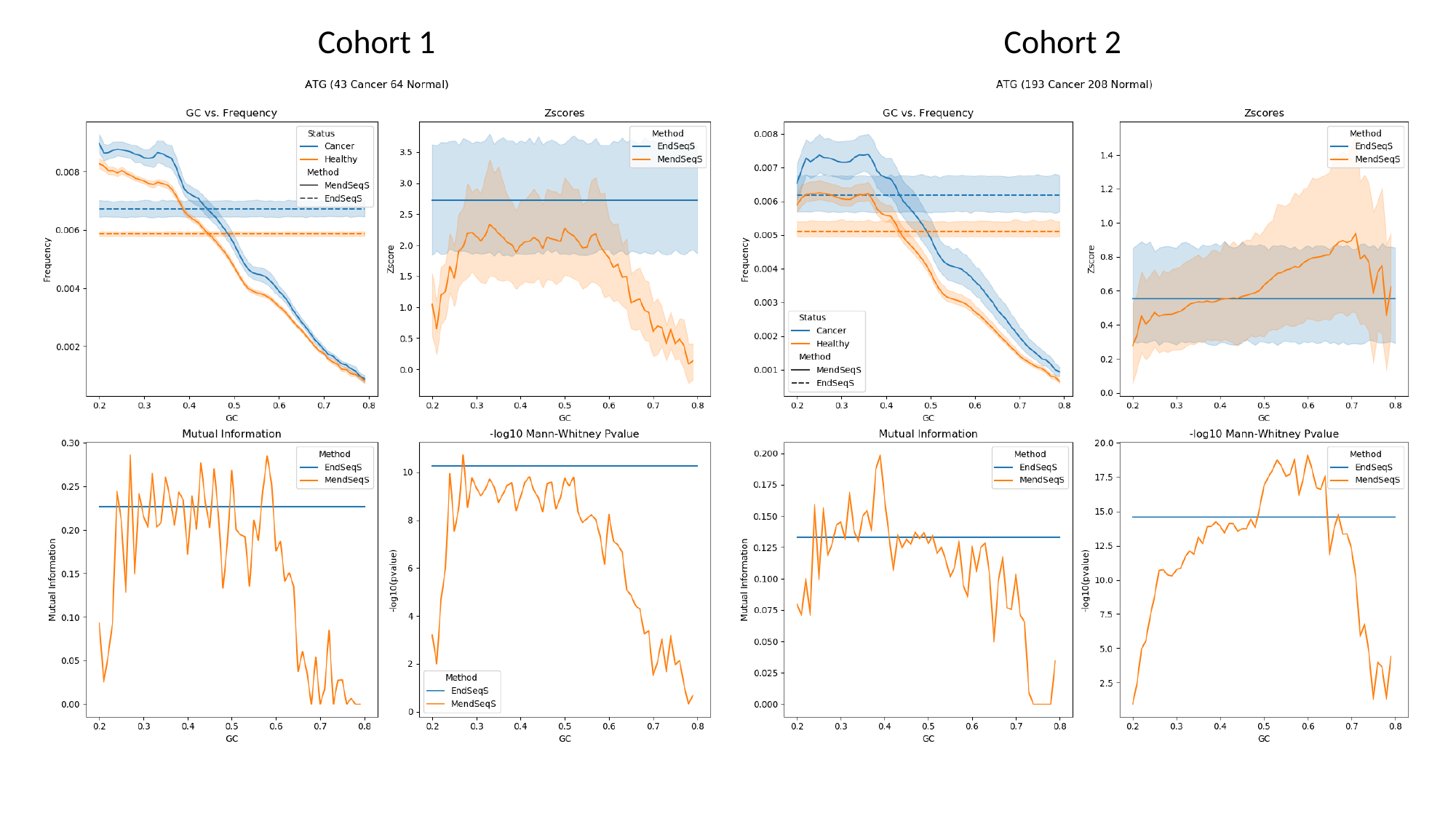

Cohort 2
Cohort 1

### Slide 52
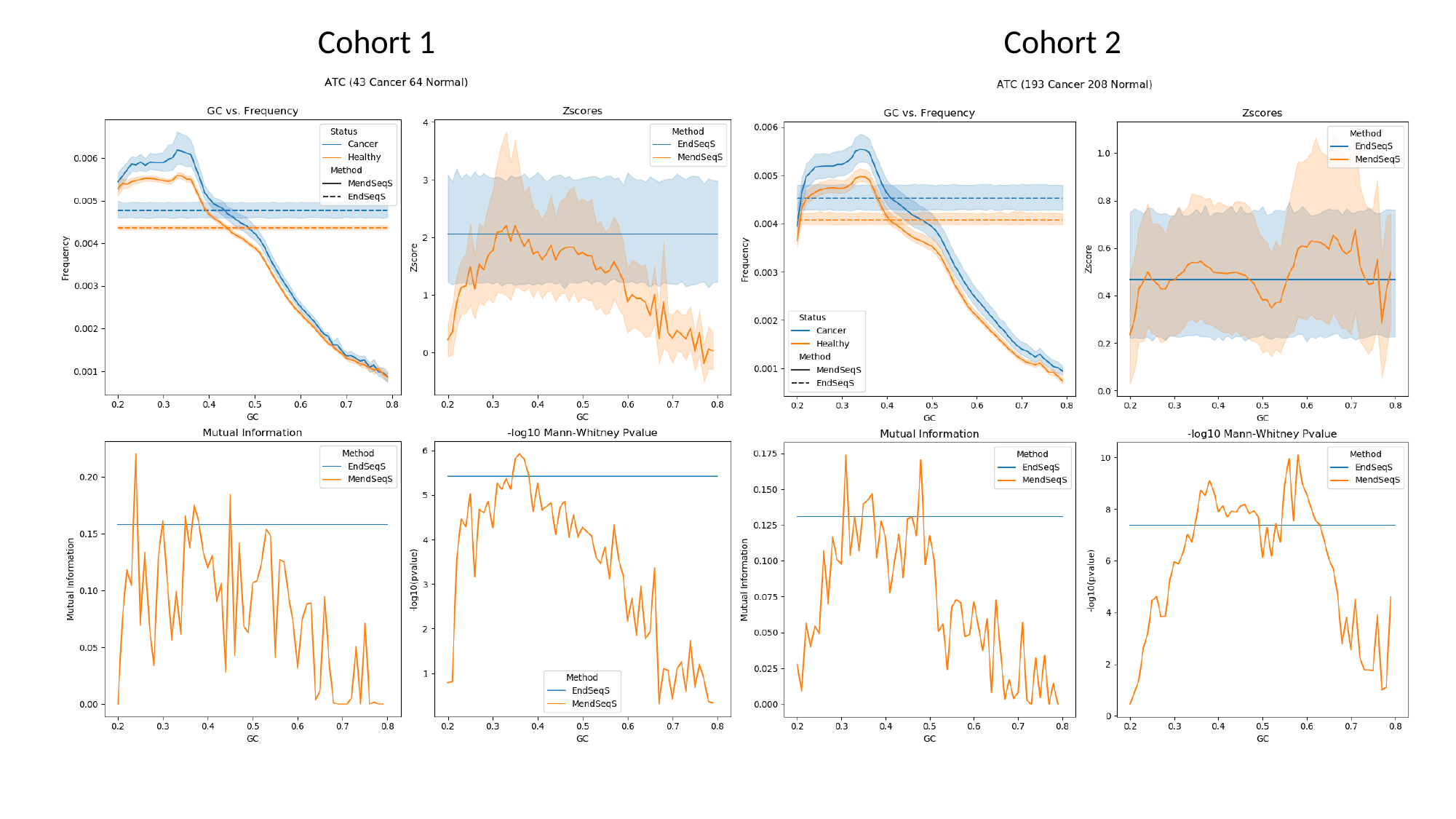

Cohort 2
Cohort 1

### Slide 53
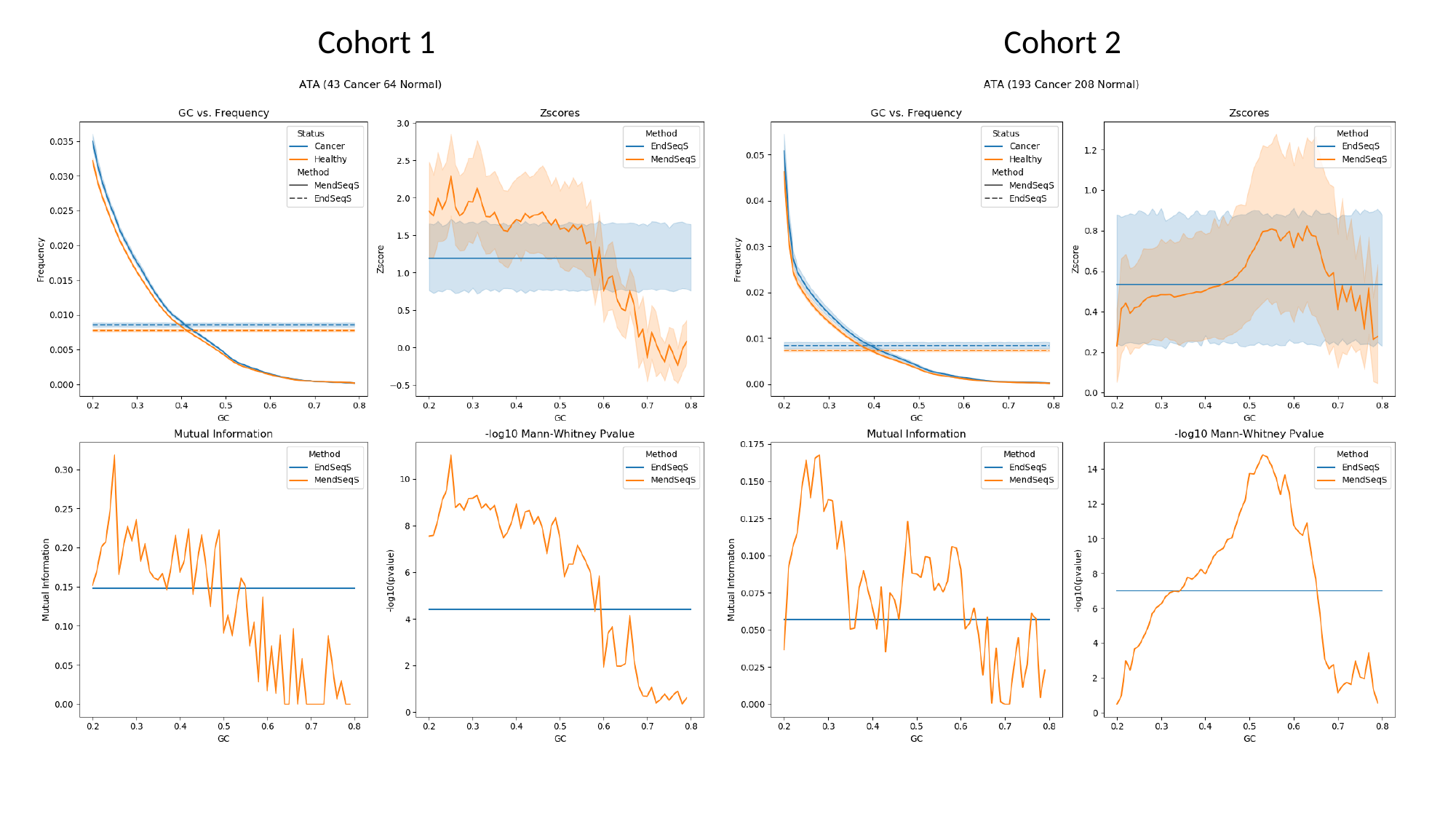

Cohort 2
Cohort 1

### Slide 54
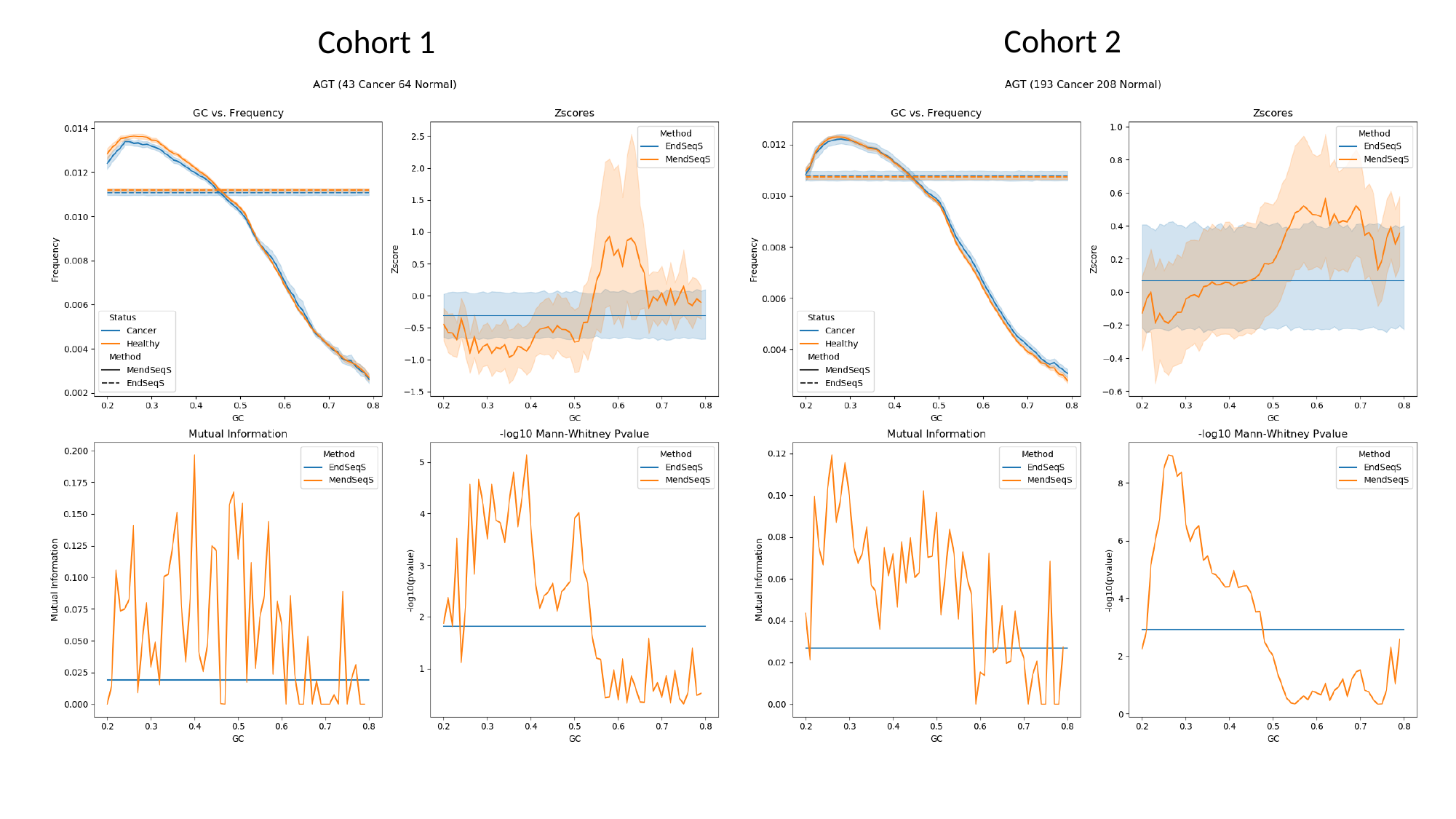

Cohort 2
Cohort 1

### Slide 55
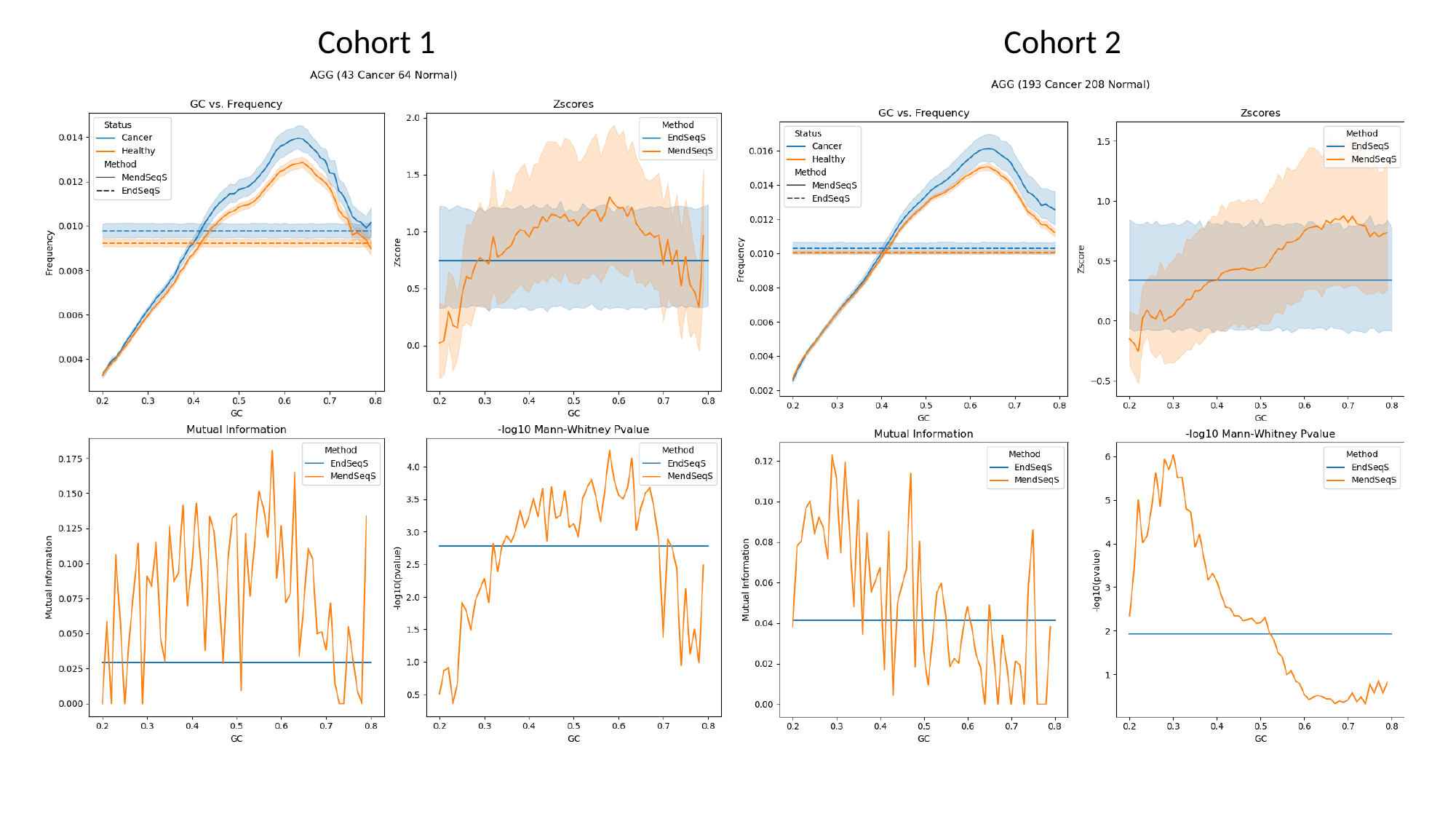

Cohort 2
Cohort 1

### Slide 56
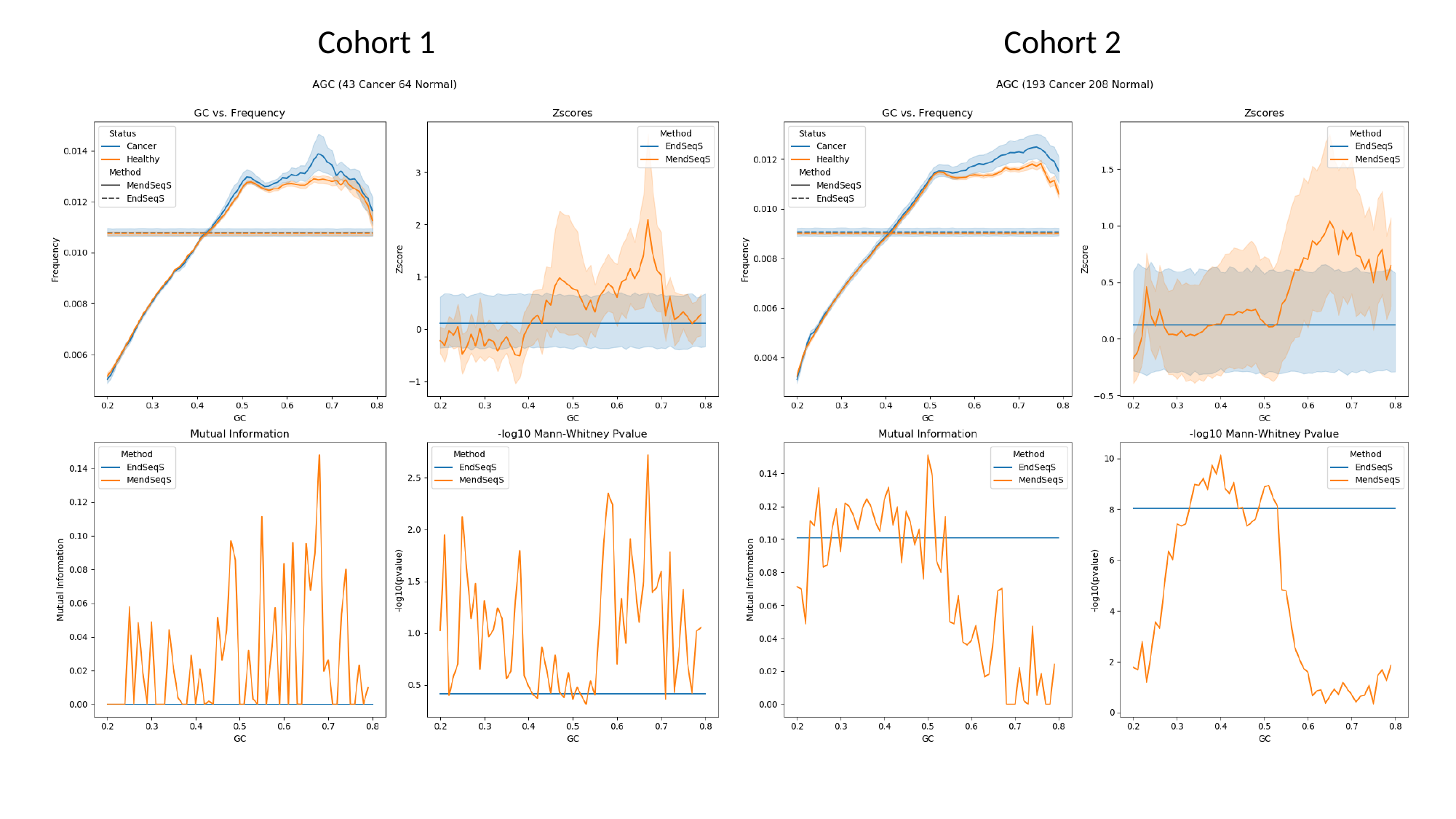

Cohort 2
Cohort 1

### Slide 57
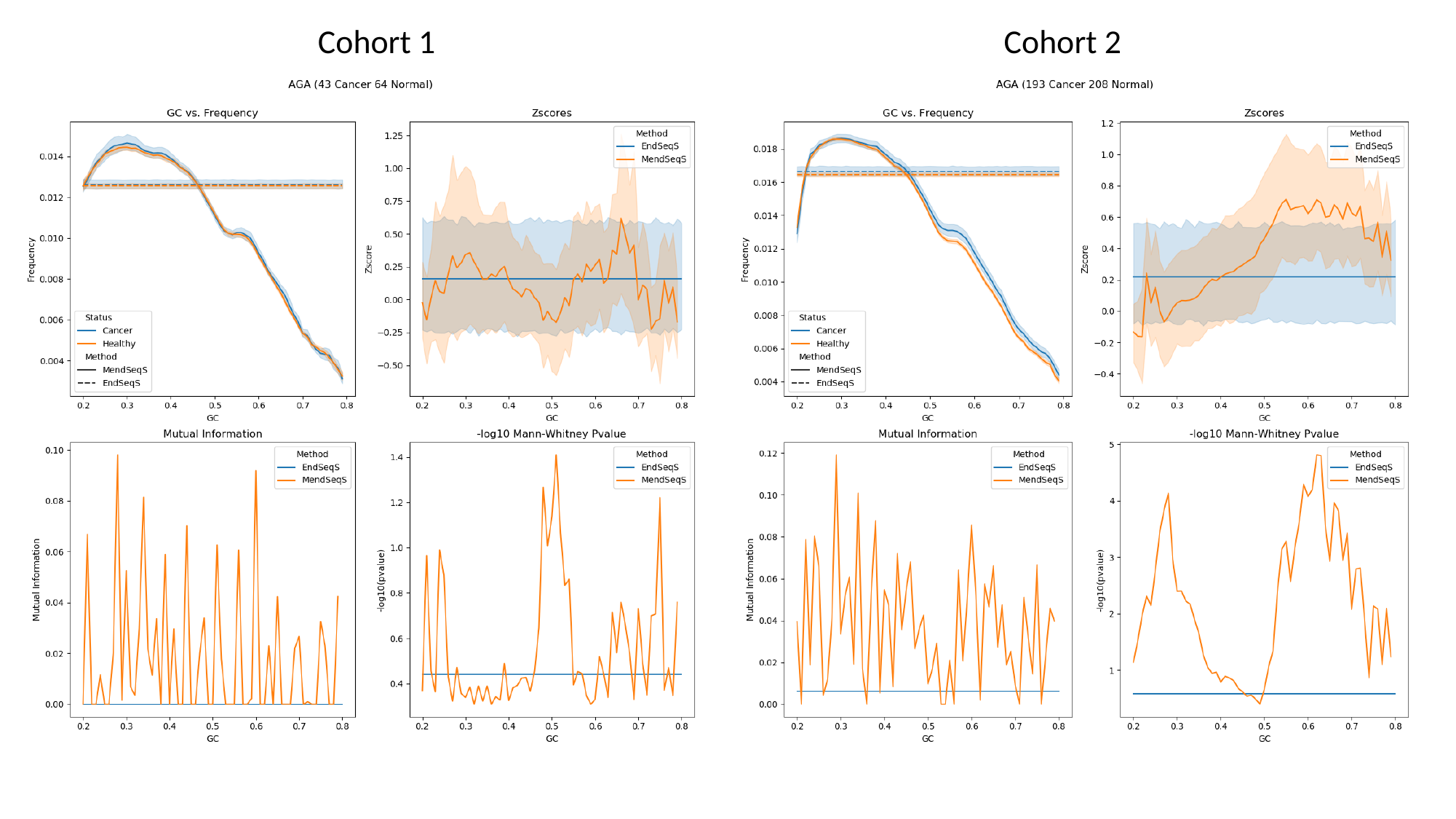

Cohort 2
Cohort 1

### Slide 58
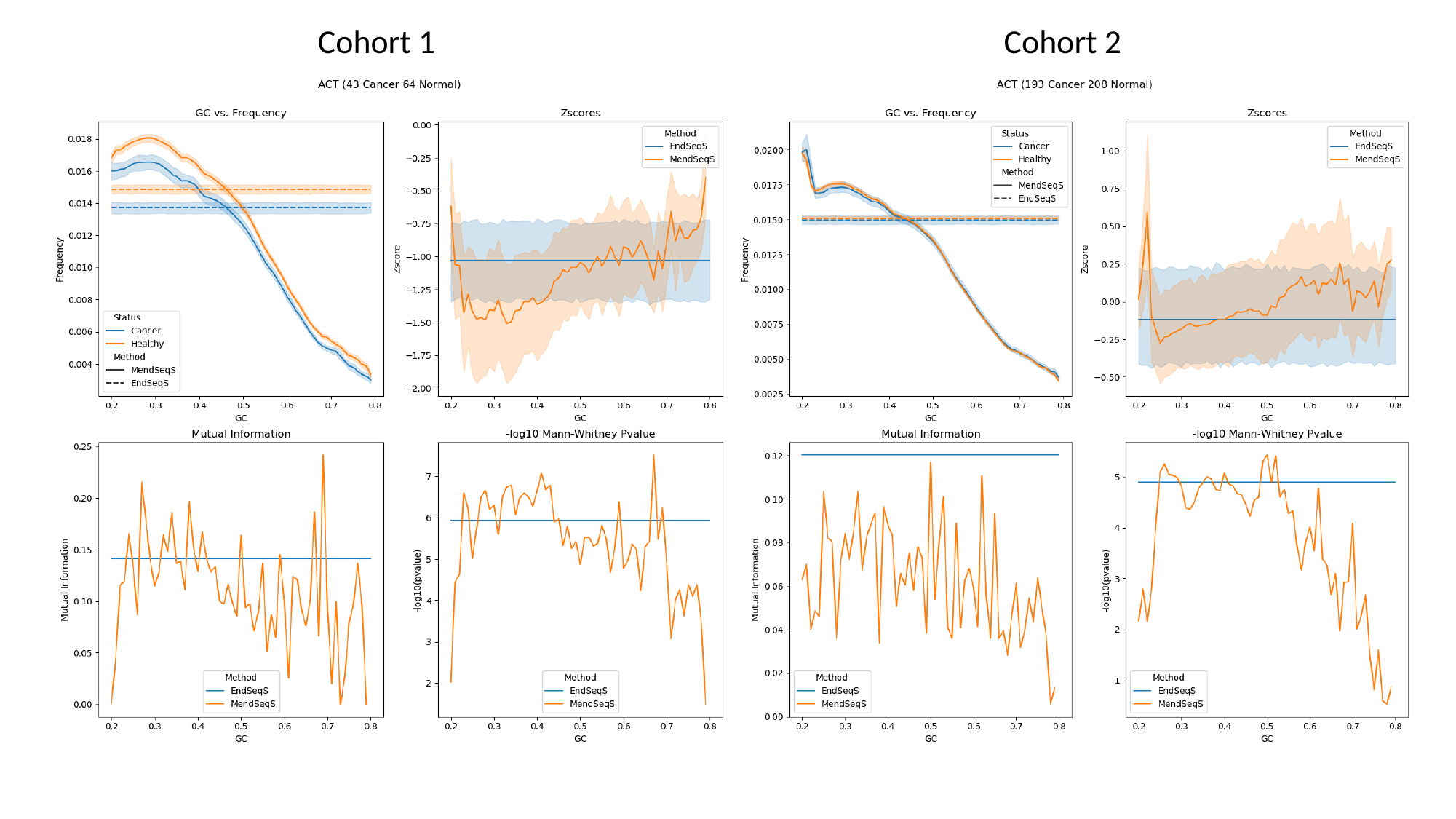

Cohort 2
Cohort 1

### Slide 59
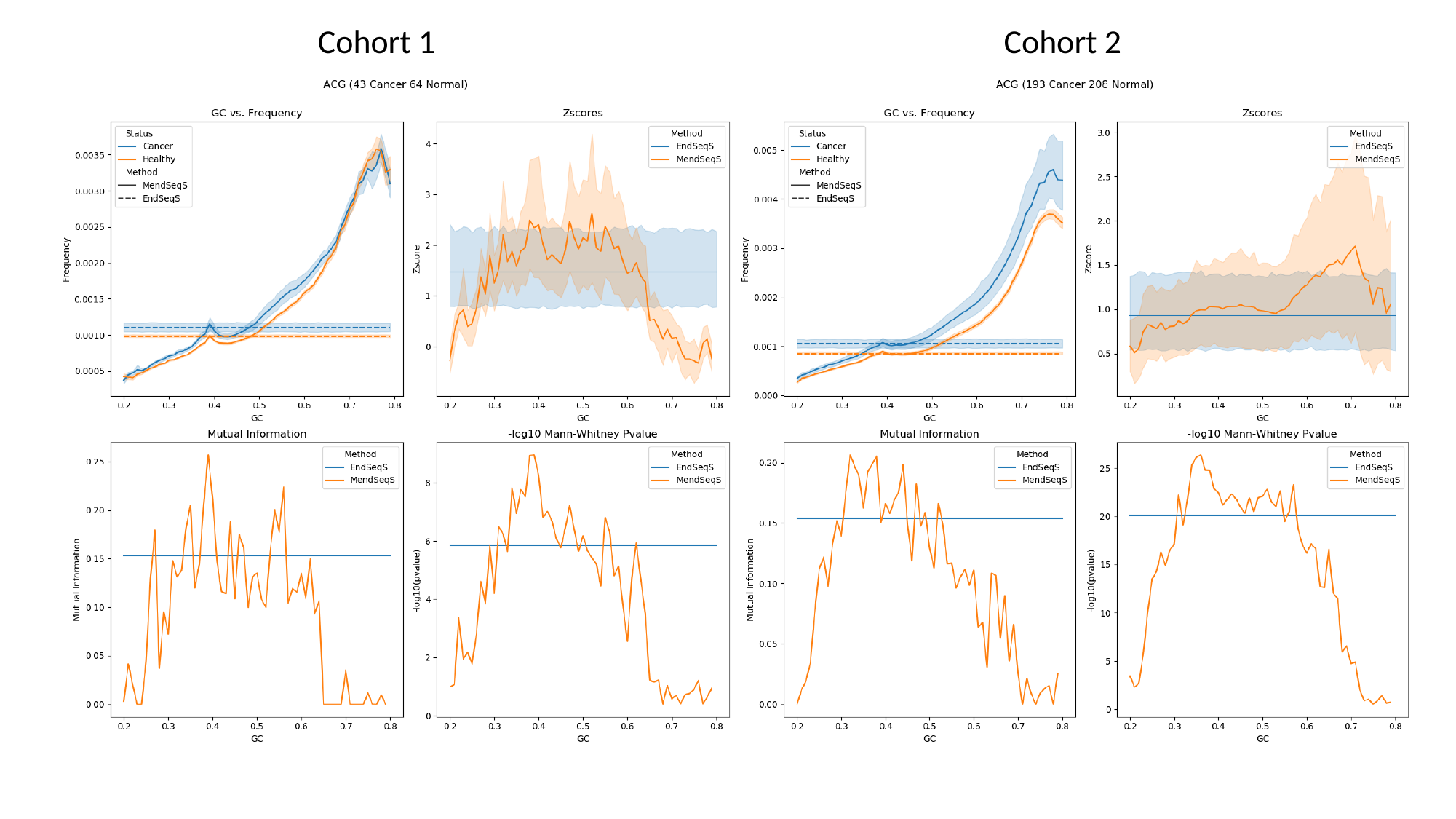

Cohort 2
Cohort 1

### Slide 60
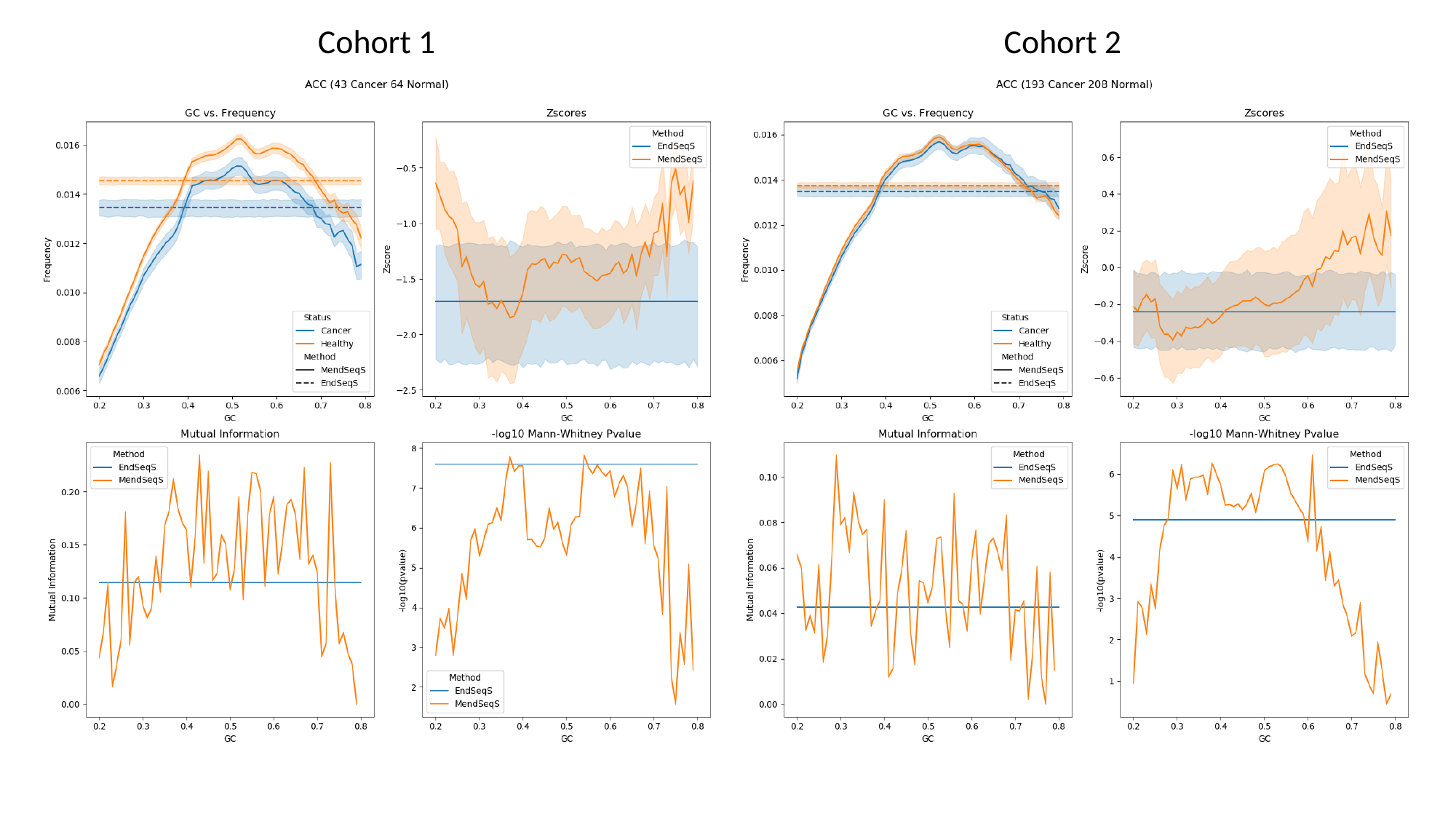

Cohort 2
Cohort 1

### Slide 61
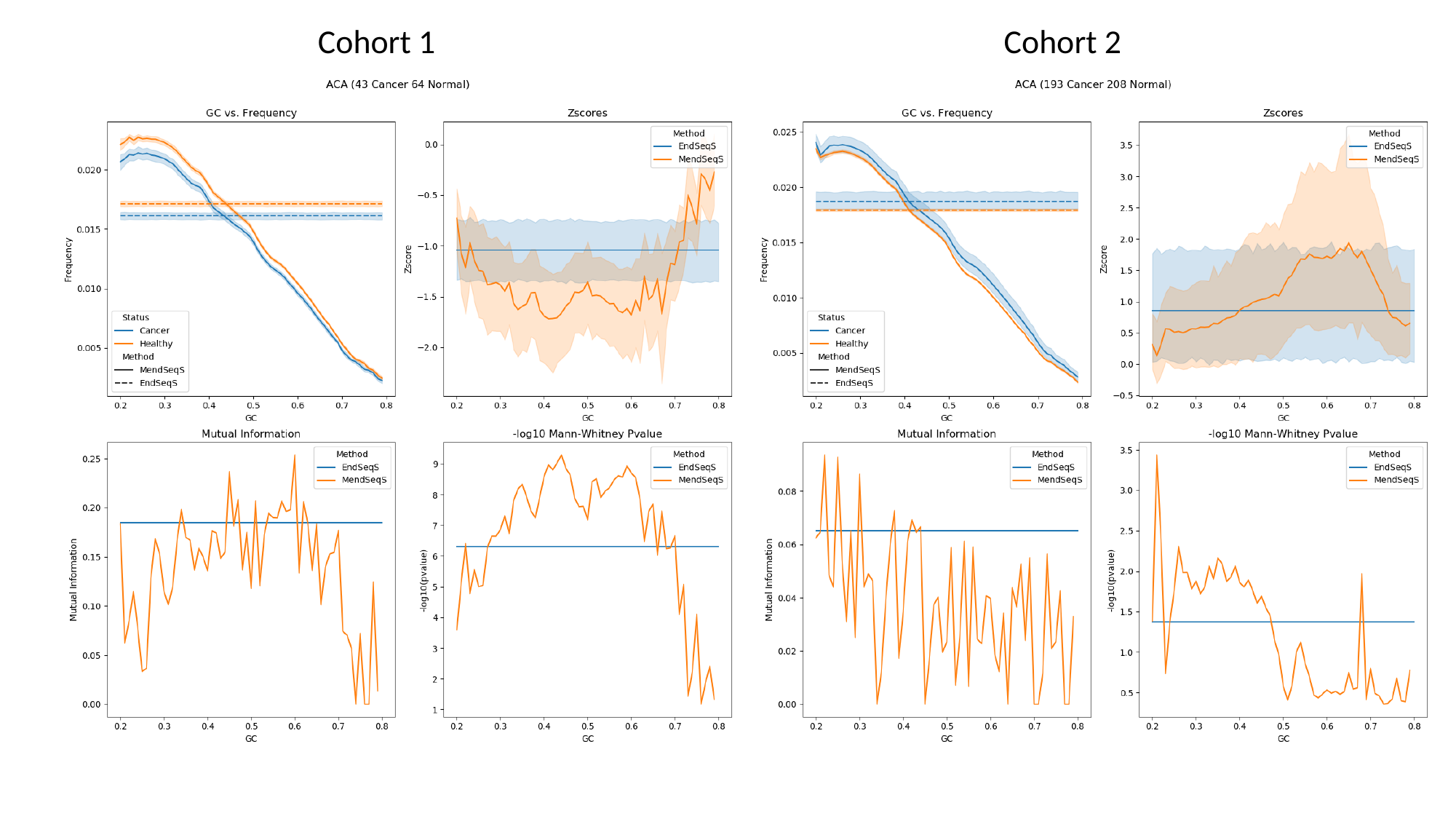

Cohort 2
Cohort 1

### Slide 62
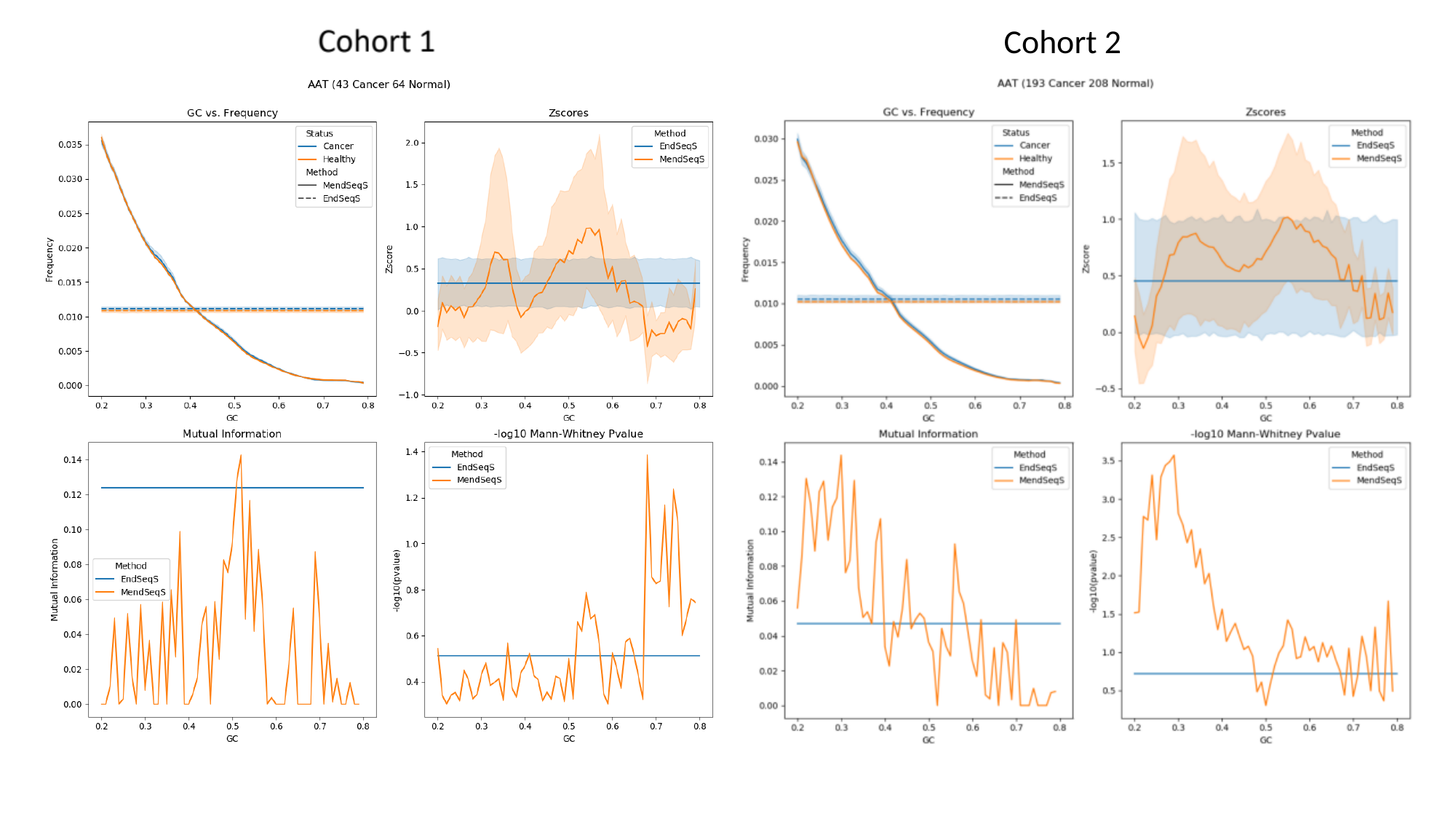

Cohort 2
Cohort 1

### Slide 63
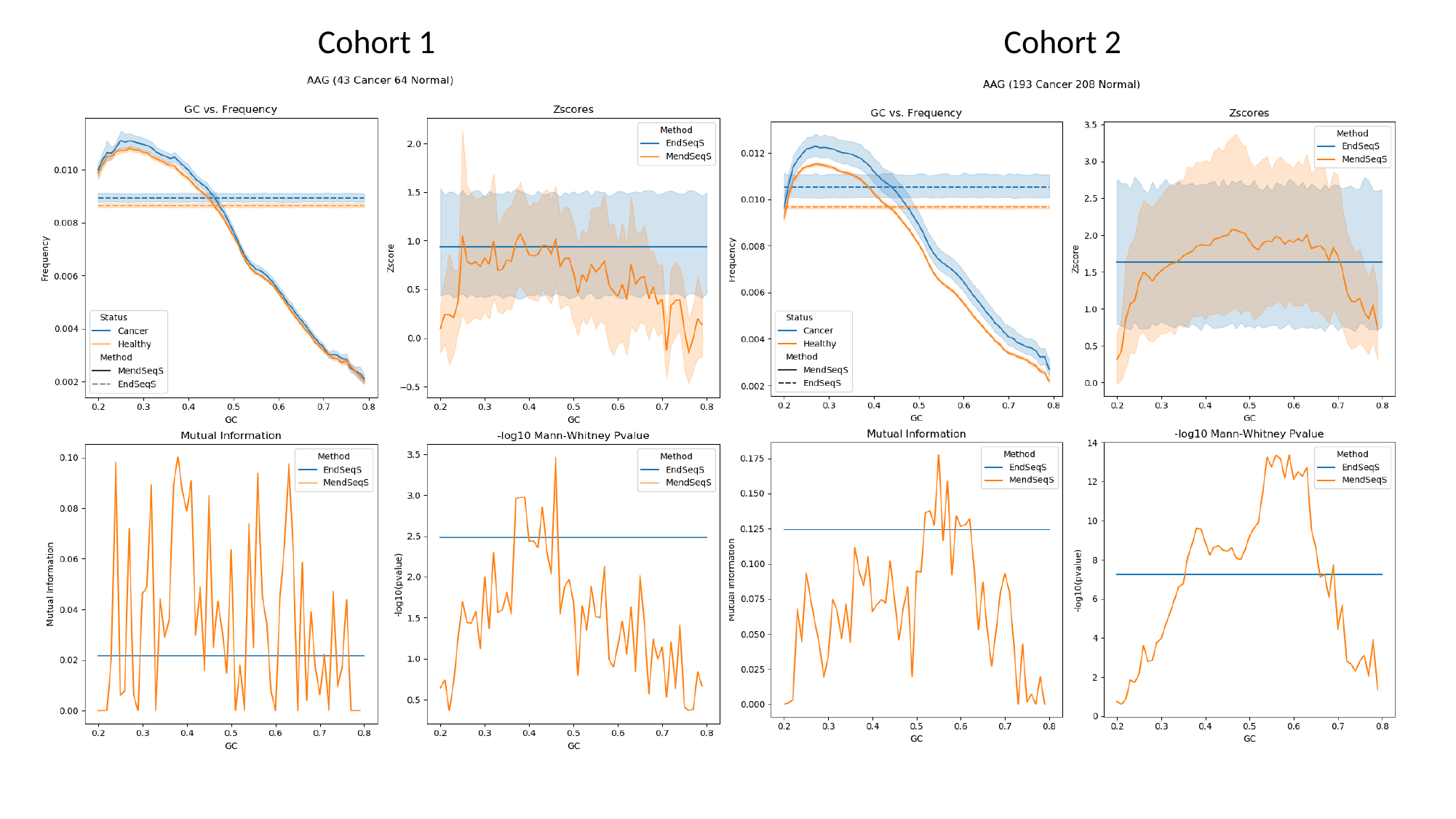

Cohort 2
Cohort 1

### Slide 64
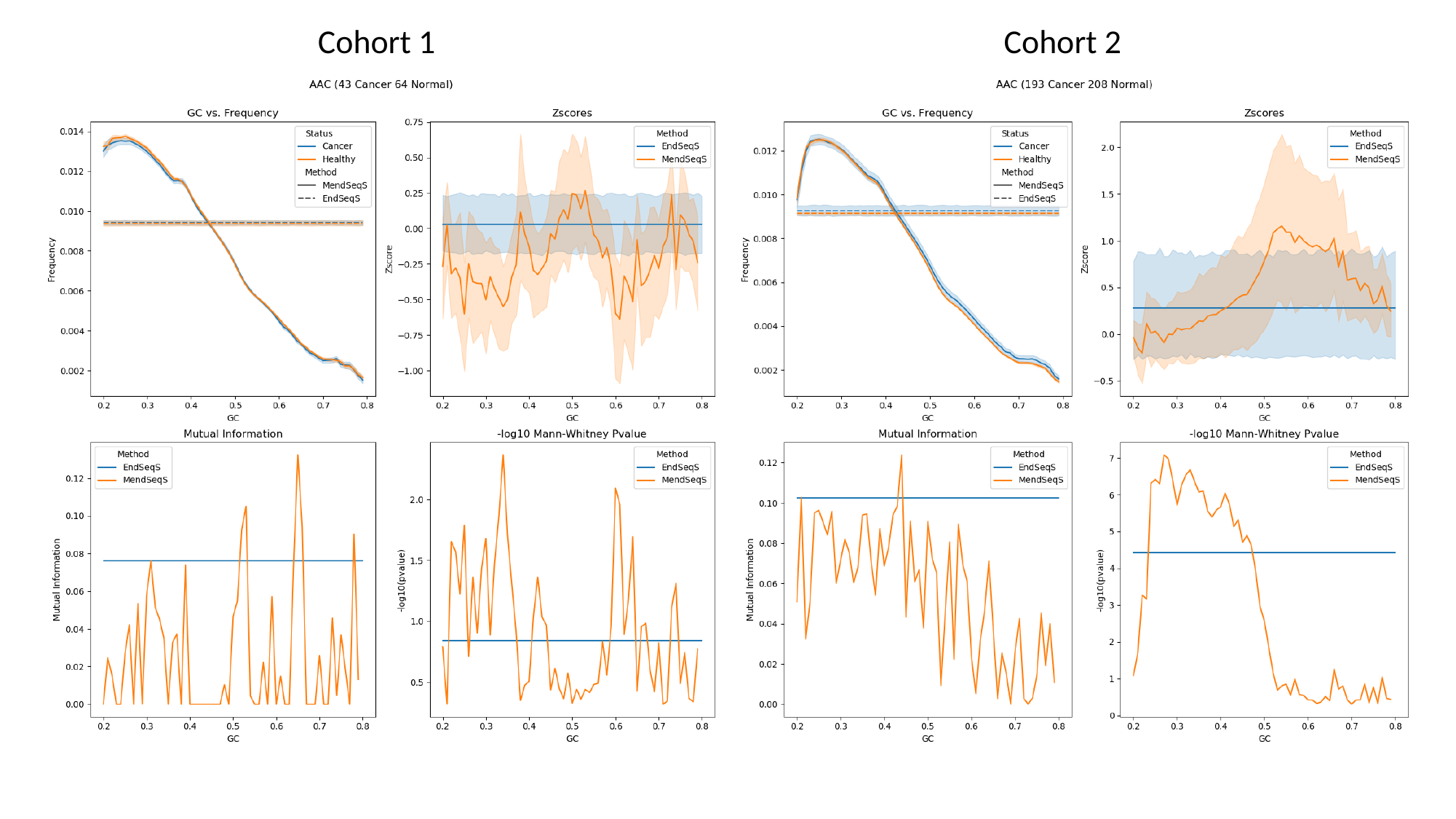

Cohort 2
Cohort 1

### Slide 65
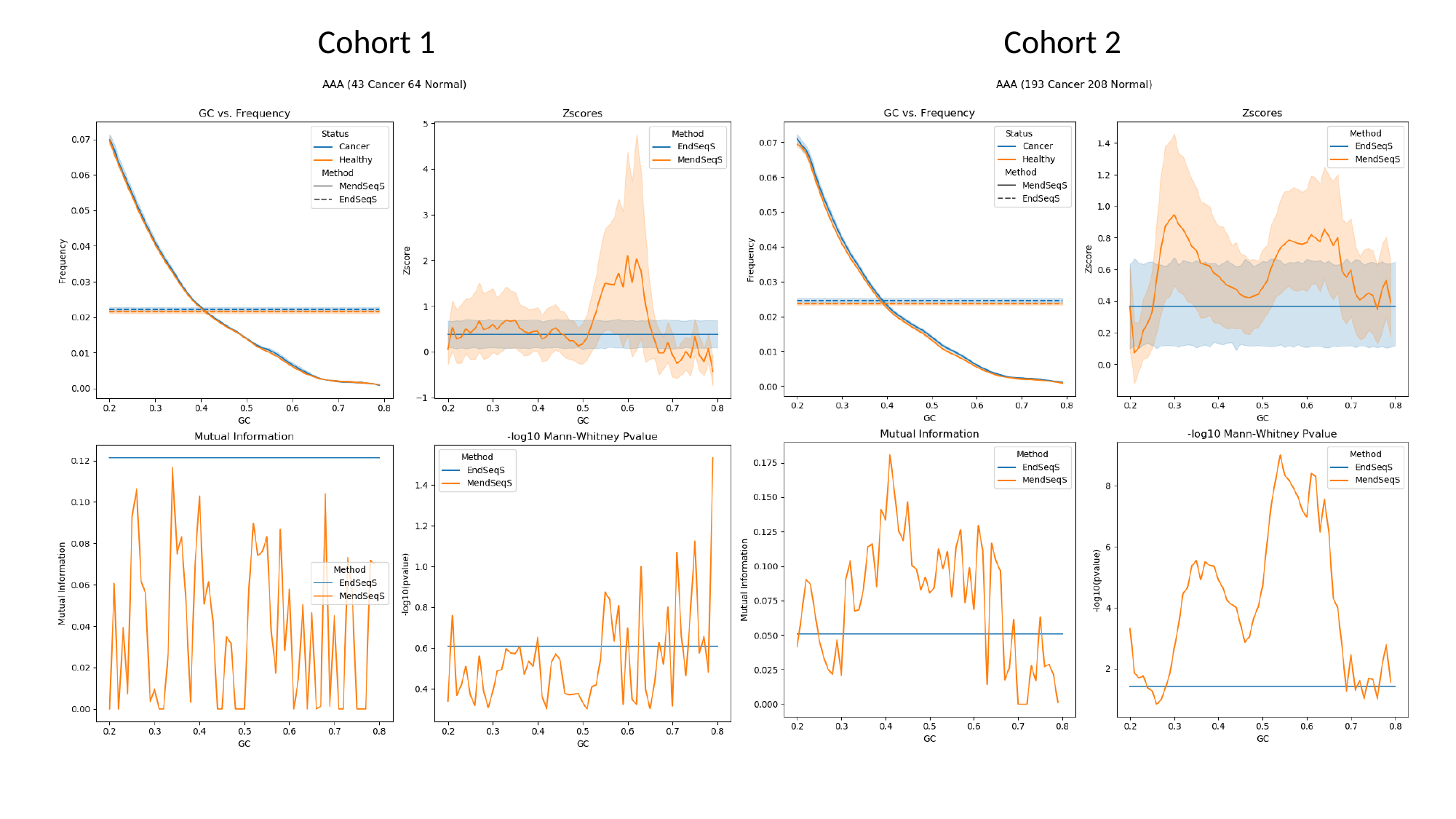

Cohort 2
Cohort 1
